# *In vivo* white matter microstructure in adolescents with early-onset psychosis: a multi-site mega-analysis via the ENIGMA Consortium

**DOI:** 10.1101/2022.02.11.22270818

**Authors:** Claudia Barth, Sinead Kelly, Stener Nerland, Neda Jahanshad, Clara Alloza, Sonia Ambrogi, Ole A Andreassen, Dimitrios Andreou, Celso Arango, Inmaculada Baeza, Nerisa Banaj, Carrie E Bearden, Michael Berk, Hannes Bohman, Josefina Castro-Fornieles, Yann Chye, Benedicto Crespo-Facorro, Elena de la Serna, Covadonga M Díaz-Caneja, Tiril P Gurholt, Catherine E Hegarty, Anthony James, Joost Janssen, Cecilie Johannessen, Erik G Jönsson, Katherine H Karlsgodt, Peter Kochunov, Noemi G Lois, Mathias Lundberg, Anne M Myhre, Saül Pascual-Diaz, Fabrizio Piras, Runar E Smelror, Gianfranco Spalletta, Therese S Stokkan, Gisela Sugranyes, Chao Suo, Sophia I Thomopoulos, Diana Tordesillas-Gutiérrez, Daniela Vecchio, Kirsten Wedervang-Resell, Laura A Wortinger, Paul M. Thompson, Ingrid Agartz

**Affiliations:** Department of Psychiatric Research, Diakonhjemmet Hospital, Oslo, Norway; Norwegian Centre for Mental Disorders Research (NORMENT), Institute of Clinical Medicine, University of Oslo, Oslo, Norway; Department of Psychosis Studies, King’s College London, London, UK; Imaging Genetics Center, Mark & Mary Stevens Neuroimaging & Informatics Institute, Keck School of Medicine, University of Southern California, Marina del Rey, CA, USA; Department of Child and Adolescent Psychiatry, Institute of Psychiatry and Mental Health, Hospital General Universitario Gregorio Marañón, IiSGM, CIBERSAM, Madrid, Spain; Laboratory of Neuropsychiatry, Santa Lucia Foundation IRCCS, Rome, Italy; Norwegian Center for Mental Disorders Research (NORMENT), Division of Mental Health and Addiction, Oslo University Hospital, Oslo, Norway; Centre for Psychiatry Research, Department of Clinical Neuroscience, Karolinska Institutet & Stockholm Health Care Services, Stockholm Region, Stockholm, Sweden; School of Medicine, Universidad Complutense, Madrid, Spain; Department Child and Adolescent Psychiatry and Psychology, 2017SGR881 Institute of Neuroscience, Hospital Clinic Barcelona. CIBERSAM. August Pi i Sunyer Biomedical Research Institute (IDIBAPS), University of Barcelona, Barcelona, Spain; Department of Psychiatry and Biobehavioral Sciences, Semel Institute for Neuroscience and Human Behavior, UCLA, Los Angeles, California, USA; Department of Psychology, UCLA, Los Angeles, California, USA; Deakin University, Institute for Mental and Physical Health and Clinical Translation, School of Medicine, Barwon Health, Geelong, Australia; Department of Neuroscience, Child and Adolescent Psychiatry, Uppsala University, Uppsala, Sweden; Department of Clinical Science and Education Södersjukhuset, Karolinska Institutet, Stockholm, Sweden; Turner Institute for Brain and Mental Health and School of Psychological Sciences, Monash University, Melbourne, Victoria, Australia; Hospital Universitario Virgen del Rocío, Department of Psychiatry, Instituto de Investigación Sanitaria de Sevilla, IBiS, CIBERSAM, Sevilla, Spain; Highfield Unit, Warneford Hospital, Oxford, UK; Department of Psychiatry, University of Oxford, Oxford, UK; Maryland Psychiatric Research Center, Department of Psychiatry, University of Maryland School of Medicine, Baltimore, Maryland, USA; Department of Child and Adolescent Psychiatry, Institute of Psychiatry and Mental Health, Hospital General Universitario Gregorio Marañón, IiSGM, Madrid, Spain; Section of Child and Adolescent Mental Health Research, Division of Mental Health and Addiction, Oslo University Hospital, Oslo, Norway; Magnetic Resonance Imaging Core Facility, August Pi i Sunyer Biomedical Research Institute (IDIBAPS), University of Barcelona, Barcelona, Spain; Menninger Department of Psychiatry and Behavioral Sciences, Baylor College of Medicine, Houston, TX, USA; Department of Radiology. Marqués de Valdecilla University Hospital, Valdecilla Biomedical Research Institute IDIVAL, Spain; Advanced Computing and e-Science, Instituto de Física de Cantabria (UC-CSIC), Santander (Cantabria), Spain

**Author notes:** **Corresponding author:** Claudia Barth, PhD.

**Keywords:** Early-onset psychosis, diffusion tensor imaging, multi-site analysis, psychosis, brain white matter, adolescence

## Abstract

**Background:** Emerging evidence suggests brain white matter alterations in adolescents with early-onset psychosis (EOP; age of onset <18 years). However, as neuroimaging methods vary and sample sizes are modest, results remain inconclusive. Using harmonized data processing protocols and a mega-analytic approach, we compared white matter microstructure in EOP and healthy controls (CTR) using diffusion tensor imaging (DTI).

**Methods:** Our sample included 321 adolescents with EOP (mean age: 16.3 ± 1.4 years, 46.4% females) and 265 adolescent CTR (mean age: 16.0 ± 1.7 years, 57.7% females) pooled from nine sites. All sites extracted mean fractional anisotropy (FA), mean diffusivity (MD), radial diffusivity (RD), and axial diffusivity (AD) for 25 white matter regions of interest per participant. ComBat harmonization was performed for all DTI measures to adjust for scanner differences. Multiple linear regression models were fitted to investigate case-control differences and associations with clinical variables in regional DTI measures.

**Results:** We found widespread lower FA in EOP compared to CTR, with the largest effect sizes in the superior longitudinal fasciculus (Cohen’s *d* = 0.37), posterior corona radiata (*d* = 0.32), and superior fronto□occipital fasciculus (*d* = 0.31). We also found widespread higher RD and more localized higher MD and AD. We detected significant effects of diagnostic subgroup, sex, and duration of illness, but not medication status.

**Conclusion:** Using the largest EOP DTI sample to date, our findings suggest a profile of widespread white matter microstructure alterations in adolescents with EOP, most prominently in male patients with early-onset schizophrenia and patients with a shorter duration of illness.

## INTRODUCTION

Brain white matter alterations are well-documented in adults with psychotic disorders. A recent meta-analysis from the Enhancing Neuro Imaging Genetics through Meta-Analysis (ENIGMA) Consortium (n = 4,322) reported widespread lower fractional anisotropy (FA) in adult patients with schizophrenia relative to healthy controls, with the largest effect sizes in the anterior corona radiata (*d* = 0.40) and corpus callosum (*d* = 0.39) (1). Emerging evidence suggests similar alterations in adolescents with early-onset psychosis (EOP). However, as methods vary across studies and sample sizes are small to modest (ranging from 12 to 55 participants), results in EOP remain inconclusive (for review see (2)). To address this issue, the ENIGMA EOP Working Group initiated the largest collaborative mega-analysis of white matter structure in EOP to date.

The term EOP covers rare and heterogeneous psychiatric disorders, affecting 0.05 - 0.5% of the world’s population (3, 4) and encompasses both the schizophrenia and affective psychosis spectra. Compared to adult-onset psychosis, patients with EOP show worse long-term prognosis (5-8), and EOP significantly contributes to the lifetime disease burden for adolescents (9). Psychotic symptoms in EOP emerge before 18 years of age, during adolescence (10)- a sensitive period for brain development. To date, there is insufficient knowledge on how brain maturation is linked to the emergence of psychosis and neuroimaging studies on these co-occurring processes are important. While a few magnetic resonance imaging (MRI) studies in EOP report grey matter brain abnormalities (11-14), including a recent multisite mega-analysis from the ENIGMA EOP Working Group (15), less is known about putative white matter alterations. This is a critical research gap, as understanding how white matter microstructure is affected in EOP may provide important insights into the pathophysiology of psychotic disorders during adolescent brain development.

Microstructural properties of white matter are commonly studied using diffusion tensor imaging (DTI), which maps the Brownian movement of water molecules in the brain *in vivo*. Common DTI measures include FA and mean, axial, and radial diffusivity (MD, AD, RD). While FA is a summary measure that reflects the degree of diffusion directionality, AD describes diffusion along the primary axis, and RD characterizes diffusion perpendicular to it (16). MD is a measure of overall diffusion within a voxel. Although FA is generally sensitive to microstructural changes, it is not specific to the type of change (e.g., radial or axial) (16). Both AD and RD have been associated with different putative biological underpinnings: lower AD has been linked to axonal damage (16) and higher RD to disruptions in myelination (17). These DTI measures change throughout the lifespan, with FA increasing and RD and MD decreasing throughout adolescence until early adulthood in healthy individuals (18, 19). Sex differences in this pattern also exist: females show changes in white matter microstructure mainly during mid-adolescence, while white matter changes in males appear to occur from childhood through early adulthood (20). The trajectories of AD are less well known (18, 19).

Several studies have used DTI to compare white matter microstructure in youth with EOP with healthy controls, predominantly focusing on early-onset schizophrenia (EOS (2)). Most studies found widespread lower FA in patients relative to controls. However, the white matter tracts implicated were highly variable across studies (21-26). Common DTI measures beyond FA have rarely been explored. The low degree of spatial overlap between the studies may stem from phenotypic heterogeneity, such as differences in disease severity and duration, medication history, and comorbidities. Small sample sizes and differences in MRI data acquisition, processing, and analysis may further influence study outcomes.

The ENIGMA-EOP Working Group aims to address some of the methodological issues in prior MRI studies and increase statistical power by pooling data worldwide for the largest coordinated analysis on brain white matter in EOP to date. Standardized protocols for image processing, quality control, and statistical analyses were applied using the ENIGMA-DTI protocol for multi-site DTI harmonization (27).

The primary goal of the present study was to identify white matter differences in a large sample of 321 adolescents with EOP and 265 healthy controls across nine cohorts worldwide using a mega-analytic approach (28). We further included a complementary meta-analysis to illustrate between-cohort heterogeneity and allow for a direct comparison to FA findings in adult patients with schizophrenia (1). The modulating effects of sex and clinical covariates such as medication use, symptom severity, and illness duration on diffusion measures were also investigated. Based on previous findings (2), we hypothesized that adolescents with EOP would show widespread lower FA relative to healthy controls. As consistent evidence for associations between DTI measures and clinical covariates in EOP is lacking (2), follow-up analyses were exploratory in nature.

## METHODS & MATERIALS

### Study sample

The ENIGMA-EOP Working Group obtained case-control data from nine cohorts across seven countries (for information on each cohort, please see Supplementary Table S1, S2 and S3), yielding imaging and clinical data on a combined total of 321 adolescents with early-onset psychosis (EOP) and 265 age-matched healthy controls (CTR). Participants were aged 12 to 18 years at MRI image acquisition. The EOP diagnostic subgroups consisted of individuals with early-onset schizophrenia (EOS; n = 180), affective psychosis (AFP; n = 95), and other psychosis (OTP; n = 46). Diagnoses were determined using either the Diagnostic and Statistical Manual of Mental Disorders (DSM)-IV or the International Classification of Diseases (ICD)-10. To assess the presence and severity of symptoms, five cohorts used the Positive and Negative Syndrome Scale (PANSS (29)), and two cohorts used the Scale for the Assessment of Negative/Positive Symptoms (SANS (30)/SAPS (31)). Two cohorts did not acquire PANSS, SANS/SAPS or equivalent scores. Site-wise inclusion and exclusion criteria are presented in Supplementary Table S2. All study participants and/or their legal guardians provided written informed consent with approval from local institutional review boards and the respective ethics committees. The study was conducted in accordance with the Declaration of Helsinki.

### Image processing and analysis

MRI scanner and acquisition parameters for each site are detailed in Supplementary Table S3. Preprocessing of diffusion-weighted images, including eddy current correction, echo-planar imaging (EPI) induced distortion correction, initial quality control, and tensor fitting were performed locally at each site using tools and processes suitable for the acquired data (http://enigma.ini.usc.edu/protocols/dti-protocols/). Protocols for image processing and quality control procedures are available via the ENIGMA-DTI website (http://enigma.ini.usc.edu/ongoing/dti-working-group/) and on the ENIGMA GitHub page (https://github.com/ENIGMA-git/). Fractional anisotropy (FA), mean diffusivity (MD), radial diffusivity (RD), and axial diffusivity (AD) were obtained for 25 bilateral (or mid-sagittal) regions of interest (ROI) from the Johns Hopkins University White Matter atlases (JHU; see Table 1 for a list of white matter regions). While all nine sites provided FA measures (n = 586), MD, RD, and AD data were obtained from eight sites only (n = 509).

**Table 1.** Twenty-five white matter regions of interest.

| **Brain white matter tract** | **Abbreviation** |
| --- | --- |
| Average DTI measure across entire skeleton | Average FA/MD/RD/AD |
| Corpus callosum (body/genu/splenium) | CC/BCC/GCC/SCC |
| Cingulum (cingulate gyrus part) | CGC |
| Perihippocampal cingulum tract | CGH |
| Corona radiata (anterior/posterior/superior) | CR/ACR/PCR/SCR |
| Cortico-spinal tract | CST |
| External capsule | EC |
| Fornix/Stria terminalis | FX/ FXST |
| Internal capsule (anterior/posterior/retrolenticular limb) | IC/ALIC/PLIC/RLIC |
| Inferior fronto-occipital fasciculus | IFO |
| Uncinate fasciculus | UNC |
| Posterior thalamic radiation | PTR |
| Superior fronto-occipital fasciculus | SFO |
| Superior longitudinal fasciculus | SLF |
| Sagittal stratum | SS |

ComBat harmonization was performed for all DTI measures to remove unwanted scanner- and sequence-related variation (32, 33) whilst preserving biological associations in the data (Supplementary Note S1). Empirical Bayes was used to leverage information across each DTI measure, an approach that has been shown to be more robust to outliers of small within-scanner sample sizes (34). Age, sex, and diagnostic group were included as variables of interest. The ComBat-harmonization output for all DTI measures is visualized in Figures S1 and S2.

### Statistical analysis

#### Case-control findings

Case-control differences in FA, MD, AD, and RD across all ROIs were examined using multiple linear regression analysis, correcting for age, age^2^, sex, and linear and nonlinear age-by-sex interactions. All analyses included combined ROIs across both hemispheres as dependent variables. Lateralized results are reported in Supplementary Table S8. We included a complementary meta-analysis, similar to Gurholt and colleagues (15), to illustrate between-cohort heterogeneity and allow for a direct comparison to FA findings in adult patients with schizophrenia (see Supplementary Note S2 and section “*Comparison to adult patients with schizophrenia”*).

### Controlling for average, core, and periphery diffusivity measures

To examine global vs. regional white matter effects between diagnostic groups, the main DTI analyses were re-run either co-varying for (i) average, (ii) core, and (iii) periphery DTI measures. For instance, average FA constitutes FA averaged across the entire white matter skeleton, excluding gray matter. The “core” is defined as the region within the skeleton labeled by the JHU white matter atlas, the rest of the skeleton is defined as “periphery” or non-JHU. The average FA in the standard ENIGMA-DTI template consists of 112,889 voxels, while the core consists of 31,742 voxels, less than a third of the average FA. The remaining 81,147 voxels surrounding the core comprise the periphery (non-JHU). Detailed formulas to calculate core and periphery DTI measures have been published in Kelly et al. 2018 (1).

### Diagnostic subgroup analysis

For each DTI measure (FA, MD, RD, AD) as dependent variable, separate multiple linear regression models were fitted with diagnostic subgroup as fixed factor (CTR, EOS, AFP, OTP) and age, age^2^, sex, and linear and nonlinear age-by-sex interactions as covariates.

### Sex-by-diagnosis & age-by-diagnosis interactions

To further explore white matter microstructural differences between diagnostic groups across age and sex, we performed follow-up sex-stratified as well as sex-by-diagnosis and age-by-diagnosis interaction analyses. Separate multiple linear regression models for each DTI measure as dependent variable were fitted either including a sex-by-diagnosis or age-by-diagnosis interaction terms. The same covariates as above apply. The sex-stratified case-control models (female only/ males only) were covaried for age and age^2^.

### Associations with medication and other clinical measures

In adolescents with EOP, we tested for effects of duration of illness, age at onset, PANSS scores (negative/positive subscores), current medication use (user vs. non-user) and antipsychotic chlorpromazine equivalents (CPZ, see Woods 2005 http://www.scottwilliamwoods.com/files/Equivtext.doc) on DTI measures. Current medication use included antipsychotics, lithium, antidepressants, and antiepileptics. As age and age at onset are highly correlated (r = 0.70), this linear model was only adjusted for sex, whereas the other models were adjusted for age, age^2^, sex, and linear and nonlinear age-by-sex interactions.

### Comparison to adult patients with schizophrenia

To assess whether the effect sizes of the case-control differences in tract-specific FA differed between adolescent patients with EOP and adult patients with schizophrenia(1), we conducted z-tests using the following formula (35):

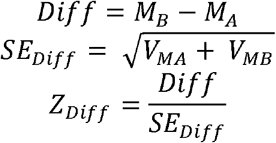

where M_A_ and M_B_ are the estimated Cohen’s *d* effect sizes of the schizophrenia and EOP sample, respectively. V_MA_ and V_MB_ reflect their variances as standard error (SE). The corresponding *p*-value was calculated using (two-tailed test):

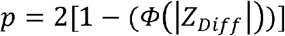

where Φ(Z) is the standard normal cumulative distribution. Meta-analytically derived Cohen’s *d* values for the adult schizophrenia sample were retrieved by SK (1) and compared to meta-and mega-analytically derived Cohen’s *d* values for EOP. To allow for such a comparison, the same covariates were used in both studies, namely age, sex, and linear and non-linear age and sex interactions (age-by-sex interaction, age^2^, and age^2^-by-sex interaction).

All statistical tests were conducted in R, version 4.1.0 (https://www.R-project.org/). We computed the Cohen’s d effect sizes ± standard error across all 25 ROIs from the t-statistics for categorical variables (36). To control for multiple comparisons, effects were considered significant if they survived the Bonferroni correction threshold of 0.05/25=0.002.

## RESULTS

### Demographic and clinical variables

Demographics and clinical characteristics for the whole sample and stratified by diagnostic subgroups are summarized in Table 2 and Table 3, respectively. Sample measures stratified by cohort and sex are displayed in Table S4 and Table S5, respectively.

**Table 2.** Sample demographics and clinical measures.

| Variables | CTR | EOP | p-value | test |
| --- | --- | --- | --- | --- |
| **N** | 265 | 321 |  |  |
| **Age** (years)* | 16.18 [14.88, 17.31] | 16.57 [15.18, 17.32] | 0.147 | KW |
| **Sex**, female N (%) | 153 (57.7) | 149 (46.4) | **0.008** | $\chi^2$ |
| **Handedness**, N (%) | | | 0.331 | $\chi^2$ |
| *Right* | 177 (90.8) | 166 (88.8) |  |  |
| *Left* | 18 (9.2) | 19 (10.2) |  |  |
| *Ambidextrous* | 0 (0.0) | 2 (1.1) |  |  |
| **Diagnostic subgroup**, N (%) |  |  |  |  |
| *EOS* |  | 180 (56.1) |  |  |
| *AFP* |  | 95 (29.6) |  |  |
| *OTP* |  | 46 (14.3) |  |  |
| **PANSS**, negative* |  | 16.00 [12.00, 21.00] |  |  |
| **PANSS**, positive* |  | 20.00 [16.00, 24.00] |  |  |
| **Age of onset** (years)* |  | 15.43 [14.12, 16.71] |  |  |
| **Duration of illness** (years)* |  | 0.60 [0.12, 1.10] |  |  |
| **CPZ** * |  | 200.0 [133.3, 333.3] |  |  |
| **AP user**, N (%) |  | 256 (89.8) |  |  |
| **Lithium user**, N (%) |  | 27 (9.9) |  |  |
| **AD user**, N (%) |  | 57 (25.0) |  |  |
| **AE user**, N (%) |  | 14 (5.2) |  |  |
| **Field strength**, 3T, N (%) | 95 (55.2) | 153 (60.5) | 0.329 | $\chi^2$ |

\*Continuous data in median [Interquartile range] and categorical data as number (%). Abbreviations: CTR = healthy controls, EOP = early-onset psychosis, N = number, EOS = early-onset schizophrenia, AFP = affective psychosis, OTP = other psychosis, PANSS = positive and negative syndrome scale, CPZ = chlorpromazine equivalent, AP = antipsychotics, AD = antidepressants, AE = antiepileptics, KW = Kruskal-Wallis. Significant results are highlighted in bold.

**Table 3.** Demographics and clinical measures, EOP stratified by diagnostic subgroups.

| Variables | CTR | EOS | AFP | OTP | p-value<br>general | p-value<br>EOS vs<br>AFP | p-value<br>EOS vs<br>OTP | p-value<br>AFP vs<br>OTP | test |
| --- | --- | --- | --- | --- | --- | --- | --- | --- | --- |
| N | 265 | 180 | 95 | 46 |  |  |  |  |  |
| Age (years)* | 16.18 [14.88, 17.31] | 16.59 [15.39, 17.25] | 16.66 [15.41, 17.39] | 15.85 [15.00, 17.38] | 0.305 | 0.497 | 0.327 | 0.258 | KW |
| Sex, female N (%) | 153 (57.7) | 70 (38.9) | 54 (56.8) | 25 (54.3) | **0.001** | **0.007** | 0.084 | 0.921 | $\chi^2$ |
| Handedness, N (%) | | | | | 0.564 | 0.773 | 0.510 | 0.957 | $\chi^2$ |
| Right | 177 (90.8) | 120 (89.6) | 24 (88.9) | 22 (84.6) |  |  |  |  |  |
| Left | 18 (9.2) | 12 (9.0) | 3 (11.1) | 4 (15.4) |  |  |  |  |  |
| Ambidextrous | 0 (0.0) | 2 (1.5) | 0 (0.0) | 0 (0.0) |  |  |  |  |  |
| PANSS, negative* |  | 18.00 [14.00, 23.00] | 14.00 [10.00, 19.50] | 15.00 [11.00, 18.00] | **<0.001** | **<0.001** | **0.001** | 0.640 | KW |
| PANSS, positive* |  | 21.00 [18.00, 24.00] | 19.00 [13.00, 24.00] | 16.00 [14.00, 22.00] | **0.003** | **0.033** | **0.001** | 0.293 | KW |
| Age of onset (years)* |  | 15.40 [14.00, 16.44] | 16.05 [14.64, 16.94] | 15.00 [14.20, 16.64] | **0.007** | **0.002** | 0.912 | 0.056 | KW |
| Duration of illness<br>(years)* |  | 1.00 [0.24, 1.49] | 0.21 [0.08, 0.57] | 0.62 [0.22, 1.04] | **<0.001** | **<0.001** | 0.381 | **0.003** | KW |
| CPZ* |  | 200.0 [150.0, 383.5] | 183.3 [100.0, 300.0] | 166.7 [100.0, 258.3] | **0.033** | **0.033** | **0.042** | 0.648 | KW |
| AP user, N (%) | | 156 (90.2) | 71 (89.9) | 29 (87.9) | 0.923 | 1.000 | 0.932 | 1.000 | $\chi^2$ |
| Lithium user, N (%) | | 3 (1.8) | 24 (32.9) | 0 (0.0) | **<0.001** | **<0.001** | 1.000 | **0.001** | $\chi^2$ |
| AD user, N (%) | | 18 (14.4) | 34 (47.9) | 5 (15.6) | **<0.001** | **<0.001** | 1.000 | **0.004** | $\chi^2$ |
| AE user, N (%) | | 11 (6.5) | 3 (4.2) | 0 (0.0) | 0.283 | 0.691 | 0.286 | 0.584 | $\chi^2$ |
| Field strength, 3T, N (%) | 95 (55.2) | 69 (43.7) | 65 (89.0) | 19 (86.4) | **<0.001** | **<0.001** | **0.001** | 1.000 | $\chi^2$ |

\*Continuous data in median [Interquartile range] and categorical data as number (%). Abbreviations: CTR = healthy controls, EOP = early-onset psychosis, EOS = early-onset schizophrenia, AFP = affective psychosis, OTP = other psychosis, N = number, PANSS = positive and negative syndrome scale, CPZ = chlorpromazine equivalent, AP = antipsychotics, AD = antidepressants, AE = antiepileptics, KW = Kruskal-Wallis. Significant results are highlighted in bold.

### Case-control differences

The mega□analysis revealed widespread lower FA in adolescents with EOP relative to healthy controls (see Figure 1, Supplementary Table S6), including the Average FA, CC, GCC, IC, PCR, PTR, RLIC, SFO, and SLF (p ≤ 0.002). Follow-up analyses showed higher MD in the FX and UNC; higher RD in Average RD, CGC, FX, PCR, PTR, SLF, and UNC; and higher AD in the FX in adolescents with EOP relative to CTR (p ≤ 0.002, see Figure 1). The complementary meta-analysis of case-control FA differences corroborated the significant effect for the SLF (Supplementary Table S15). Forest plots illustrate the variability among sites (Supplementary Figure S7), suggesting a great degree of heterogeneity.

**Figure 1.**
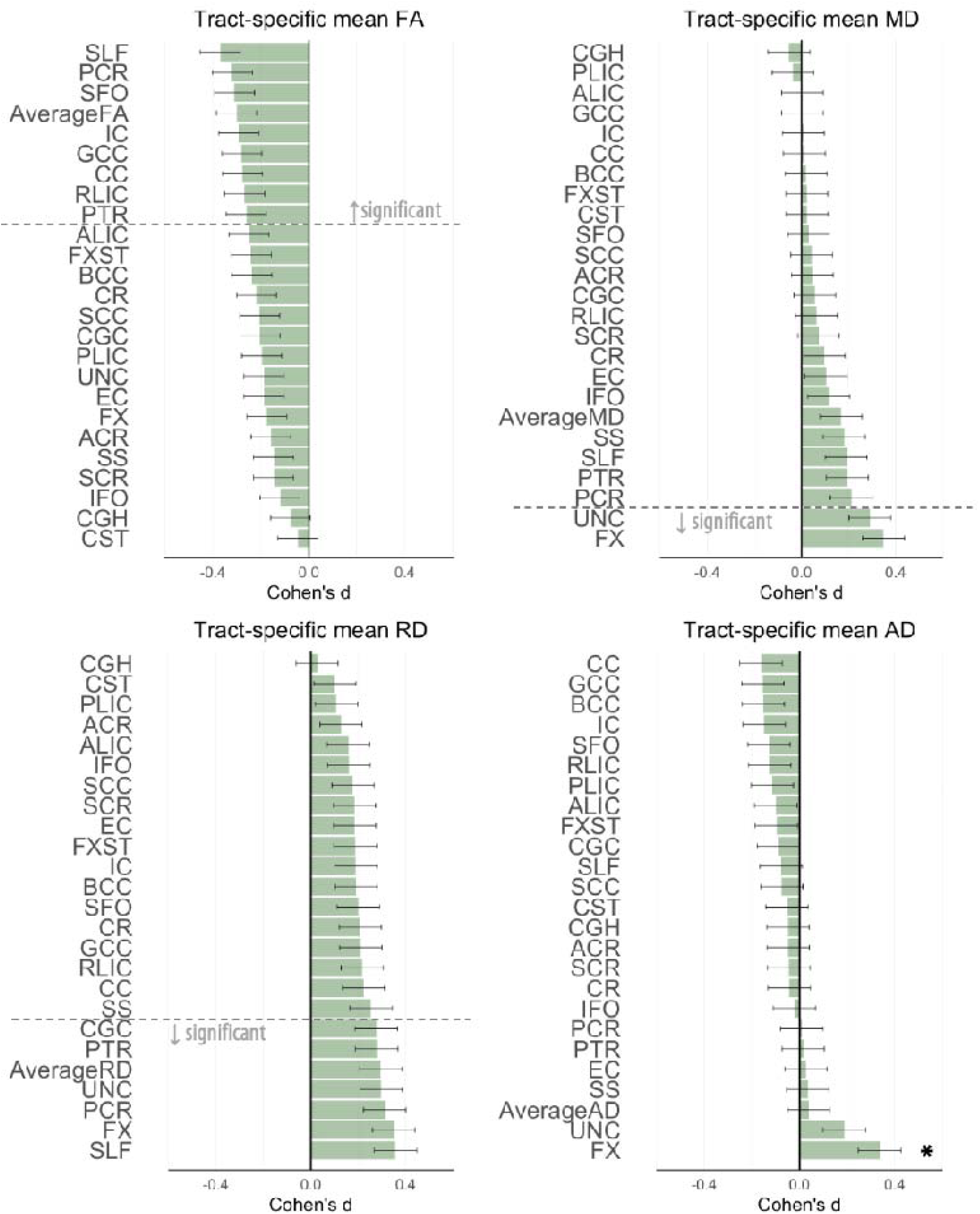
Cohen’s *d* values for differences in diffusion measures between adolescents with early-onset psychosis and healthy controls. Cohen’s *d* values and their standard errors are displayed, sorted by effect size. Stars and dashed lines indicate significant results (p ≤ 0.002). Abbreviation: FA = fractional anisotropy, MD = mean diffusivity, RD = radial diffusivity, AD = axial diffusivity. For white matter tract abbreviations, see Table 1.

### Controlling for average, core, and periphery diffusivity measures

Adjusting for average FA or core FA, none of the FA case-control differences remained significant after correction for multiple comparisons. Covarying for periphery FA, there was significantly lower FA in the SLF only (p ≤ 0.002, see Supplementary Table S7 and Figure S3-6). For MD, after covarying for average or periphery MD, the significance of case-control-differences in the FX remained. After additional adjustment for core MD, the case-control differences in the FX and UNC were still significant. For RD, no case-control differences remained significant after adjustment for average, core, or periphery RD. Lower AD in the FX remained significant after all additional adjustments.

### Diagnostic subgroup findings

When stratifying EOP by diagnostic subgroup, only adolescents with EOS showed significantly lower FA in 14 ROIs relative to CTR, including: Average FA, ALIC, BCC, CC, CR, FXST, GCC, IC, PCR, PTR, RLIC, SCC, SFO and SLF (p ≤ 0.002; see Figure 2 and Supplementary Table S10). MD and AD were only significantly higher in the FX of adolescents with EOS compared to CTR. We found higher RD in the CC, CGC, FX, PCR, PTR, RLIC, SLF, and SS in adolescents with EOS, and higher Average RD in adolescents with AFP, relative to CTR (p ≤ 0.002). We observed no significant white matter microstructural alterations in adolescents with OTP relative to CTR.

**Figure 2.**
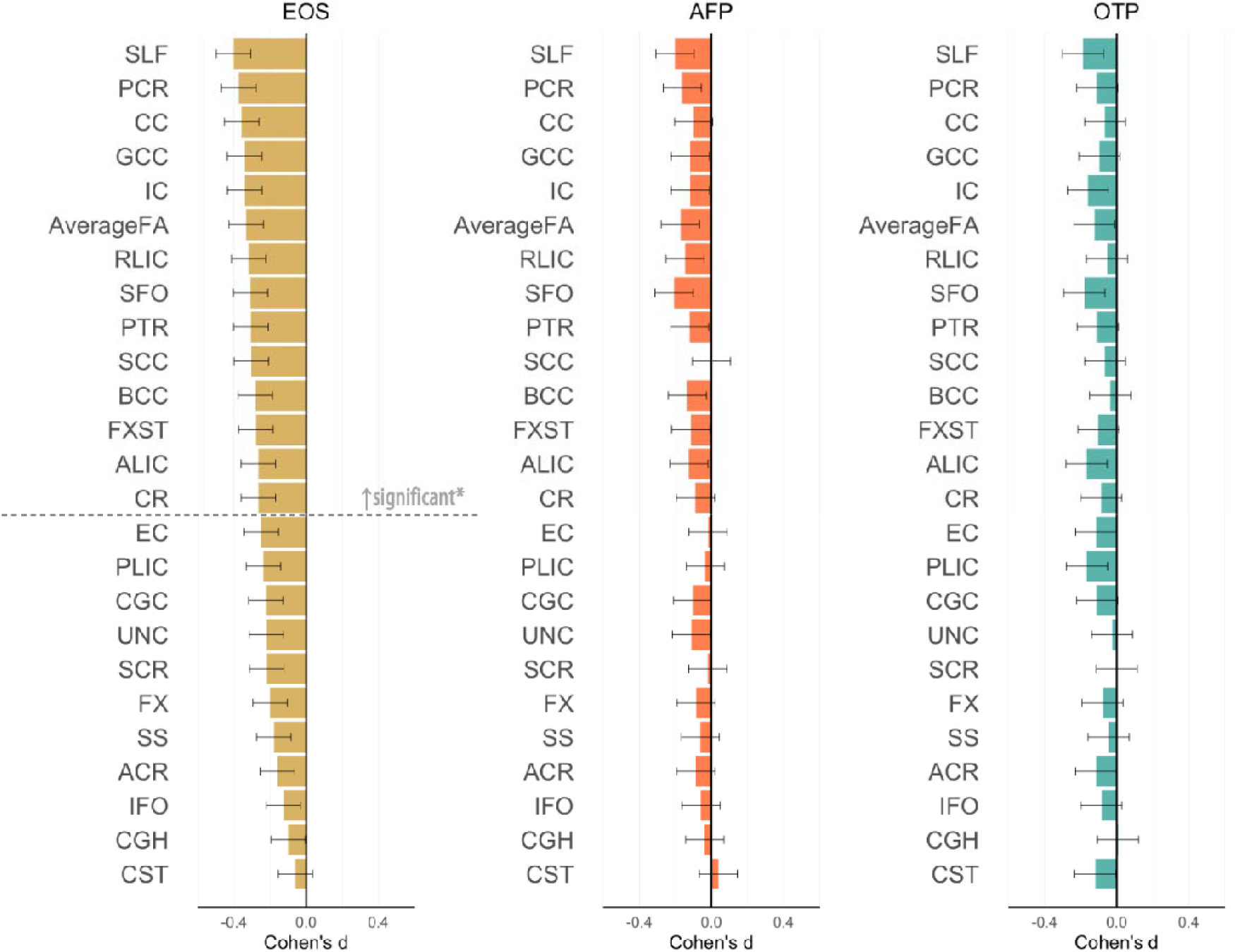
Fractional anisotropy (FA) differences between adolescents with early-onset psychosis and healthy controls, stratified by diagnostic subgroups. Cohen’s d values and their standard errors are displayed, sorted by effect size for EOS. Stars and dashed lines indicate significant results (p ≤ 0.002). Abbreviations: EOS = early-onset schizophrenia (n = 180), AFP = affective psychosis (n = 95), OTP = other psychosis (n = 46). For white matter tract abbreviations see Table 1.

### Sex-by-diagnosis and age-by-diagnosis interactions

To examine sex differences in relation to diagnosis, sex-by-diagnosis interactions were estimated. After Bonferroni correction, no significant interactions for FA, MD, and AD were found (Supplementary Table S11). We did find a significant sex-by-diagnosis interaction for RD in the SCC (p ≤ 0.002). Sex-stratified analyses showed that only male adolescents with EOP showed widespread lower FA relative to healthy male controls (EOP = 172, CTR = 112; p ≤ 0.002, 12 ROIs: Average FA, BCC, CC, CGC, EC, FX, FXST, GCC, PCR, SCC, SFO, and SLF; Figure 3, Supplementary Table S9). Similarly, tract-specific RD and MD were only higher in males with EOP relative to male controls (p ≤ 0.002, Males: EOP = 147, CTR = 98, *RD*: Average RD, CC, CGC, CR, FX, FXST, PCR, PTR, RLIC, SCC, SLF, SS, UNC; *MD*: PCR, UNC). Tract-specific AD was not significantly different in patients vs. CTR. In females, FA, MD, RD and AD did not differ significantly between EOP and CTR after correction for multiple comparisons (FA: EOP = 149, CTR = 153; non-FA: EOP = 131, CTR = 133). A sex-stratified diagnostic subgroup analysis showed that the male-specific effects were again driven by the EOS group. No significant age-by-diagnosis interactions were found for FA, MD, RD, and AD (Supplementary Table S12).

**Figure 3.**
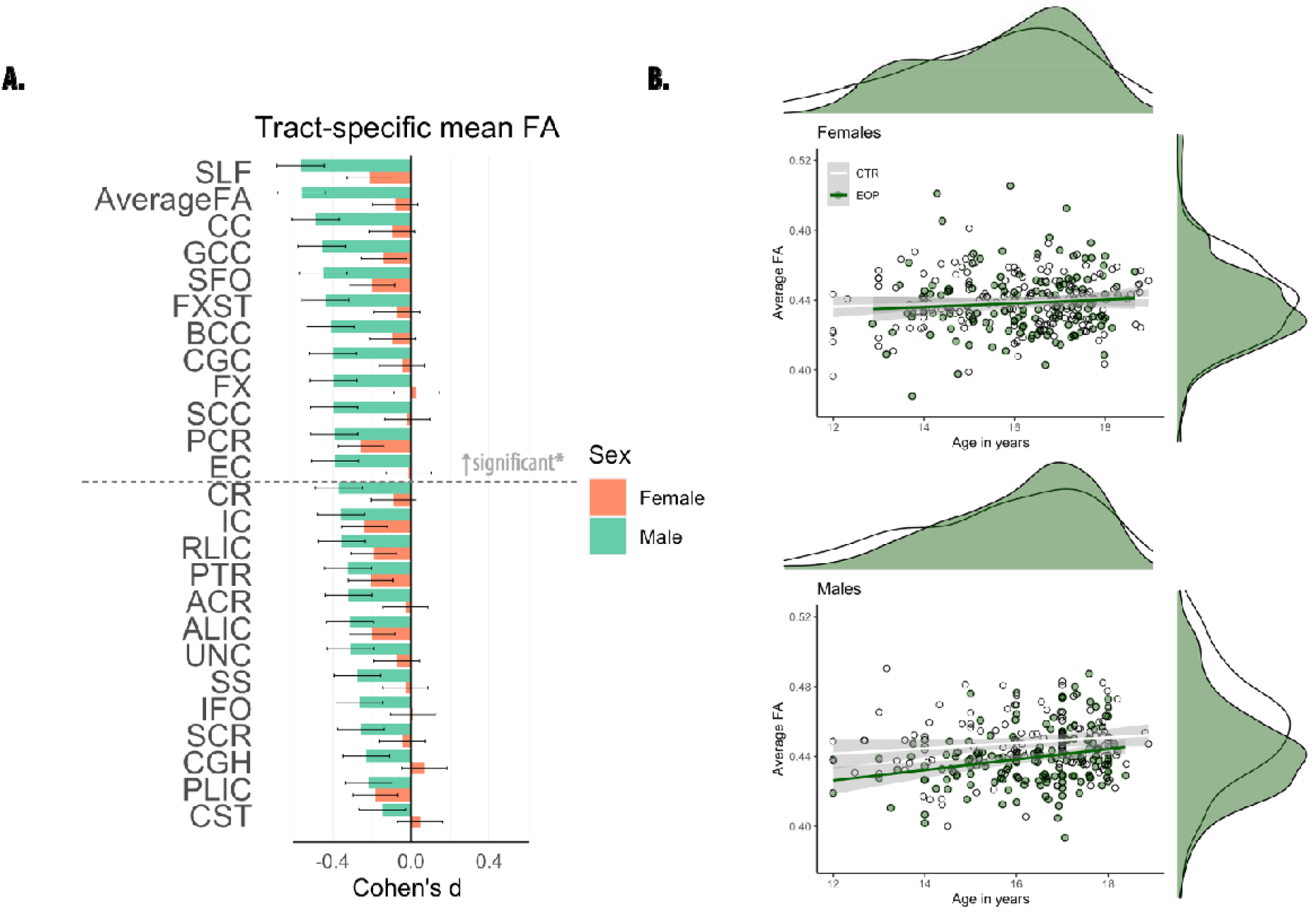
Fractional anisotropy (FA) differences between adolescents with early-onset psychosis and healthy controls, stratified by sex. A) Cohen’s *d* values and their standard errors are displayed, sorted by effect size for males. Stars and dashed lines indicate significant results (p ≤ 0.002) for males only*. In females, FA did not differ significantly between patients and healthy controls. For white matter tract abbreviations see Table 1. B) Marginal plots with distributions displaying average FA across the entire skeleton and age for females (upper panel) and males (lower panel) by diagnostic group. Diagnostic group-specific regression lines are shown.

### Associations with clinical measures in adolescents with EOP

After correction for multiple comparisons, we found no significant associations between current medication use, CPZ and tract-specific FA, MD, RD or AD in adolescents with EOP (whole sample: antipsychotics user = 256, non-user 29; Lithium user = 27, non-user = 247; antidepressants user = 57, non-user = 171; antiepileptics user = 14, non-user = 257, CPZ = 234; see Supplementary Table S11). Similarly, no DTI measures were significantly associated with symptom severity measures (n = 249, PANSS negative/positive). However, we did find a significant negative association between duration of illness and MD in the ALIC and the IC (p ≤ 0.002, see Supplementary Figure S8). Average RD and AD of BCC and CC were also negatively associated with duration of illness (p ≤ 0.002). Furthermore, AD of the ALIC was positively associated with age of onset (see Supplementary Figure S9).

### Comparison to adult patients with schizophrenia

Differences in the magnitude of tract-specific effect sizes between EOP and adult schizophrenia were observed, with effects being generally less pronounced in EOP (see Figure 4). We observed larger effect sizes for FA in EOP relative to adult schizophrenia only for the SLF and IC (including PLIC and RLIC), but these differences were not statistically significant (see Supplementary Table S16).

**Figure 4.**
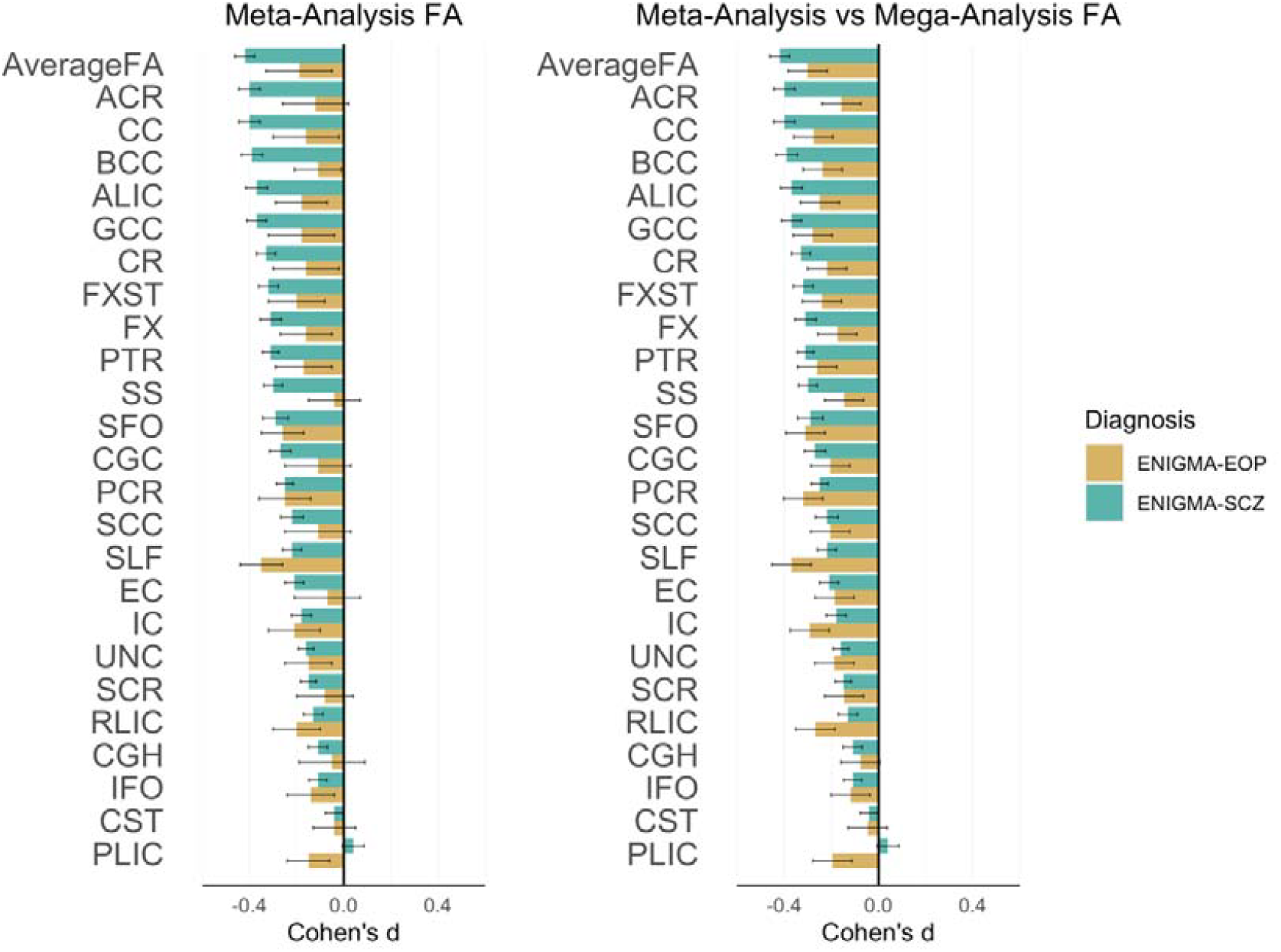
Effect sizes in early-onset psychosis (EOP) and adult schizophrenia (SCZ) relative to healthy controls from a prior publication^1^. In both EOP and SCZ, meta-analytic results are adjusted for age, sex, and linear and nonlinear age and sex interactions (age-by-sex interaction, age^2^, and age^2^-by-sex interaction). Cohen’s *d* values and their standard errors are displayed. SK provided values for SCZ.

## DISCUSSION

We found widespread lower FA in patients with EOP compared to CTR, with the largest effect sizes in the superior longitudinal fasciculus (SLF, *d* = 0.37), posterior corona radiata (PCR, *d* = 0.32), and superior fronto□occipital fasciculus (SFO, *d* = 0.31). Regions of the corpus callosum (CC), internal capsule (IC), and posterior thalamic radiation (PTR) also showed significant effects with Cohen’s *d* > 0.25. Lower FA in EOP was accompanied by widespread higher radial diffusivity (RD), and more localized higher mean diffusivity (MD) of the fornix (FX) and uncinate fasciculus (UNC). Higher axial diffusivity (AD) for EOP individuals was observed in the FX. Case-control differences in brain white matter microstructure were driven by individuals diagnosed with EOS, which represent the majority of the EOP sample.

### Regional specificity

The SLF is a major association tract connecting the parietal and temporal lobes with the frontal cortex, and has been implicated in working memory, attention, language, and emotion processing (37). Significant deficits in SLF white matter microstructure have previously been reported in youth with subsyndromal psychotic-like symptoms (38), clinical high-risk groups (39), EOP (22), and in recently diagnosed schizophrenia (40). These findings suggest that the SLF may play a role in the development of psychosis. Similarly, lower FA in the PCR and corpus callosum (CC) has also been reported in clinical high-risk populations (39, 41, 42). In addition, differences in white matter of the SFO, PCR, and CC have been associated with the transition to psychosis (43). Similar to other major white matter tracts, FA within the SLF increases significantly during adolescence (44). Interestingly, FA of the SLF showed a positive association with working memory performance in healthy individuals and patients in EOP (40), and may partially mediate increases in verbal fluency as a function of increasing age (44). Hence, FA deficits in the SLF may contribute to cognitive disturbances commonly reported in psychotic disorders.

A complementary meta-analysis of FA differences corroborated the significant effect of lower FA in the SLF. Yet, similar to Gurholt et al. (15), the meta-analysis suggests a great degree of heterogeneity across the included samples, likely reflecting differences in inclusion procedures (Table S2), imaging sequences (Table S3), and the inherent clinical diversity of EOP. Furthermore, not all samples from the mega-analysis could be included in the meta-analysis due to sample size limitations (n < 10), resulting in a higher variance between tracts and diagnostic groups in the meta-relative to the mega-analysis (see Figure 4).

In the present study, no regional FA differences remained significant after covarying for global effects, with the exception of lower FA in the SLF, thus suggesting that lower FA across the entire white matter skeleton is driving the difference in FA across almost all ROIs. Similar effects were observed for MD, RD, and AD. These findings reflect a pattern of globally lower DTI measures, commonly observed in adults with schizophrenia (1). While the signature of widespread FA deficits in EOP appears similar to that of adult schizophrenia, differences in the magnitude of tract-specific effect sizes between EOP and adult schizophrenia were observed, with effects being generally less pronounced and more variable in EOP (see Figure 4). In adult schizophrenia, lower FA was most prominent in the anterior corona radiata (*d* = 0.4) and CC (*d* = 0.39) (1). In EOP, effect sizes were largest for the SLF (*d* = 0.37), PCR (*d* = 0.32), and SFO (*d* = 0.31). Larger effect sizes for FA in EOP relative to adult schizophrenia were only found for the SLF and internal capsule (posterior/retrolenticular limb). Yet, using z tests, these differences in effect sizes between adolescent EOP and adult schizophrenia were not statistically significant, likely influenced by largely different sample sizes between studies (adolescent EOP, n = 586, adult schizophrenia, n = 4,322).

### Clinical and demographic correlates

The diagnostic subgroup analysis showed that widespread lower FA was limited to EOS (n = 180). No case-control differences in FA were found for AFP (n = 95) and OTP (n = 46). This finding is not in line with previous studies in adolescents (45, 46) and adult affective psychosis (47), reporting lower FA in different white matter tracts. Inconsistencies with previous results may be explained by the unbalanced sizes of the subgroups and differences in subgroup characteristics such as age of onset, disease duration, and psychotic features (Table 3). Larger samples are needed to stratify for clinical subgroups in relation to white matter structure.

No significant age-by-diagnosis interactions were observed. However, MD in the ALIC and IC were negatively associated with duration of illness. Similarly, average RD and AD in the BCC and CC were negatively associated with the duration of illness. Furthermore, AD in the ALIC was positively associated with age of onset. These findings suggest that the white matter differences in these regions may be linked to disease progression, as opposed to developmental factors. However, as EOS is associated with a longer duration of illness, these findings may also reflect an effect of the diagnostic subgroup.

We found no significant sex-by-diagnosis interaction for DTI measures, except for RD in the SCC. However, a sex-disaggregated analysis showed that male adolescents with EOP had widespread lower FA relative to male controls, whereas females with EOP did not significantly differ from female controls. Plotting average FA by sex and diagnostic group against age further highlighted consistently lower FA values in male patients compared to male controls across the studied age range (Figure 3, B). Average FA in female patients did not differ from female controls between the ages of 12 to 18 years. This finding suggests more pronounced white matter alterations in male EOP patients relative to same-sex controls and female patients. Sex differences in the developmental trajectory of white matter have been widely observed, with males typically showing protracted white matter maturation compared to females (20, 48, 49). As these differences may correspond to the impact of sex hormones on white matter during pubertal maturation (50), our findings may reflect the potential protective effect of estrogen for females against development and severity of psychosis (51). However, longitudinal studies are needed to establish whether white matter maturation differs between the sexes in EOP relative to healthy same sex controls.

There were no significant differences between medication users and non-users and impact of CPZ on white matter microstructure, which is similar to findings observed in adult schizophrenia (1). In addition, no significant associations between white matter and symptom severity were found, also in agreement with previous findings in EOP (2) and adult samples (1).

### Study limitations

This study is subject to some limitations. Firstly, the cross-sectional design of this study does not allow for a more thorough investigation of the effects of sex, duration of illness, and medication exposure. Secondly, FA is a summary measure of white matter microstructure that does not map perfectly onto the microstructural properties of the underlying tissue. The observed differences in FA may be influenced by a number of neurobiological processes, including changes in fiber organization such as packing density and axon branching, as well as alterations in myelination (16, 52, 53). In this study, we found that lower FA largely overlapped with higher RD, indicative of either demyelination or dysmyelination (16), with minimal changes to AD. However, inflammatory processes associated with psychosis onset can also impact DTI measures (54-56). Advanced imaging techniques, such as free-water imaging, separates the contribution of extracellular water from water diffusing along the axon to allow for improved specificity to detect microstructural differences (57, 58). When considering our findings, it is also important to account for the potential impact of ongoing white matter maturation in adolescent EOP, e.g., FA increases are typically reported throughout childhood and early adulthood. However, studies of white matter maturation in EOP have reported inconsistent findings, suggesting diverging, converging, or parallel developmental trajectories to healthy controls (2).

Finally, tract-based spatial statistics (TBSS) is commonly used to perform voxel-based analysis of white matter (59). However, the method is not without limitations. For example, spatial normalization can result in misalignment, with smaller tracts being particularly susceptible (60). In addition, smaller atlas ROIs such as the FX and CST are more vulnerable to partial volume effects and motion artifacts. Nevertheless, the ENIGMA-DTI Working Group has rigorously tested the reproducibility of measures using this TBSS approach for ROI analyses (61). Further, we combined neuroimaging datasets from nine sites, introducing heterogeneity due to different scanners, vendors, and sequences. In line with recommendations from the ENIGMA consortium, we addressed this issue by using the batch adjustment method, ComBat, which has been shown to reduce site-related heterogeneity and to, therefore increase statistical power (33). However, residual scanner effects may still be present. Despite these methodological limitations, the present study represents the most extensive analysis of white matter differences in EOP to date. The pooling of data worldwide and the implementation of harmonized protocols for image processing, quality control, and statistical analyses allow for greater statistical power and more robust estimates of effect size.

## Conclusion

In the largest analysis of white matter differences in EOP to date, we found widespread lower FA and higher RD with more localized differences in MD and AD for EOP relative to healthy controls. The largest effects were observed in the SLF and PCR, and in interhemispheric and thalamo-cortical regions. Differences were most pronounced in patients with EOS, and in male EOS patients relative to same-sex controls. The global pattern of widespread microstructural alterations observed in EOP further solidifies the hypothesis that schizophrenia may be a disorder of global brain structural connectivity. Future analyses of longitudinal data will allow for a more in-depth investigation of brain maturation in EOP and for further explorations of the effects of sex, duration of illness, and medication exposure.

## Data Availability

All data produced in the present study are available upon reasonable request to the authors.

## ACKNOWLEDGMENTS

**ENIGMA Core:** U.S. National Institutes of Health grants, R01 EB015611, U54 EB020403, R01 MH116147, R01 MH117601, R01 MH121246, S10 OD023696. **YTOP:** The Research Council of Norway (#223273, #2137000, #250358, #288083); South-Eastern Norway Regional Health Authority (#2017112); KG Jebsen Stiftelsen (#SKGJ-MED-008). **MADRID:** Celso Arango has received funding from CIBERSAM: Instituto de Salud Carlos III, Spanish Ministry of Science and Innovation, co-financed by ERDF Funds from the European Commission, “A way of making Europe”; CIBERSAM; Instituto de Investigación Sanitaria Gregorio Marañón; Madrid Regional Government (B2017/BMD-3740 AGES-CM-2, Youth Employment Operational Program and Youth Employment Initiative); European Social Fund and EU Structural Funds; EU Seventh Framework Program under grant agreements FP7-HEALTH-2009-2.2.1-2-241909, FP7-HEALTH-2009-2.2.1-3-242114, FP7-HEALTH-2013-2.2.1-2-602478, and FP7-HEALTH-2013-2.2.1-2-603196; EU H2020 Program under the Innovative Medicines Initiative 2 Joint Undertaking (grant agreement No 115916, Project PRISM, and grant agreement No 777394, Project AIMS-2-TRIALS), Fundación Familia Alonso, Fundación Alicia Koplowitz and Fundación Mutua Madrileña. Covadonga M. Díaz-Cenaj has received grant support from Instituto de Salud Carlos III (PI17/00481, PI20/00721, JR19/00024). **FEMS**: This project was supported by an unrestricted grant from AstraZeneca. Michael Berk is supported by a National Health and Medical Research Council (NHMRC) Senior Principal Research Fellowship (1059660 and APP1156072). **SCAPS**: The Swedish Research Council (#521-2014-3487, #2017-00949); FORMAS (#259-2012-31). **OXFORD:** MRC G0500092. MEND: NIMH R01 MH101506. **PAFIP:** This study was supported by grants from Carlos III Health Institute (IPI17/00402, PI17/01056, PI14/00639 and PI14/00918) cofunded by The European Union through FEDER funds and Fundación Instituto de Investigación Marqués de Valdecilla (NCT0235832 and NCT02534363). No pharmaceutical company has financially supported the study. **BARCELONA:** Elena de la Serna has received grant support from Instituto de Salud Carlos III (PI20/00654). Gisela Sugranyes has received research funds from Instituto de Salud Carlos III (PI1800976 and PI2100330), the Alicia Koplowitz Foundation (AKOPLOWITZ20_004), Ajut a la Recerca Pons Bartran (FCRB_PB1_2018) and a Brain and Behaviour Research Foundation NARSAD 2017 Young Investigator Grant (26731). **UCLA:** NIMH grants U01MH081902, P50MH066286.

We thank the research team and participants at each contributing site for the opportunity to conduct this research.

## CONFLICT OF INTEREST

For work unrelated to the contents of this manuscript, the following authors received funding from third-parties. **Celso Arango:** has been a consultant to or has received honoraria or grants from Acadia, Angelini, Gedeon Richter, Janssen Cilag, Lundbeck, Medscape, Otsuka, Roche, Sage, Servier, Shire, Schering Plough, Sumitomo Dainippon Pharma, Sunovion, and Takeda; **Michael Berk**: was supported by an unrestricted grant from AstraZeneca; **Covadonga M. Díaz-Caneja**: has received honoraria from Exeltis and Angelini; **Gisela Sugranyes** has received honoraria from Angelini; **Paul M. Thompson, Neda Jahanshad**: MPI of a research grant from Biogen, Inc.; **Ole A. Andreassen**: has received speaker’s honorarium from Lundbeck and is a consultant to HealthLytix.

## Notes

### Author Declarations

The ENIGMA-EOP Working Group obtained case-control data from nine cohorts across seven countries. The study was coordinated by Claudia Barth and Ingrid Agartz, who got approval for this work by the Norwegian Regional Committees for Medical and Health Research Ethics (REK, Number: 2009/691). For each site, all study participants and/or their legal guardians provided written informed consent with approval from local institutional review boards and the respective ethics committees.

