## Supplemental Material for "*In vivo* white matter microstructure in adolescents with early-onset psychosis: a multi-site mega-analysis via the ENIGMA Consortium"

**Overview**

1. **Notes**


**S1** ComBat Harmonization

**S2** Meta-analysis


2. **Figures**


**S1** Fractional anisotropy measures before and after ComBat harmonization using principal component analysis.

**S2** Mean, radial and axial diffusivity measures before and after ComBat harmonization using principal component analysis.

**S3** Cohen's d values for differences in fractional anisotropy (FA) between adolescents with early-onset psychosis and healthy controls, unadjusted as well as adjusted for core, periphery or average FA.

**S4** Cohen's d values for differences in mean diffusivity (MD) between adolescents with early-onset psychosis and healthy controls, unadjusted as well as adjusted for core, periphery or average MD.

**S5** Cohen's d values for differences in radial diffusivity (RD) between adolescents with early-onset psychosis and healthy controls, unadjusted as well as adjusted for core, periphery or average RD.

**S6** Cohen's d values for differences in axial diffusivity (AD) between adolescents with early-onset psychosis and healthy controls, unadjusted as well as adjusted for core, periphery or average AD.

**S7** Forest plots showing site-wise fractional anisotropy differences between early-onset psychosis patients and healthy adolescent controls – bilateral tracts.

**S8** Significant associations between duration of illness and diffusion measures in patients with early-onset psychosis.

**S9** Significant association between age of illness onset and mean axial diffusivity (AD) in the anterior limb of the internal capsule of patients with early-onset psychosis.


3. **Tables**


**S1** Site overview.

**S2** Site-wise inclusion and exclusion criteria.

**S3** Diffusion weighted imaging acquisitions parameters, stratified by site and scanner.

**S4** Demographic and clinical characteristics, stratified by site.

**S5** Demographic and clinical characteristics, stratified by sex.

**S6** Multiple linear regression output for case-control differences in bilateral regional diffusion measures.

**S7** Multiple linear regression output for case-control differences in bilateral regional mean diffusion metrics, adjusted for average, core or periphery diffusion measures.

**S8** Multiple linear regression output for case-control differences in lateralized regional diffusion measures.

**S9** Multiple linear regression output for case-control differences in bilateral regional diffusion measures, stratified by sex.

**S10** Multiple linear regression output for diagnostic subgroup differences relative to healthy controls in bilateral regional diffusion measures.

**S11** Multiple linear regression output for sex-by-diagnostic group interactions in bilateral regional diffusion measures.

**S12** Multiple linear regression output for age-by-diagnostic group interactions in bilateral regional diffusion measures.

**S13** Multiple linear regression output for association between medication use and bilateral regional diffusion measures, in patients with early-onset psychosis.

**S14** Multiple linear regression output for association between clinical measures and bilateral regional diffusion measures, in patients with early-onset psychosis.

**S15** Meta-analytic results for fractional anisotropy differences between adolescents with early-onset psychosis and healthy controls.

**S16** Direct comparison of meta- and mega-analytically derived effect sizes for case-control FA differences between early-onset psychosis (EOP) and adult schizophrenia (SCZ)

#### 1. Notes

##### *S1: ComBat - Harmonization*

ComBat is a batch-effect correction tool from the genomics literature, which has become increasingly popular as a harmonization procedure for multi-site neuroimaging data. It employs a Bayesian framework to estimate the additive and multiplicative effects of site, which are used to adjust for scanner-related effects. To assess the effectiveness of ComBat harmonization in our study, we performed principal component analysis (PCA) of DTI measures before and after ComBat harmonization and created scatterplots (Figure S1-S2) where the first principal component (PC1) was plotted against the second principal component (PC2). Data from each site was marked with a unique color, and their geometric means were denoted with a crossed circle. Figures S1-S2 demonstrate how the large differences in geometric means between sites were drastically reduced after ComBat harmonization, indicating that the procedure had successfully reduced site differences.

##### *S2: Meta-analysis*

A complementary random-effects inverse-variance weighted meta-analysis for the main model investigating case-control FA differences was conducted in R (metafor package) to investigate the heterogeneity between sites. In line with Kelly et al. 2018 (1), all sites with a minimum of 10 participants per diagnostic group were included. We followed a similar procedure as described in Gurholt et al. 2020 (2). Site-wise linear regression using the lm function was conducted for each structure with diagnosis (i.e., patient-control status) as the variable of interest, while adjusting for age, sex, and linear and nonlinear age and sex interactions (age-by-sex interaction, $age^2$ and $age^2$-by-sex interaction) as covariates. The Cohen's d effect sizes of case-control differences and their standard errors were computed for each site (3).

For each white matter tract, we pooled the site-wise Cohen's d effect sizes and standard errors using an inverse variance-weighted random-effects model fitted by a restricted maximum-likelihood estimator, through the rma-function/metafor R package (version 2.0.0, (4)). This yields an estimate of the cross-site Cohen's d effect size, standard error, and 95% confidence interval of the effect size, as well as the z- and p-values. Forest plots were generated to illustrate the variability across sites, using the forest function from the metafor package.

#### 2. Figures

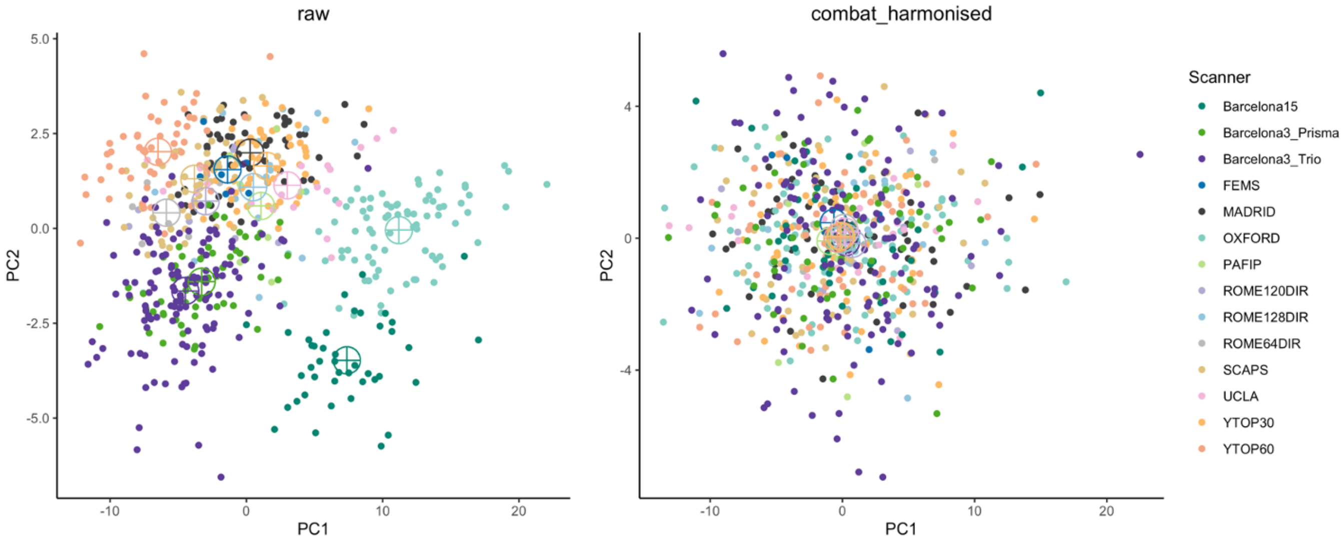


**Figure S1| Fractional anisotropy measures before and after ComBat harmonization using principal component analysis.** Abbreviations: PC = principal component.

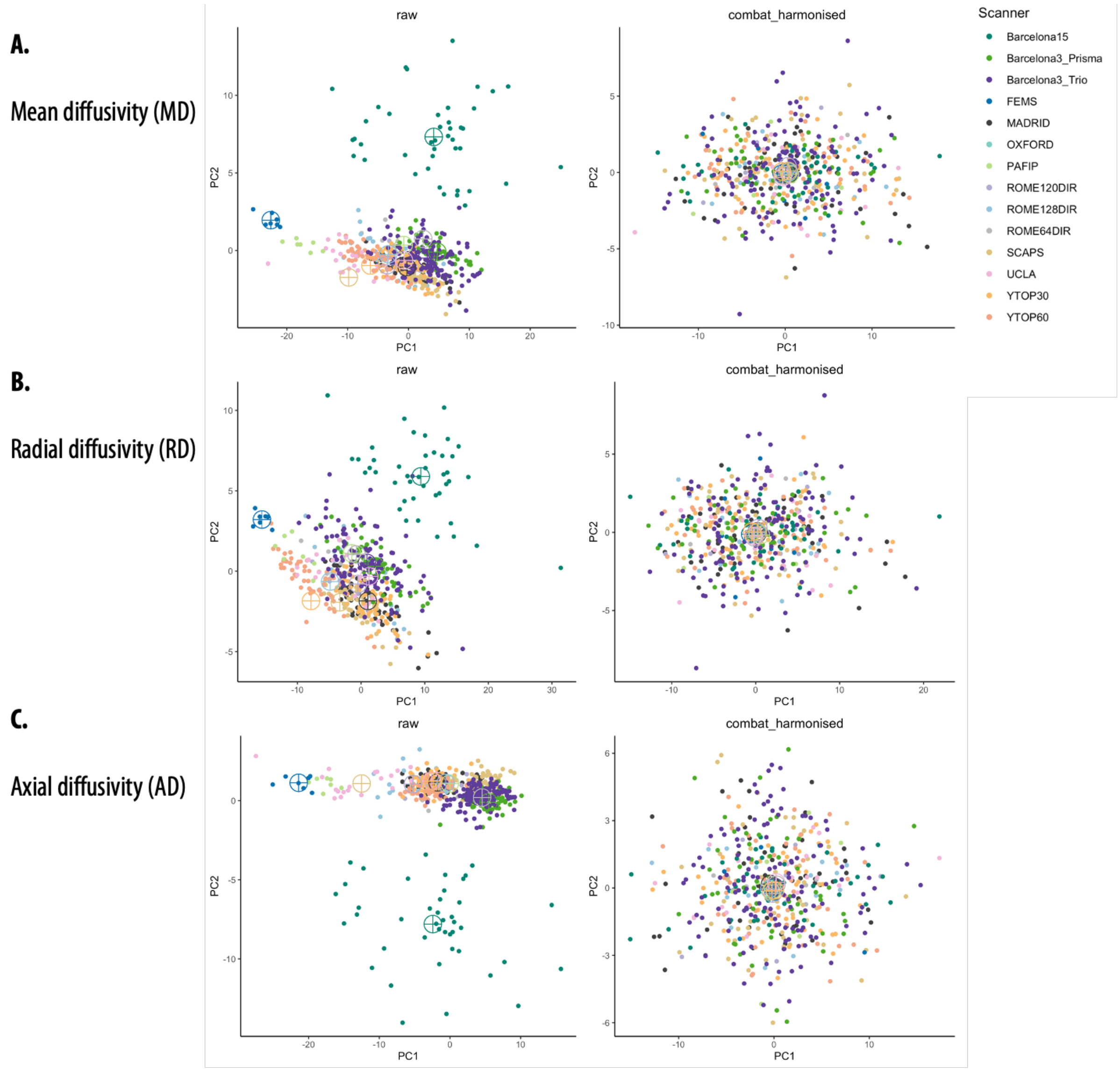


**Figure S2| Mean, radial, and axial diffusivity measures before and after ComBat harmonization using principal component analysis.**

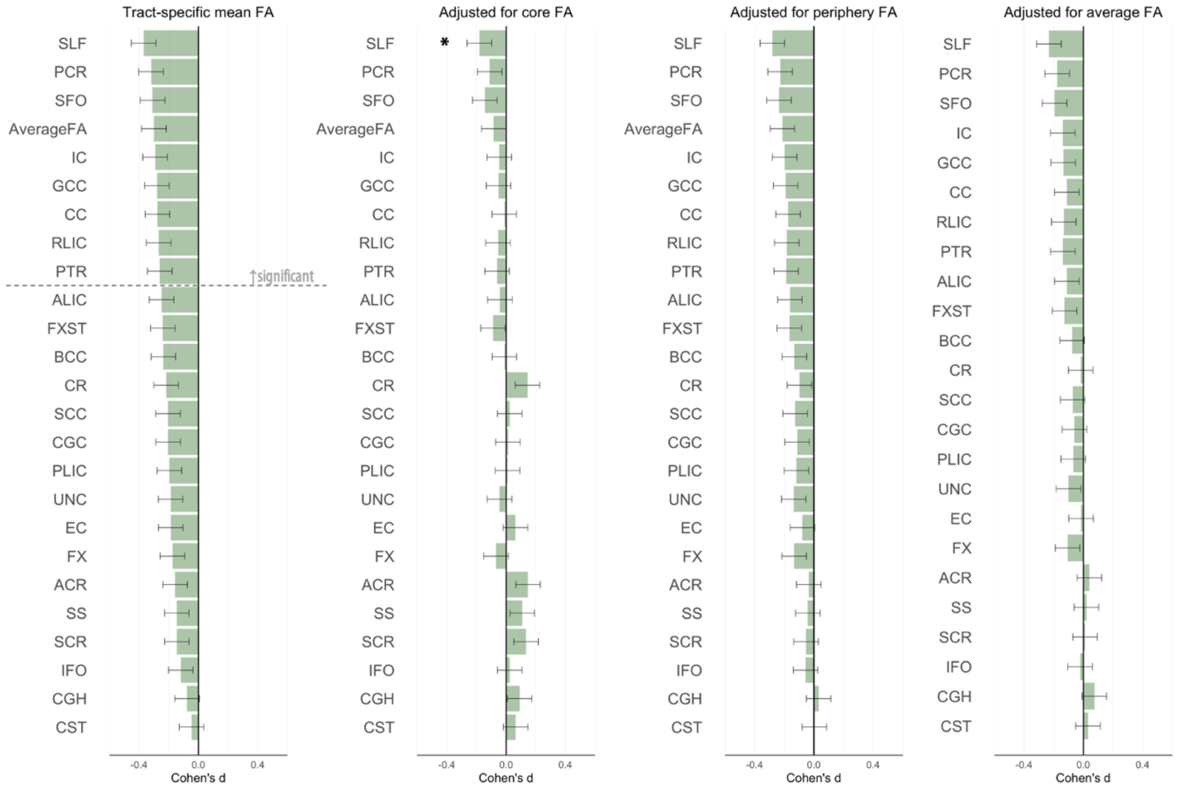


**Figure S3| Cohen's d values for differences in fractional anisotropy (FA) between adolescents with early-onset psychosis and healthy controls, unadjusted as well as adjusted for core, periphery, or average FA.** Cohen's d values and their standard errors are displayed, sorted in increasing magnitude of effect. Stars and dashed lines indicate significant results ($p \leq 0.002$). For white matter tract abbreviations, see Table 1, main text.

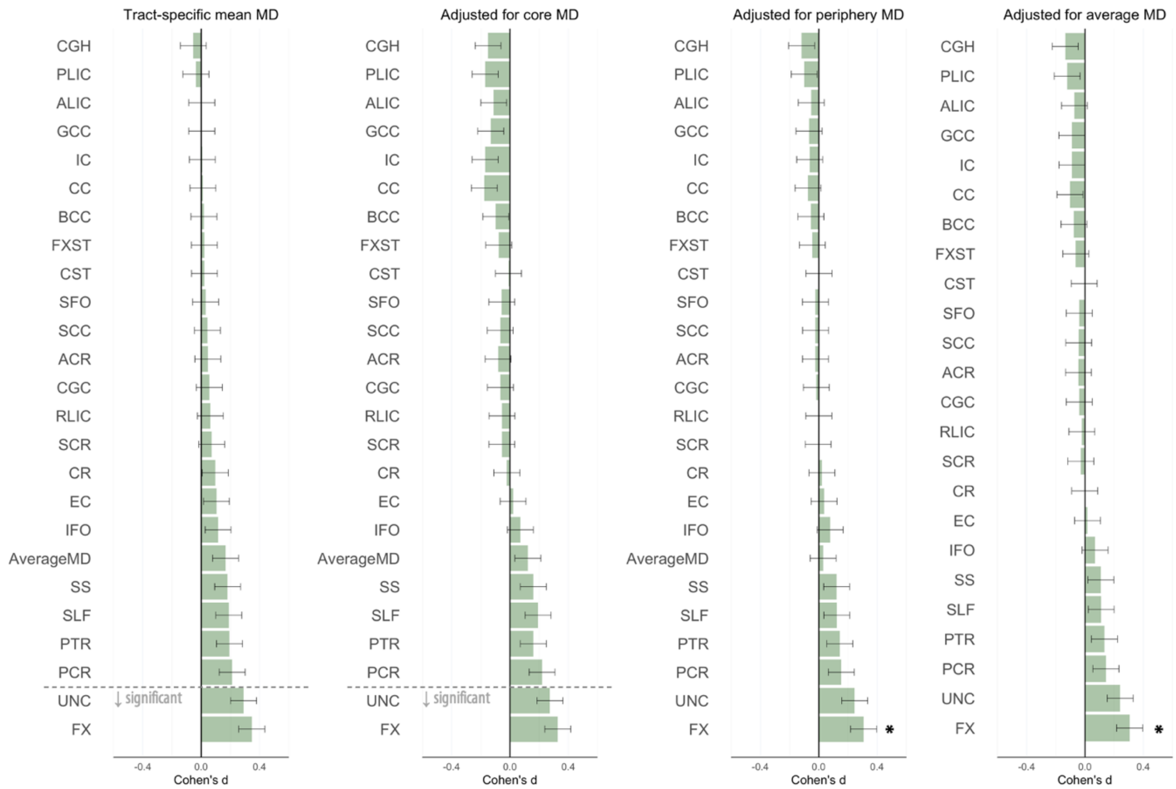


**Figure S4| Cohen's d values for differences in mean diffusivity (MD) between adolescents with early-onset psychosis and healthy controls, unadjusted as well as adjusted for core, periphery or average MD.** Cohen's d values and their standard errors are displayed, sorted in increasing magnitude of effect. Stars and dashed lines indicate significant results ($p \leq 0.002$). For white matter tract abbreviations, see Table 1, main text.

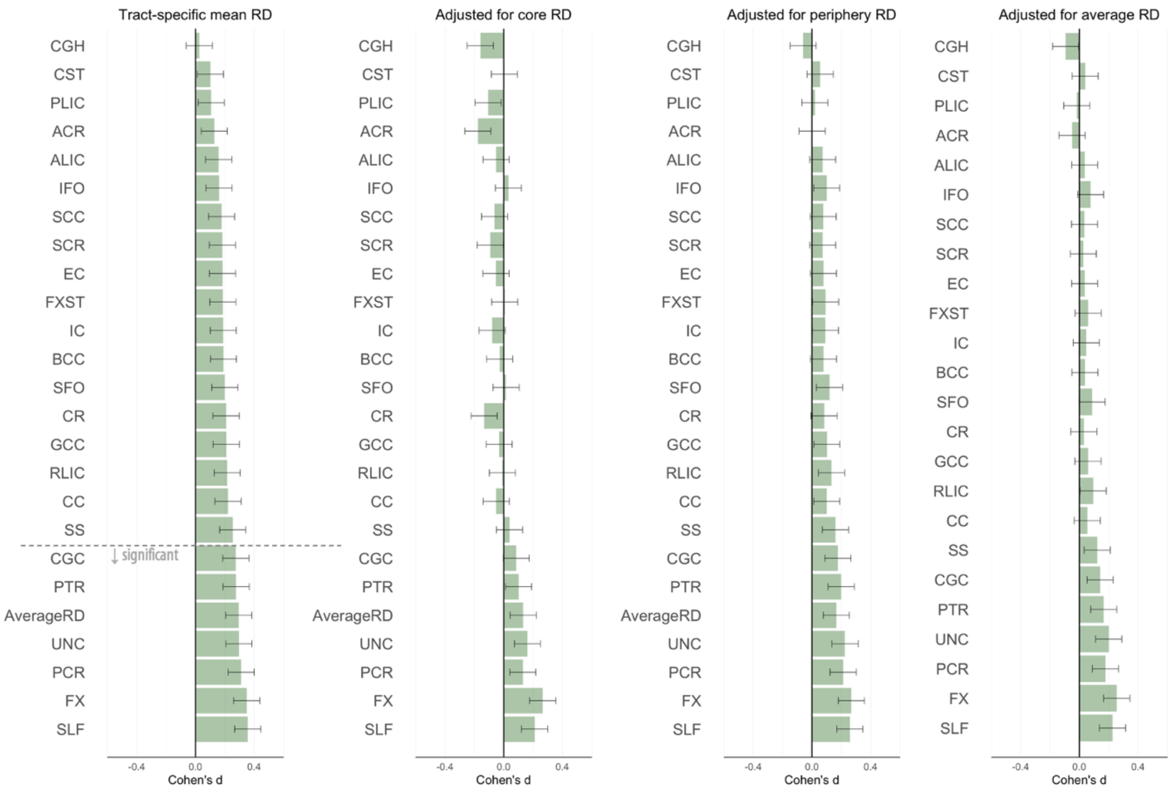


**Figure S5| Cohen's d values for differences in radial diffusivity (RD) between adolescents with early-onset psychosis and healthy controls, unadjusted as well as adjusted for core, periphery, or average RD.** Cohen's d values and their standard errors are displayed, sorted in increasing magnitude of effect. Dashed line indicates significant results ($p \leq 0.002$). For white matter tract abbreviations, see Table 1, main text.

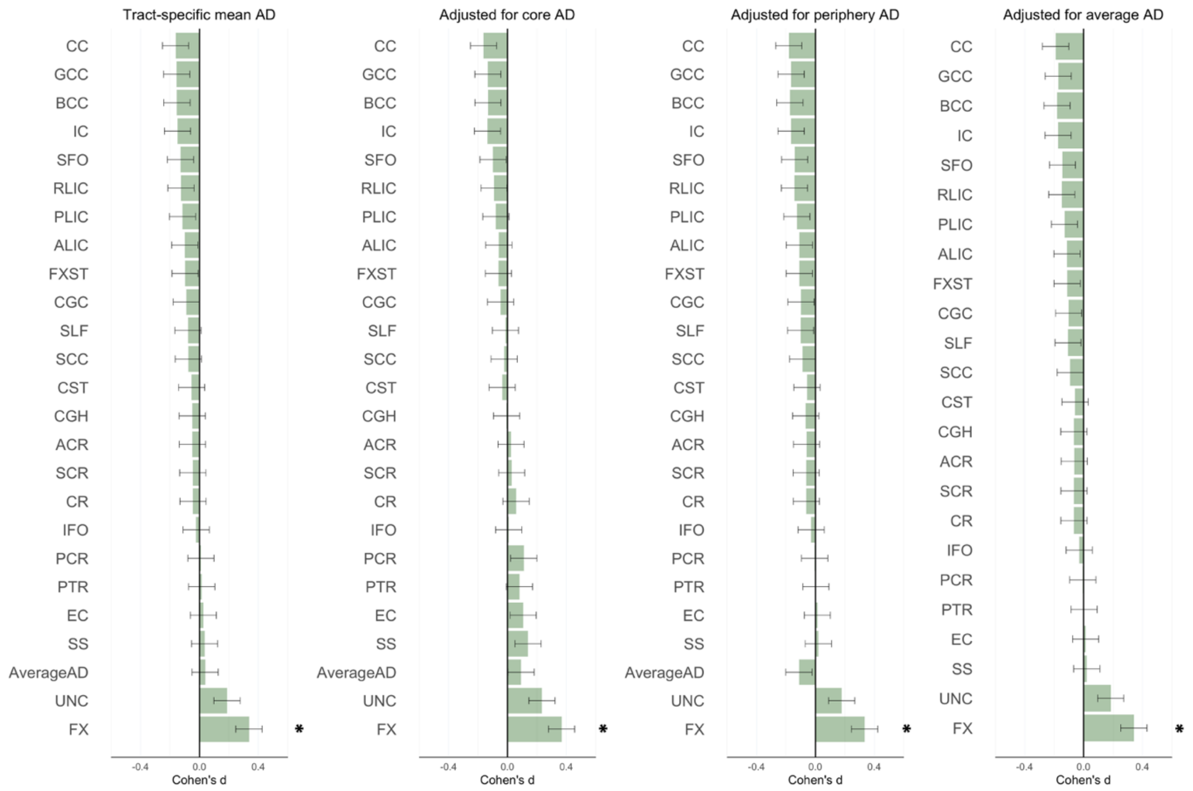


**Figure S6| Cohen's d values for differences in axial diffusivity (AD) between adolescents with early-onset psychosis and healthy controls, unadjusted as well as adjusted for core, periphery, or average AD.** Cohen's d values and their standard errors are displayed, sorted in increasing magnitude of effect. Stars indicate significant results ($p \leq 0.002$). For white matter tract abbreviations, see Table 1, main text.

### Supplemental Material

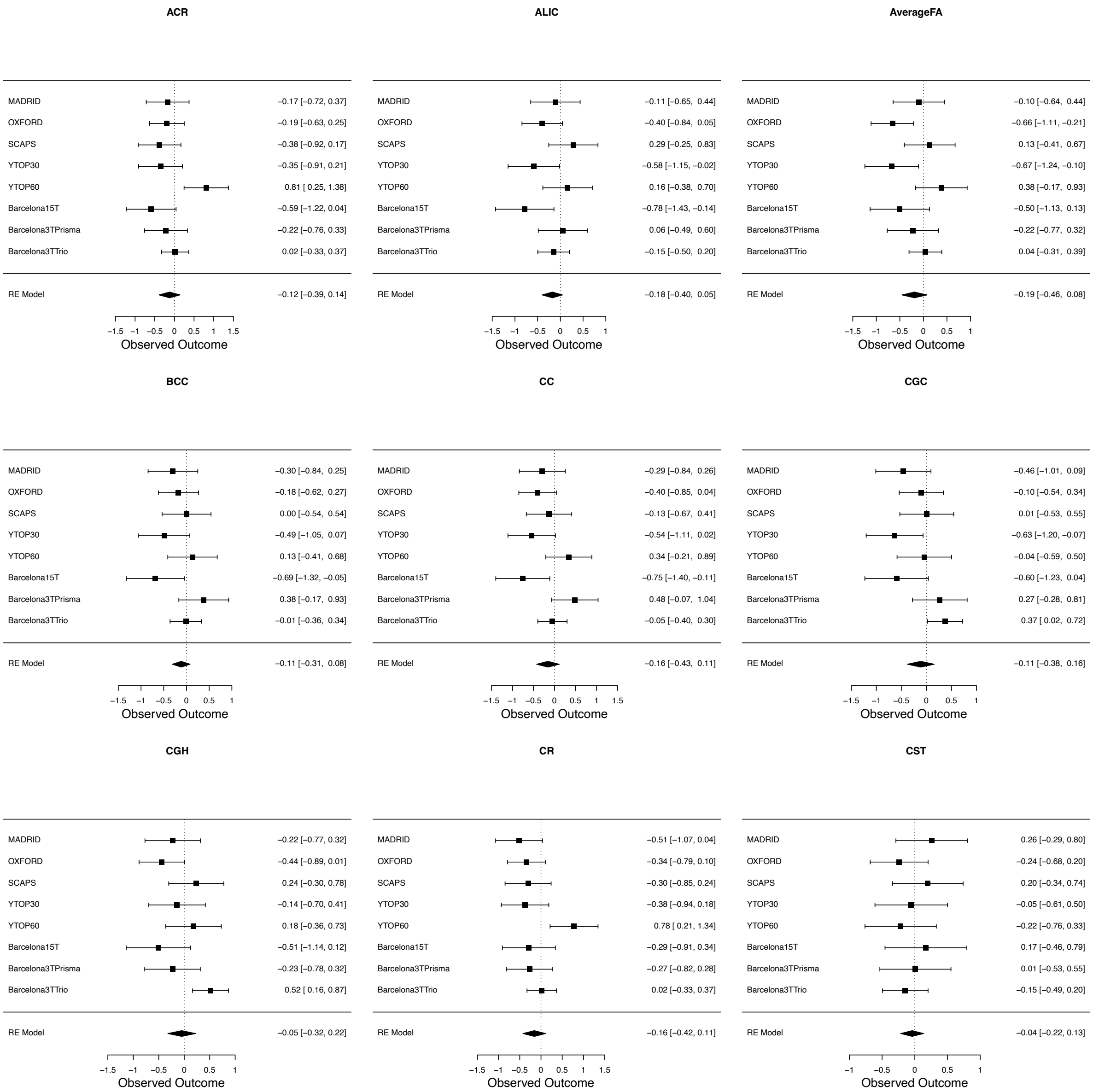

### Supplemental Material

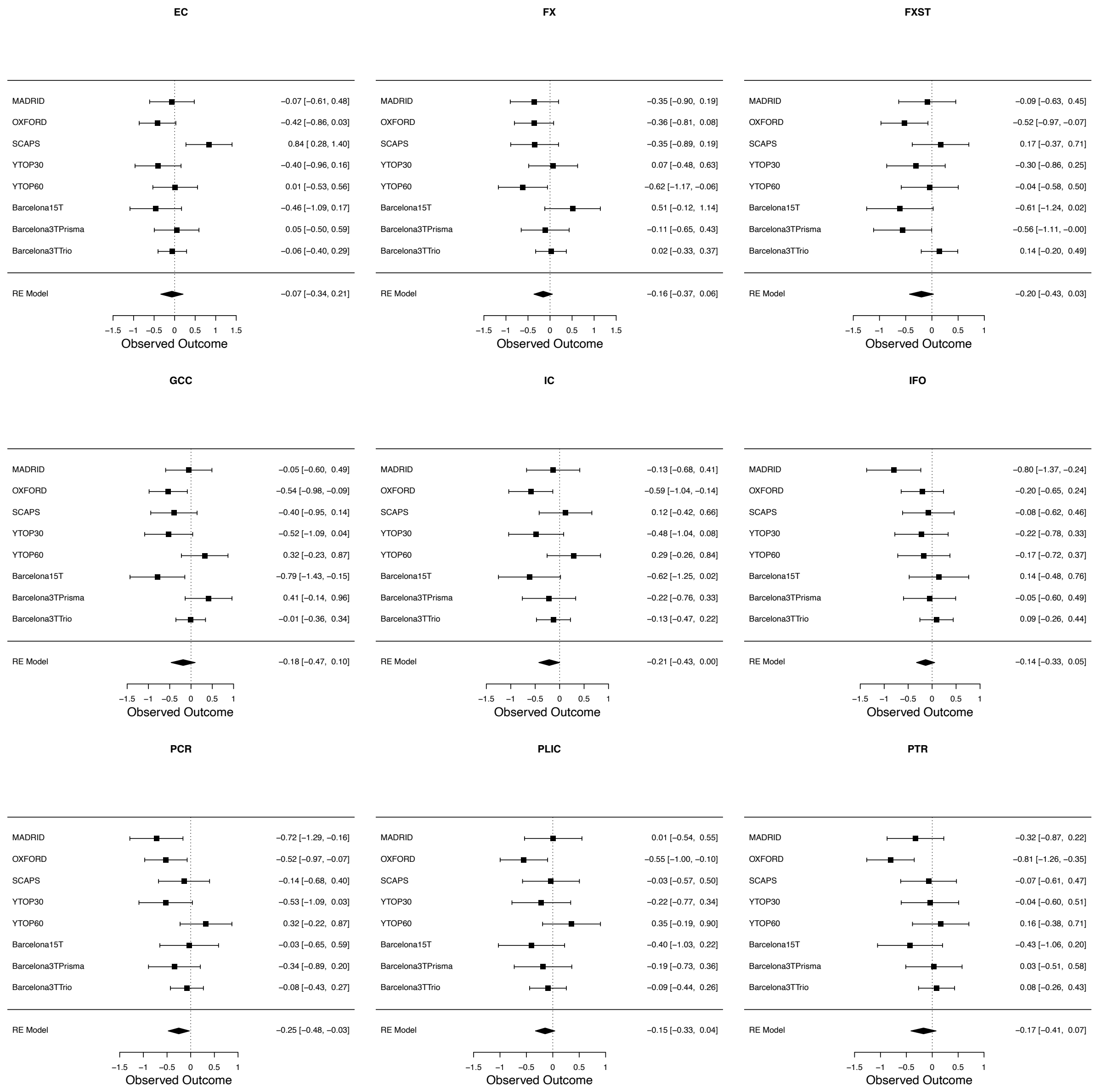

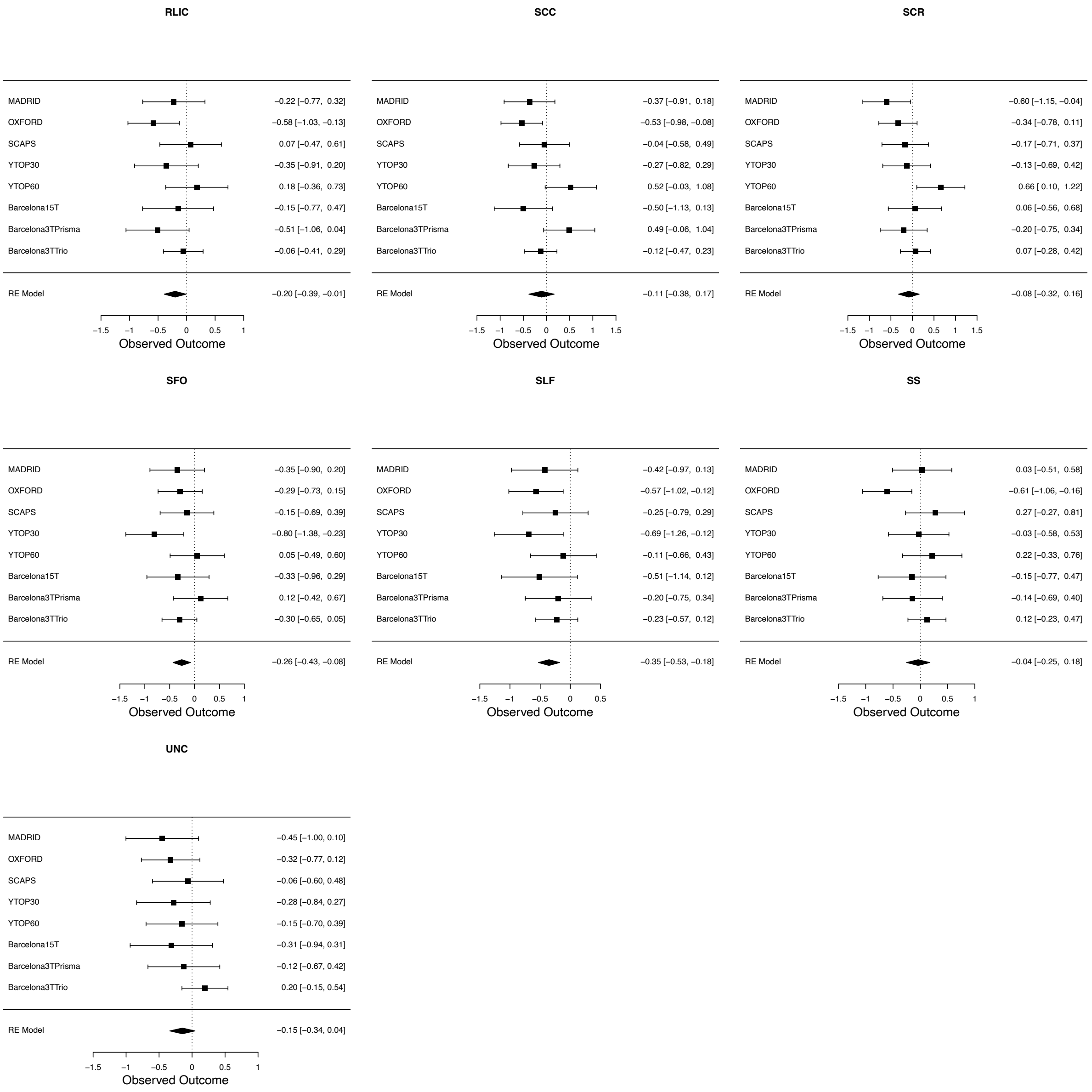


**Figure S7| Forest plots showing site-wise fractional anisotropy differences between early-onset psychosis patients and healthy adolescent controls – bilateral tracts.** Output is adjusted for age, sex, and linear and nonlinear age and sex interactions (age-by-sex interaction, age$^2$, and age$^2$-by-sex interaction). Cohen's *d* effect size and 95% confidence interval are shown for each cohort. The diamond indicates the pooled effect size across cohorts.

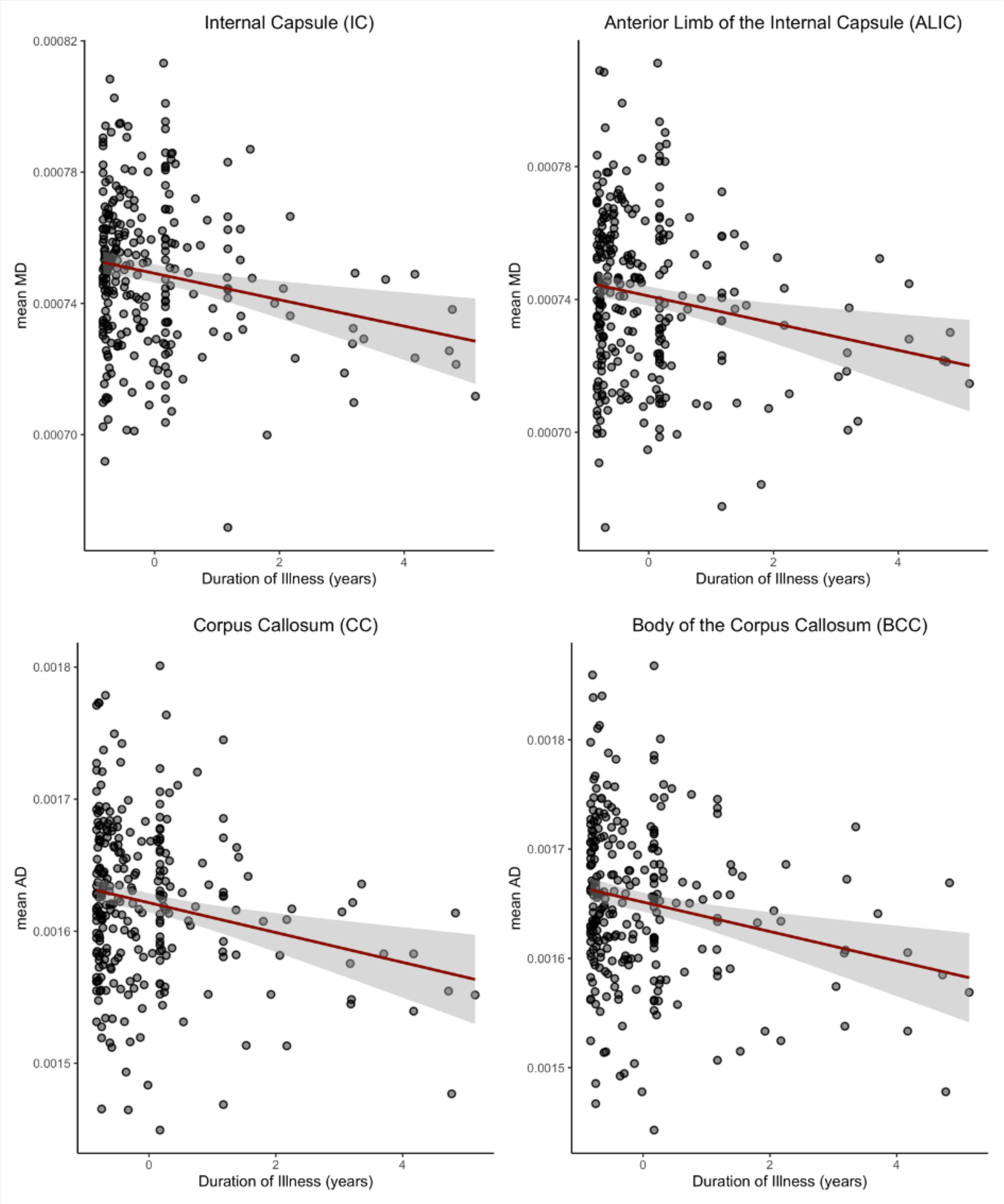


**Figure S8| Significant associations between duration of illness and diffusion measures in patients with early-onset psychosis.** Raw values with regression line and standard error are displayed. Abbreviation: MD = mean diffusivity, AD = axial diffusivity.

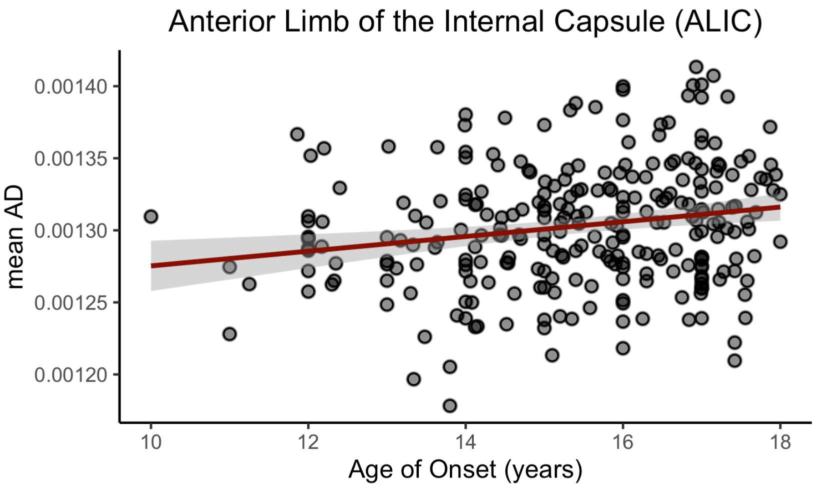


**Figure S9| Significant association between age of illness onset and mean axial diffusivity (AD) in the anterior limb of the internal capsule of patients with early-onset psychosis.** Raw values with regression line and standard error are displayed.

#### 3. Tables

**Table S1| Site overview.**

| Cohort | PI | Institution* | City | Country | Years of Data Inclusion |
|---|---|---|---|---|---|
| BARCELONA | Inmaculada Baeza | University of Barcelona | Barcelona | Spain | 2003 - 2020 |
| FEMS | Michael Berk | Deakin University | Geelong | Australia | 2009 - 2013 |
| MADRID | Celso Arango | Hospital General Universitario Gregorio Marañón | Madrid | Spain | 2006 - 2014 |
| OXFORD | Anthony James | University of Oxford | Oxford | UK | 2005 - 2009 |
| PAFIP | Benedicto Crespo-Facorro | University of Sevilla | Sevilla | Spain | 2008 - 2012 |
| ROME | Gianfranco Spalletta | IRCCS Santa Lucia Foundation | Rome | Italy | 2012 - present |
| SCAPS | Ingrid Agartz, Mathias Lundberg | Karolinska Institutet | Stockholm | Sweden | 2012 - 2019 |
| UCLA | Carrie E. Bearden | University of California | Los Angeles | USA | 2011- 2015 |
| YTOP | Ingrid Agartz | University of Oslo | Oslo | Norway | 2012 - present |

* More detailed affiliations can be found in the affiliation section

**Table S2| Site-wise inclusion and exclusion criteria.**

| Site | Diagnosis | Diagnostic tool | Recruitment information | Inclusion criteria | Exclusion criteria |
|---|---|---|---|---|---|
| BARCELONA | EOP | K-SADS/ DSM-IV-TR | Referral from inpatient or outpatient units of the Department of Child and Adolescent Psychiatry and Psychology of the Hospital Clinic Barcelona | Age between 7-18 years; onset of first psychotic positive symptom within a psychotic episode before age of 18; diagnosis of a psychotic disorder per DSM-IV-TR criteria; written informed consent | Intellectual disability per DSM-IV-TR criteria (IQ < 70 & impaired functioning); pervasive developmental disorder; past history of head trauma with loss of consciousness; pregnancy |
| | CTR | K-SADS/ DSM-IV-TR | Local catchment area via advertisements | Age between 7-18 years; written informed consent | Past history of psychotic illness; current diagnosis of any Axis-I DSM-IV-TR disorder; intellectual disability per DSM-IV-TR criteria (IQ < 70 & impaired functioning); past history of head trauma with loss of consciousness; pregnancy |
| FEMS* | EOP | DSM-IV | Referral from early psychosis services within the Geelong and the Southern Health sites | Psychotic disorders included: bipolar I disorder with psychotic features. schizoaffective disorder; age between 15-25 years; written informed consent; not have had a previous treated manic episode; quetiapine and lithium therapy for at least 1 month prior to randomization | Clinically relevant systemic disorder; pregnancy; sensitivity/allergy to quetiapine/lithium or their compounds; non-fluency in English; history of epilepsy; clinically relevant biochemical or hematological abnormalities; immediate risk of self-harm or risk to others; organic mental disease; IQ < 70; uncontrolled Diabetes Mellitus; use of cytochrome P450 3A4 inhibitors and/or cytochrome P450 inducers 14 days before enrollment; absolute neutrophil count of $1.5x10^9$ per liter |
| | CTR | NA | Local catchment area via advertisements; among | Matched to patients; age between 15-25 years; no | Clinically relevant systemic disorder; pregnancy; non-fluency |

| | | | | | |
|---|---|---|---|---|---|
| | | | hospital visitors (friends of patients); via Melbourne Neuropsychiatry. Parkville. Melbourne | history of mental illness; written informed consent | in English; history of epilepsy; clinically relevant biochemical or hematological abnormalities; organic mental disease; IQ < 70; uncontrolled Diabetes Mellitus; use of cytochrome P450 3A4 inhibitors and/or cytochrome P450 inducers 14 days before enrollment; absolute neutrophil count of $1.5x10^9$ per liter |
| MADRID* | EOP | K-SADS/ DSM-IV-TR | Referral from adolescent inpatient unit (Hospital General Universitario Gregorio Marañón) or local clinical services (PIENSA program) | Age between 7-18 years; onset of first psychotic positive symptom within a psychotic episode before age of 18; diagnosis of a psychotic disorder per DSM-IV-TR criteria; written informed consent | Intellectual disability per DSM-IV-TR criteria (IQ < 70 & impaired functioning); pervasive developmental disorder; past history of head trauma with loss of consciousness; pregnancy |
| | CTR | K-SADS/ DSM-IV-TR | Local catchment area via advertisements | Age between 7-18 years; written informed consent | Past history of psychotic illness; current diagnosis of any Axis-I DSM-IV-TR disorder; intellectual disability per DSM-IV-TR criteria (IQ < 70 & impaired functioning); past history of head trauma with loss of consciousness; pregnancy |
| OXFORD* | EOP | K-SADS-PL/DSM-IV | Local adolescent psychiatric units | Psychotic disorders included: schizophrenia | Moderate mental impairment; a history of substance abuse or pervasive developmental disorder; significant head injury; neurological disorder or major medical disorder |
| | CTR | KSADS-PL | Local general practitioners' practice | Healthy adolescents | Any medical/emotional/behavioral disorders; moderate mental impairment; a history of substance abuse or pervasive developmental disorder; significant head injury; |

| | | | | | |
|---|---|---|---|---|---|
| | | | | | neurological disorder or major medical disorder |
| PAFIP | EOP | DSM-IV | Local adolescent psychiatric units or local clinical services | SCID Axis I diagnosis confirmed by an independent psychiatrist 6 months after the initial contact; written informed consent | DSM-IV criteria for (1) drug dependence (except nicotine dependence), (2) mental retardation, and when having a history of neurological disease or head injury. |
| | CTR | Comprehensive Assessment of Symptoms and History | | Matched to patients (age, sex, laterality index, drug history, years of education); written informed consent | Current or past history of psychiatric, neurological or general medical illnesses, including substance dependence and significant loss of consciousness; presence of psychosis in first-degree relatives |
| ROME | EOP | DSM-V psychiatric and personality disorders using the SCID-5-RV and SCID-5-PD | Local catchment area and referral from adolescent psychiatric units | Age between 10-18 years; onset of first psychotic positive symptom within a psychotic episode before age of 18; suitability for MRI scanning; written informed consent | history of alcohol or drug abuse in the two years before the assessment; lifetime drug dependence; traumatic head injury with loss of consciousness; past or present major medical illness or neurological disorders; intellectual disability; pervasive developmental disorder |
| | CTR | DSM-V psychiatric and personality disorders using the SCID-5-RV and SCID-5-PD | Local catchment area via advertisements | Matched to patients; age between 10-18 years; suitability for MRI scanning; written informed consent | history of alcohol or drug abuse in the two years before the assessment; lifetime drug dependence; traumatic head injury with loss of consciousness; past or present major medical illness or neurological disorders; any psychiatric disorder or intellectual disability |
| SCAPS* | EOP | DSM-IV | Specialist care unit of psychosis and bipolar disorder in the | Psychotic disorders included: schizophrenia. schizoaffective disorder. | Substance-induced psychotic disorder; IQ < 70; previous |

| | | | | | |
|---|---|---|---|---|---|
| | | | department of Child and Adolescent Psychiatry in Stockholm. Sweden | psychotic depression; unspecified psychosis; bipolar I and II disorder; age between 12-18 years | moderate to severe head injury; organic brain disease |
| | CTR | NA | Invitation by letter after random draw from the Swedish National Registry | Age between 12-18 years. good command of the Swedish language to complete interview and neurocognitive tests | History of mental health issues (contact with specialist services); previous or current use of psychotropic medication; first-degree relatives with a history of psychotic disorders; IQ < 70; previous moderate to severe head injury; organic brain disease |
| UCLA* | EOP | DSM-IV. SCID Axis I Disorders | In-and outpatient clinics for child and adolescent mental health in the Greater Los Angeles area/website/local advertisements | Psychotic disorders included: schizophrenia spectrum disorder; age between 12-18 years; informed consent; no MRI contra-indications | Substance-induced psychotic disorder; IQ < 70; previous moderate to severe head injury; significant comorbid medial/neurological condition and/or history of head trauma with loss of consciousness |
| | CTR | DSM-IV. SCID Axis I Disorders | Local advertisements (online/brochures) in the Los Angeles areas | Matched to patients; age between 12-18 years; no history of major mental disorders; informed consent; no MRI contra-indications | History of mental health issues (contact with specialist services); first-degree relatives with a history of psychotic disorders; IQ < 70; significant comorbid medial/neurological condition and/or history of head trauma with loss of consciousness |
| YTOP* | EOP | K-SADS-PL (2009)/ DSM-IV | In-and outpatient clinics for child and adolescent mental health in the greater Oslo area | Age between 12-18 years; diagnosis of psychotic disorder; good command of the Norwegian language to complete interview and neurocognitive tests | Substance-induced psychotic disorder; IQ < 70; previous moderate to severe head injury; organic brain disease |
| | CTR | K-SADS-PL (2009) | Invitation by letter after random draw from the | Age between 12-18 years; good command of the Norwegian | History of mental health issues (contact with specialist services); previous or current use of |

| | | | | | |
|---|---|---|---|---|---|
| | | | Norwegian National Registry | language to complete interview and neurocognitive tests | psychotropic medication; first-degree relatives with a history of psychotic disorders; IQ < 70; previous moderate to severe head injury; organic brain disease |

* Gurholt et al. 2020. Human Brain Mapping; Abbreviation: EOP = early-onset psychosis; CTR = healthy controls; DSM = Structured Clinical Interview for Diagnostic and Statistical Manual of Mental Disorder; SCID = Structured Clinical Interview for DSM Disorders; K-SADS = Kiddie Schedule for Affective Disorders and Schizophrenia (PL = present and lifetime version); IQ = intelligence quotient; MRI = magnetic resonance imaging.

#### Table S3| Diffusion weighted imaging acquisitions parameters, stratified by site and scanner.

| Site | Scanner | Field Strength (T. Tesla) | Voxel size and slice thickness | Gradient directions and b-value (mm/s$^2$) | b=0 scans | TR (ms) | TE (ms) | Flip Angle (°) |
|---|---|---|---|---|---|---|---|---|
| BARCELONA | Siemens Trio Tim | 3T | 2.0x2.0x2.0 | 30 at b = 800 | 1 | 8600 | 97 | 90 |
| | Siemens Prisma | 3T | 2.0x2.0x2.0 | 30 at b = 800 | 1 | 8600 | 97 | 90 |
| | GE Genesis Signa | 1.5T | 1.0x1.0x5.0 (n=40)<br>1.17x1.17x5.0 (n=6) | 25 at b = 100 | 1 | 10000 | [79.9 – 93.1]<br>(mean: 86.2) | 90 |
| FEMS | Siemens TrioTim | 3T | 2.0x2.0x2.0 | 60 at b = 2000 | 10 | 8800 | 99 | 90 |
| MADRID | Philips Intera | 1.5T | 2.0x2.0x2.0 | 32 at b = 1000 | 1 | 11888 | 71 | 30 |
| OXFORD | Siemens Sonata | 1.5T | 2.5x2.5x2.5 | 60 at b = 1000 | 5 | 8500 | 89 | 19 |
| PAFIP | Philips | 3T | 2.0x2.0x2.0 | 64 at b = 1300 | 1 | 9577 | 77 | 90 |
| ROME | Philips Achieva | 3T | 2x2x2 | 128 at b = 1000 | 1 | 10000 | 70 | 90 |
| | | | 2x2x2 | 120 at b = 1000 | 1 | 10000 | 76 | 90 |
| | | | 2x2x2 | 64 at b = 1000 | 6 | 10000 | 76 | 90 |
| SCAPS | GE Discovery MR750 | 3T | 0.94 × 0.94 × 2.9 | 60 at b = 1000 | 10 | 6000 | 82.9 | 90 |
| UCLA | Siemens TrioTim | 3T | 1.0x1.0x1.2 | 64 at b = 1000 | | 2300 | 2.91 | 9 |
| YTOP | GE Signa HDxt | 3T | 1.875x1.875x2.5 | 30 at b = 1000 | 2 | 15000 | 85 | 90 |
| | GE Discovery MR750 | 3T | 2.0x2.0x2.0 | 60 at b = 1000 | 5 | 8150 | 83 | 90 |

Abbreviations: GE = general electrics, TR = repetition time, TE = echo time.

**Table S4| Demographic and clinical characteristics, stratified by site.**

| Cohort | Dx | N | Age (years) | Sex, Female N (%) | Hand (R/L/A) | Dx (EOS/ AFP/ OTP) | PANSS, negative | PANSS, positive | AOO (years) | DOI (years) | CPZ | AP user, N (%) | LIT user, N (%) | AD user, N (%) | AE user, N (%) |
|---|---|---|---|---|---|---|---|---|---|---|---|---|---|---|---|
| BARCELONA | CTR | 84 | 16.6 [14.9. 17.5] | 51 (60.7) | 15/3/0 | | | | | | | | | | |
| | EOP | 141 | 16.5 [15.1, 17.3] | 70 (49.6) | 36 /1/0 | 57/68/16 | 15.0 [11.0, 21.0] | 19.0 [15.0, 24.0] | 15.9 [14.6, 16.9] | 0.2 [0.1, 0.5] | 200.0 [150.0, 350.0] | 135 (98.5) | 23 (16.8) | 47 (34.3) | 4 (2.9) |
| FEMS | CTR | 2 | 16.0 [16.0. 16.0] | 1 (50.0) | 2/0/0 | | | | | | | | | | |
| | EOP | 5 | 18.0 [17.0. 18.0] | 1 (20.0) | 3/1/0 | 1/4/0 | | | 16.5 [16.3. 16.8] | 0.6 [0.5. 0.7] | | 2 (100.0) | 3 (100.0) | | |
| MADRID | CTR | 26 | 16.0 [13.3. 16.8] | 12 (46.2) | 20/2/0 | | | | | | | | | | |
| | EOP | 28 | 16.5 [14.0. 17.0] | 6 (21.4) | 23/2/0 | 28/0/0 | 23.0 [16.0. 29.0] | 23.5 [19.8. 30.5] | 16.0 [14.8. 17.0] | 0.0 [0.0. 1.0] | | 28 (100.0) | 0 (0.0) | 1 (3.6) | 2 (7.1) |
| OXFORD | CTR | 38 | 16.2 [14.9. 17.2] | 20 (52.6) | 34/4/0 | | | | | | | | | | |
| | EOP | 43 | 16.7 [15.4. 17.0] | 18 (41.9) | 33/8/2 | 43/0/0 | 16.0 [13.5. 18.0] | 23.0 [21.0. 24.0] | 14.8 [13.6. 15.9] | 1.3 [0.9. 2.2] | 300.0 [200.0, 424.0] | 43 (100.0) | 0 (0.0) | | 3 (7.0) |
| PAFIP* | CTR | 3 | 18.6 [18.5. 18.8] | 3 (100.0) | 3/0/0 | | | | | | | | | | |
| | EOP | 4 | 17.6 [17.5. 17.9] | 2 (50.0) | 4/0/0 | 4/0/0 | 6.0* [3.0, 11.8] | 15.0* [14.8, 16.3] | 17.4 [17.3. 17.5] | 0.4 [0.2. 0.7] | 200.0 [170.0, 200.0] | 4 (100.0) | | | |
| ROME | CTR | 12 | 14.0 [13.0. 15.0] | 8 (66.7) | 10/2/0 | | | | | | | | | | |
| | EOP | 19 | 16.0 [15.5. 18.0] | 6 (31.6) | 16/3/0 | 13/1/5 | 14.0 [10.5. 19.0] | 15.0 [14.0. 25.5] | 15.0 [14.0. 16.5] | 1.0 [1.0. 1.5] | 200.0 [166.7, 325.0] | 19 (100.0) | 1 (5.3) | 3 (15.8) | 4 (21.1) |

#### Supplemental Material

| | | | | | | | | | | | | | | | |
|---|---|---|---|---|---|---|---|---|---|---|---|---|---|---|---|
| SCAPS | CTR | 24 | 16.9<br>[16.2. 17.7] | 19<br>(79.2) | 22/2/0 | | | | | | | | | | |
| | EOP | 31 | 16.7<br>[15.7. 17.5] | 17<br>(54.8) | 7/1/0 | 2/20/9 | | | 15.2<br>[14.1. 16.6] | 1.0<br>[0.1. 2.3] | 133.0<br>[0.0, 241.5] | | | | |
| UCLA* | CTR | 7 | 16.0<br>[14.0. 16.5] | 2<br>(28.6) | 6/1/0 | | | | | | | | | | |
| | EOP | 13 | 16.0<br>[14.0. 17.0] | 3<br>(23.1) | 13/0/0 | 12/0/1 | 29.0*<br>[23.5, 31.5] | 24.0*<br>[8.0, 25.5] | 14.0<br>[12.0. 15.0] | 2.0<br>[1.0. 2.0] | 67.0<br>[0.0, 133.0] | 8<br>(61.5) | 0<br>(0.0) | 3<br>(25.0) | 1<br>(8.3) |
| YTOP | CTR | 69 | 16.2<br>[15.2. 17.4] | 37<br>(53.6) | 65/4/0 | | | | | | | | | | |
| | EOP | 37 | 16.5<br>[15.9. 17.6] | 26<br>(70.3) | 31/3/0 | 20/2/15 | 20.0<br>[14.0. 23.0] | 18.0<br>[15.0. 21.0] | 15.3<br>[13.8. 16.2] | 1.1<br>[0.6. 1.8] | 25.0<br>[0.0, 133.5] | 17<br>(51.5) | 0<br>(0.0) | 3<br>(9.4) | 0<br>(0.0) |

* These groups used SANS and SAPS scores (Scale for the Assessment of Negative/Positive Symptoms). Continuous data in median [Interquartile range] and categorical data as number (%). Abbreviations: Dx = Diagnosis, CTR = healthy controls, EOP = early-onset psychosis, R = right, L = left, A = ambidextrous, EOS = early-onset schizophrenia, AFP = affective psychosis, OTP = other psychosis, PANSS = positive and negative syndrome scale, AOO = age of onset, DOI = duration of illness, CPZ = chlorpromazine equivalent, AP = antipsychotics, LIT = lithium, AD = antidepressants, AE = antiepileptics.

**Table S5| Demographic and clinical characteristics, stratified by sex.**

| | Male | | Female | | | |
|---|---|---|---|---|---|---|
| **Variables** | **CTR** | **EOP** | **CTR** | **EOP** | **p-value** | **test** |
| **N** | 112 | 172 | 153 | 149 | | χ2 |
| **Age** (years)* | 16.19 [14.86, 17.38] | 16.66 [15.45, 17.40] | 16.18 [14.89, 17.27] | 16.48 [15.07, 17.30] | 0.259 | KW |
| **Handedness**, N (%) | | | | | 0.756 | χ2 |
| Right | 74 (89.2) | 95 (90.5) | 103 (92.0) | 71 (86.6) | | |
| Left | 9 (10.8) | 9 (8.6) | 9 (8.0) | 10 (12.2) | | |
| Ambidextrous | 0 (0.0) | 1 (1.0) | 0 (0.0) | 1 (1.2) | | |
| **Diagnostic subgroup**, N (%) | | | | | **0.009** | |
| EOS | | 110 (64.0) | | 70 (47.0) | | |
| AFP | | 41 (23.8) | | 54 (36.2) | | |
| OTP | | 21 (12.2) | | 25 (16.8) | | |
| **PANSS, negative*** | | 17.00 [12.25, 22.00] | | 16.00 [12.00, 21.00] | 0.375 | KW |
| **PANSS, positive*** | | 22.00 [17.25, 25.00] | | 19.00 [15.00, 23.00] | **<0.001** | KW |
| **Age of onset** (years)* | | 15.88 [14.50, 16.91] | | 15.16 [13.99, 16.30] | **0.009** | KW |
| **Duration of illness** (years)* | | 0.59 [0.08, 1.08] | | 0.64 [0.17, 1.14] | 0.144 | KW |
| **CPZ*** | | 200.0 [133.3, 300.0] | | 200.0 [116.7, 374.8] | 0.929 | |
| **AP user**, N (%) | | 144 (92.9) | | 112 (86.2) | 0.093 | χ2 |
| **Lithium user**, N (%) | | 17 (11.3) | | 10 (8.1) | 0.484 | χ2 |
| **AD user**, N (%) | | 23 (18.7) | | 34 (32.4) | **0.026** | χ2 |
| **AE user**, N (%) | | 7 (4.7) | | 7 (5.7) | 0.936 | χ2 |
| **Field strength**, 3T, N (%) | 38 (50.7) | 84 (57.1) | 57 (58.8) | 69 (65.1) | 0.272 | χ2 |

*Continuous data in median [Interquartile range] and categorical data as number (%). Abbreviations: CTR = healthy controls, EOP = early-onset psychosis, N = number, EOS = early-onset schizophrenia, AFP = affective psychosis, OTP = other psychosis, PANSS = positive and negative syndrome scale, CPZ = chlorpromazine equivalent, AP = antipsychotics, AD = antidepressants, AE = antiepileptics, KW = Kruskal-Wallis. Significant results are highlighted in bold.

#### *Multiple Linear Regression Outputs*

**Table S6| Multiple linear regression output for case-control differences in bilateral regional diffusion measures.**

| Tract | metric | t-value | p-value | Cohen's *d* | S.E. |
|---|---|---|---|---|---|
| ACR | FA | -1.891 | 0.059 | -0.158 | 0.083 |
| | MD | 0.508 | 0.612 | 0.046 | 0.089 |
| | RD | 1.421 | 0.156 | 0.127 | 0.089 |
| | AD | -0.537 | 0.592 | -0.048 | 0.089 |
| ALIC | FA | -2.986 | 0.003 | -0.249 | 0.083 |
| | MD | 0.042 | 0.966 | 0.004 | 0.089 |
| | RD | 1.762 | 0.079 | 0.158 | 0.089 |
| | AD | -1.115 | 0.265 | -0.100 | 0.089 |
| Average | FA | -3.606 | **3.374e-04** | -0.301 | 0.083 |
| | MD | 1.863 | 0.063 | 0.167 | 0.089 |
| | RD | 3.282 | **0.001** | 0.294 | 0.089 |
| | AD | 0.426 | 0.671 | 0.038 | 0.089 |
| BCC | FA | -2.842 | 0.005 | -0.237 | 0.083 |
| | MD | 0.209 | 0.834 | 0.019 | 0.089 |
| | RD | 2.126 | 0.034 | 0.191 | 0.089 |
| | AD | -1.709 | 0.088 | -0.153 | 0.089 |
| CC | FA | -3.316 | **0.001** | -0.277 | 0.083 |
| | MD | 0.130 | 0.897 | 0.012 | 0.089 |
| | RD | 2.470 | 0.014 | 0.221 | 0.089 |
| | AD | -1.809 | 0.071 | -0.162 | 0.089 |
| CGC | FA | -2.449 | 0.015 | -0.204 | 0.083 |
| | MD | 0.624 | 0.533 | 0.056 | 0.089 |
| | RD | 3.068 | **0.002** | 0.275 | 0.089 |
| | AD | -0.998 | 0.319 | -0.089 | 0.089 |
| CGH | FA | -0.922 | 0.357 | -0.077 | 0.083 |
| | MD | -0.603 | 0.547 | -0.054 | 0.089 |
| | RD | 0.286 | 0.775 | 0.026 | 0.089 |
| | AD | -0.552 | 0.581 | -0.049 | 0.089 |
| CR | FA | -2.618 | 0.009 | -0.219 | 0.083 |
| | MD | 1.076 | 0.282 | 0.096 | 0.089 |
| | RD | 2.320 | 0.021 | 0.208 | 0.089 |
| | AD | -0.494 | 0.622 | -0.044 | 0.089 |
| CST | FA | -0.557 | 0.577 | -0.047 | 0.083 |
| | MD | 0.249 | 0.803 | 0.022 | 0.089 |
| | RD | 1.129 | 0.259 | 0.101 | 0.089 |
| | AD | -0.595 | 0.552 | -0.053 | 0.089 |
| EC | FA | -2.242 | 0.025 | -0.187 | 0.083 |
| | MD | 1.165 | 0.245 | 0.104 | 0.089 |
| | RD | 2.054 | 0.040 | 0.184 | 0.089 |
| | AD | 0.289 | 0.773 | 0.026 | 0.089 |
| FX | FA | -2.102 | 0.036 | -0.175 | 0.083 |

| | | | | | |
|---|---|---|---|---|---|
| | MD | 3.871 | **1.226e-04** | 0.347 | 0.089 |
| | RD | 3.891 | **1.135e-04** | 0.349 | 0.089 |
| | AD | 3.751 | **1.964e-04** | 0.336 | 0.089 |
| FXST | FA | -2.883 | 0.004 | -0.241 | 0.083 |
| | MD | 0.243 | 0.808 | 0.022 | 0.089 |
| | RD | 2.082 | 0.038 | 0.187 | 0.089 |
| | AD | -1.086 | 0.278 | -0.097 | 0.089 |
| GCC | FA | -3.352 | **0.001** | -0.280 | 0.083 |
| | MD | 0.042 | 0.966 | 0.004 | 0.089 |
| | RD | 2.338 | 0.020 | 0.210 | 0.089 |
| | AD | -1.729 | 0.084 | -0.155 | 0.089 |
| IC | FA | -3.501 | **4.999e-04** | -0.292 | 0.083 |
| | MD | 0.086 | 0.932 | 0.008 | 0.089 |
| | RD | 2.106 | 0.036 | 0.189 | 0.089 |
| | AD | -1.666 | 0.096 | -0.149 | 0.089 |
| IFO | FA | -1.430 | 0.153 | -0.119 | 0.083 |
| | MD | 1.287 | 0.199 | 0.115 | 0.089 |
| | RD | 1.787 | 0.074 | 0.160 | 0.089 |
| | AD | -0.248 | 0.804 | -0.022 | 0.089 |
| PCR | FA | -3.830 | **1.422e-04** | -0.320 | 0.083 |
| | MD | 2.366 | 0.018 | 0.212 | 0.089 |
| | RD | 3.467 | **0.001** | 0.311 | 0.089 |
| | AD | 0.109 | 0.913 | 0.010 | 0.089 |
| PLIC | FA | -2.348 | 0.019 | -0.196 | 0.083 |
| | MD | -0.405 | 0.686 | -0.036 | 0.089 |
| | RD | 1.194 | 0.233 | 0.107 | 0.089 |
| | AD | -1.285 | 0.199 | -0.115 | 0.089 |
| PTR | FA | -3.123 | **0.002** | -0.261 | 0.083 |
| | MD | 2.164 | 0.031 | 0.194 | 0.089 |
| | RD | 3.088 | **0.002** | 0.277 | 0.089 |
| | AD | 0.165 | 0.869 | 0.015 | 0.089 |
| RLIC | FA | -3.214 | **0.001** | -0.268 | 0.083 |
| | MD | 0.703 | 0.483 | 0.063 | 0.089 |
| | RD | 2.412 | 0.016 | 0.216 | 0.089 |
| | AD | -1.402 | 0.162 | -0.126 | 0.089 |
| SCC | FA | -2.458 | 0.014 | -0.205 | 0.083 |
| | MD | 0.480 | 0.632 | 0.043 | 0.089 |
| | RD | 1.976 | 0.049 | 0.177 | 0.089 |
| | AD | -0.846 | 0.398 | -0.076 | 0.089 |
| SCR | FA | -1.753 | 0.080 | -0.146 | 0.083 |
| | MD | 0.805 | 0.421 | 0.072 | 0.089 |
| | RD | 2.047 | 0.041 | 0.183 | 0.089 |
| | AD | -0.511 | 0.609 | -0.046 | 0.089 |
| SFO | FA | -3.716 | **2.220e-04** | -0.310 | 0.083 |
| | MD | 0.336 | 0.737 | 0.030 | 0.089 |
| | RD | 2.231 | 0.026 | 0.200 | 0.089 |

| | | | | | |
|---|---|---|---|---|---|
| | AD | -1.438 | 0.151 | -0.129 | 0.089 |
| SLF | FA | -4.426 | **1.150e-05** | -0.370 | 0.083 |
| | MD | 2.113 | 0.035 | 0.189 | 0.089 |
| | RD | 3.975 | **8.071e-05** | 0.356 | 0.090 |
| | AD | -0.871 | 0.384 | -0.078 | 0.089 |
| SS | FA | -1.755 | 0.080 | -0.147 | 0.083 |
| | MD | 2.020 | 0.044 | 0.181 | 0.089 |
| | RD | 2.825 | 0.005 | 0.253 | 0.089 |
| | AD | 0.386 | 0.700 | 0.035 | 0.089 |
| UNC | FA | -2.249 | 0.025 | -0.188 | 0.083 |
| | MD | 3.242 | **0.001** | 0.291 | 0.089 |
| | RD | 3.294 | **0.001** | 0.295 | 0.089 |
| | AD | 2.089 | 0.037 | 0.187 | 0.089 |

Significant results are highlighted in bold ($p < 0.002$). Abbreviation: FA = fractional anisotropy, MD = mean diffusivity, RD = radial diffusivity, AD = axial diffusivity, S.E. = standard error. Abbreviations for tracts see Table 1 in main text.

**Table S7| Multiple linear regression output for case-control differences in bilateral regional diffusion measures, adjusted for average, core or periphery diffusion measures.**

| Tract | Adjustment | metric | t-value | p-value | Cohen's *d* | S.E. |
|---|---|---|---|---|---|---|
| ACR | Average | FA | 0.482 | 0.630 | 0.040 | 0.083 |
| | Core | FA | 1.759 | 0.079 | 0.147 | 0.083 |
| | Periphery | FA | -0.404 | 0.686 | -0.034 | 0.083 |
| | Average | MD | -0.503 | 0.615 | -0.045 | 0.089 |
| | Core | MD | -0.909 | 0.364 | -0.081 | 0.089 |
| | Periphery | MD | -0.247 | 0.805 | -0.022 | 0.089 |
| | Average | RD | -0.555 | 0.579 | -0.050 | 0.089 |
| | Core | RD | -1.967 | 0.050 | -0.176 | 0.089 |
| | Periphery | RD | 0.017 | 0.987 | 0.001 | 0.089 |
| | Average | AD | -0.712 | 0.477 | -0.064 | 0.089 |
| | Core | AD | 0.268 | 0.789 | 0.024 | 0.089 |
| | Periphery | AD | -0.684 | 0.494 | -0.061 | 0.089 |
| ALIC | Average | FA | -1.335 | 0.183 | -0.112 | 0.083 |
| | Core | FA | -0.510 | 0.610 | -0.043 | 0.083 |
| | Periphery | FA | -1.940 | 0.053 | -0.162 | 0.083 |
| | Average | MD | -0.807 | 0.420 | -0.072 | 0.089 |
| | Core | MD | -1.241 | 0.215 | -0.111 | 0.089 |
| | Periphery | MD | -0.576 | 0.565 | -0.052 | 0.089 |
| | Average | RD | 0.401 | 0.689 | 0.036 | 0.089 |
| | Core | RD | -0.580 | 0.562 | -0.052 | 0.089 |
| | Periphery | RD | 0.816 | 0.415 | 0.073 | 0.089 |
| | Average | AD | -1.255 | 0.210 | -0.113 | 0.089 |
| | Core | AD | -0.655 | 0.512 | -0.059 | 0.089 |
| | Periphery | AD | -1.219 | 0.224 | -0.109 | 0.089 |
| Average | Core | FA | -1.005 | 0.315 | -0.084 | 0.083 |
| | Periphery | FA | -2.547 | 0.011 | -0.213 | 0.083 |
| | Core | MD | 1.368 | 0.172 | 0.123 | 0.089 |
| | Periphery | MD | 0.335 | 0.738 | 0.030 | 0.089 |
| | Core | RD | 1.481 | 0.139 | 0.133 | 0.089 |
| | Periphery | RD | 1.844 | 0.066 | 0.165 | 0.089 |
| | Core | AD | 1.028 | 0.305 | 0.092 | 0.089 |
| | Periphery | AD | -1.254 | 0.210 | -0.112 | 0.089 |
| BCC | Average | FA | -0.911 | 0.363 | -0.076 | 0.083 |
| | Core | FA | -0.144 | 0.885 | -0.012 | 0.083 |
| | Periphery | FA | -1.583 | 0.114 | -0.132 | 0.083 |
| | Average | MD | -0.847 | 0.397 | -0.076 | 0.089 |
| | Core | MD | -1.083 | 0.279 | -0.097 | 0.089 |
| | Periphery | MD | -0.606 | 0.545 | -0.054 | 0.089 |
| | Average | RD | 0.423 | 0.673 | 0.038 | 0.089 |
| | Core | RD | -0.314 | 0.754 | -0.028 | 0.089 |

| | | | | | | |
|---|---|---|---|---|---|---|
| | Periphery | RD | 0.870 | 0.385 | 0.078 | 0.089 |
| | Average | AD | -2.012 | 0.045 | -0.181 | 0.089 |
| | Core | AD | -1.482 | 0.139 | -0.133 | 0.089 |
| | Periphery | AD | -1.943 | 0.053 | -0.174 | 0.089 |
| CC | Average | FA | -1.334 | 0.183 | -0.111 | 0.083 |
| | Core | FA | -0.156 | 0.876 | -0.013 | 0.083 |
| | Periphery | FA | -2.083 | 0.038 | -0.174 | 0.083 |
| | Average | MD | -1.144 | 0.253 | -0.103 | 0.089 |
| | Core | MD | -1.961 | 0.050 | -0.176 | 0.089 |
| | Periphery | MD | -0.817 | 0.414 | -0.073 | 0.089 |
| | Average | RD | 0.596 | 0.552 | 0.053 | 0.089 |
| | Core | RD | -0.571 | 0.568 | -0.051 | 0.089 |
| | Periphery | RD | 1.123 | 0.262 | 0.101 | 0.089 |
| | Average | AD | -2.119 | 0.035 | -0.190 | 0.089 |
| | Core | AD | -1.817 | 0.070 | -0.163 | 0.089 |
| | Periphery | AD | -2.026 | 0.043 | -0.182 | 0.089 |
| CGC | Average | FA | -0.719 | 0.472 | -0.060 | 0.083 |
| | Core | FA | 0.143 | 0.887 | 0.012 | 0.083 |
| | Periphery | FA | -1.352 | 0.177 | -0.113 | 0.083 |
| | Average | MD | -0.439 | 0.661 | -0.039 | 0.089 |
| | Core | MD | -0.731 | 0.465 | -0.065 | 0.089 |
| | Periphery | MD | -0.185 | 0.853 | -0.017 | 0.089 |
| | Average | RD | 1.574 | 0.116 | 0.141 | 0.089 |
| | Core | RD | 0.955 | 0.340 | 0.086 | 0.089 |
| | Periphery | RD | 1.974 | 0.049 | 0.177 | 0.089 |
| | Average | AD | -1.133 | 0.258 | -0.102 | 0.089 |
| | Core | AD | -0.528 | 0.598 | -0.047 | 0.089 |
| | Periphery | AD | -1.104 | 0.270 | -0.099 | 0.089 |
| CGH | Average | FA | 0.891 | 0.373 | 0.074 | 0.083 |
| | Core | FA | 1.087 | 0.277 | 0.091 | 0.083 |
| | Periphery | FA | 0.387 | 0.699 | 0.032 | 0.083 |
| | Average | MD | -1.508 | 0.132 | -0.135 | 0.089 |
| | Core | MD | -1.679 | 0.094 | -0.150 | 0.089 |
| | Periphery | MD | -1.311 | 0.191 | -0.117 | 0.089 |
| | Average | RD | -1.041 | 0.298 | -0.093 | 0.089 |
| | Core | RD | -1.788 | 0.074 | -0.160 | 0.089 |
| | Periphery | RD | -0.670 | 0.503 | -0.060 | 0.089 |
| | Average | AD | -0.732 | 0.465 | -0.066 | 0.089 |
| | Core | AD | -0.072 | 0.942 | -0.006 | 0.089 |
| | Periphery | AD | -0.742 | 0.458 | -0.067 | 0.089 |
| CR | Average | FA | -0.220 | 0.826 | -0.018 | 0.083 |
| | Core | FA | 1.723 | 0.085 | 0.144 | 0.083 |
| | Periphery | FA | -1.171 | 0.242 | -0.098 | 0.083 |

| | | | | | | |
|---|---|---|---|---|---|---|
| | Average | MD | -0.020 | 0.984 | -0.002 | 0.089 |
| | Core | MD | -0.240 | 0.810 | -0.022 | 0.089 |
| | Periphery | MD | 0.244 | 0.807 | 0.022 | 0.089 |
| | Average | RD | 0.337 | 0.737 | 0.030 | 0.089 |
| | Core | RD | -1.494 | 0.136 | -0.134 | 0.089 |
| | Periphery | RD | 0.924 | 0.356 | 0.083 | 0.089 |
| | Average | AD | -0.727 | 0.468 | -0.065 | 0.089 |
| | Core | AD | 0.648 | 0.517 | 0.058 | 0.089 |
| | Periphery | AD | -0.695 | 0.487 | -0.062 | 0.089 |
| CST | Average | FA | 0.370 | 0.712 | 0.031 | 0.083 |
| | Core | FA | 0.769 | 0.442 | 0.064 | 0.083 |
| | Periphery | FA | 0.044 | 0.965 | 0.004 | 0.083 |
| | Average | MD | -0.062 | 0.951 | -0.006 | 0.089 |
| | Core | MD | -0.114 | 0.909 | -0.010 | 0.089 |
| | Periphery | MD | 0.013 | 0.990 | 0.001 | 0.089 |
| | Average | RD | 0.434 | 0.665 | 0.039 | 0.089 |
| | Core | RD | 0.057 | 0.955 | 0.005 | 0.089 |
| | Periphery | RD | 0.632 | 0.528 | 0.057 | 0.089 |
| | Average | AD | -0.644 | 0.520 | -0.058 | 0.089 |
| | Core | AD | -0.405 | 0.686 | -0.036 | 0.089 |
| | Periphery | AD | -0.646 | 0.519 | -0.058 | 0.089 |
| EC | Average | FA | -0.187 | 0.852 | -0.016 | 0.083 |
| | Core | FA | 0.745 | 0.457 | 0.062 | 0.083 |
| | Periphery | FA | -0.924 | 0.356 | -0.077 | 0.083 |
| | Average | MD | 0.196 | 0.845 | 0.018 | 0.089 |
| | Core | MD | 0.236 | 0.813 | 0.021 | 0.089 |
| | Periphery | MD | 0.406 | 0.685 | 0.036 | 0.089 |
| | Average | RD | 0.398 | 0.691 | 0.036 | 0.089 |
| | Core | RD | -0.595 | 0.552 | -0.053 | 0.089 |
| | Periphery | RD | 0.867 | 0.387 | 0.078 | 0.089 |
| | Average | AD | 0.157 | 0.875 | 0.014 | 0.089 |
| | Core | AD | 1.186 | 0.236 | 0.106 | 0.089 |
| | Periphery | AD | 0.143 | 0.887 | 0.013 | 0.089 |
| FX | Average | FA | -1.277 | 0.202 | -0.107 | 0.083 |
| | Core | FA | -0.823 | 0.411 | -0.069 | 0.083 |
| | Periphery | FA | -1.595 | 0.111 | -0.133 | 0.083 |
| | Average | MD | 3.411 | **0.001** | 0.306 | 0.089 |
| | Core | MD | 3.651 | **2.89e-04** | 0.327 | 0.089 |
| | Periphery | MD | 3.433 | **0.001** | 0.308 | 0.089 |
| | Average | RD | 2.838 | 0.005 | 0.255 | 0.089 |
| | Core | RD | 2.964 | 0.003 | 0.266 | 0.089 |
| | Periphery | RD | 2.998 | 0.003 | 0.269 | 0.089 |
| | Average | AD | 3.805 | **1.59e-04** | 0.341 | 0.089 |

| | | | | | | |
|---|---|---|---|---|---|---|
| | Core | AD | 4.097 | **4.88e-05** | 0.367 | 0.090 |
| | Periphery | AD | 3.722 | **2.20e-04** | 0.334 | 0.089 |
| FXST | Average | FA | -1.525 | 0.128 | -0.127 | 0.083 |
| | Core | FA | -1.066 | 0.287 | -0.089 | 0.083 |
| | Periphery | FA | -1.972 | 0.049 | -0.165 | 0.083 |
| | Average | MD | -0.702 | 0.483 | -0.063 | 0.089 |
| | Core | MD | -0.855 | 0.393 | -0.077 | 0.089 |
| | Periphery | MD | -0.493 | 0.623 | -0.044 | 0.089 |
| | Average | RD | 0.665 | 0.507 | 0.060 | 0.089 |
| | Core | RD | 0.073 | 0.942 | 0.007 | 0.089 |
| | Periphery | RD | 1.034 | 0.302 | 0.093 | 0.089 |
| | Average | AD | -1.241 | 0.215 | -0.111 | 0.089 |
| | Core | AD | -0.687 | 0.493 | -0.062 | 0.089 |
| | Periphery | AD | -1.227 | 0.221 | -0.110 | 0.089 |
| GCC | Average | FA | -1.623 | 0.105 | -0.136 | 0.083 |
| | Core | FA | -0.618 | 0.537 | -0.052 | 0.083 |
| | Periphery | FA | -2.282 | 0.023 | -0.191 | 0.083 |
| | Average | MD | -1.006 | 0.315 | -0.090 | 0.089 |
| | Core | MD | -1.469 | 0.143 | -0.132 | 0.089 |
| | Periphery | MD | -0.742 | 0.458 | -0.067 | 0.089 |
| | Average | RD | 0.655 | 0.513 | 0.059 | 0.089 |
| | Core | RD | -0.344 | 0.731 | -0.031 | 0.089 |
| | Periphery | RD | 1.130 | 0.259 | 0.101 | 0.089 |
| | Average | AD | -1.918 | 0.056 | -0.172 | 0.089 |
| | Core | AD | -1.488 | 0.137 | -0.133 | 0.089 |
| | Periphery | AD | -1.854 | 0.064 | -0.166 | 0.089 |
| IC | Average | FA | -1.669 | 0.096 | -0.139 | 0.083 |
| | Core | FA | -0.563 | 0.574 | -0.047 | 0.083 |
| | Periphery | FA | -2.370 | 0.018 | -0.198 | 0.083 |
| | Average | MD | -0.993 | 0.321 | -0.089 | 0.089 |
| | Core | MD | -1.900 | 0.058 | -0.170 | 0.089 |
| | Periphery | MD | -0.691 | 0.490 | -0.062 | 0.089 |
| | Average | RD | 0.525 | 0.600 | 0.047 | 0.089 |
| | Core | RD | -0.885 | 0.377 | -0.079 | 0.089 |
| | Periphery | RD | 1.021 | 0.308 | 0.092 | 0.089 |
| | Average | AD | -1.933 | 0.054 | -0.173 | 0.089 |
| | Core | AD | -1.514 | 0.131 | -0.136 | 0.089 |
| | Periphery | AD | -1.852 | 0.065 | -0.166 | 0.089 |
| IFO | Average | FA | -0.261 | 0.794 | -0.022 | 0.083 |
| | Core | FA | 0.285 | 0.776 | 0.024 | 0.083 |
| | Periphery | FA | -0.683 | 0.495 | -0.057 | 0.083 |
| | Average | MD | 0.771 | 0.441 | 0.069 | 0.089 |
| | Core | MD | 0.798 | 0.425 | 0.072 | 0.089 |

| | | | | | | |
|---|---|---|---|---|---|---|
| | Periphery | MD | 0.875 | 0.382 | 0.078 | 0.089 |
| | Average | RD | 0.852 | 0.395 | 0.076 | 0.089 |
| | Core | RD | 0.354 | 0.724 | 0.032 | 0.089 |
| | Periphery | RD | 1.117 | 0.265 | 0.100 | 0.089 |
| | Average | AD | -0.328 | 0.743 | -0.029 | 0.089 |
| | Core | AD | 0.092 | 0.927 | 0.008 | 0.089 |
| | Periphery | AD | -0.329 | 0.742 | -0.030 | 0.089 |
| PCR | Average | FA | -2.109 | 0.035 | -0.176 | 0.083 |
| | Core | FA | -1.342 | 0.180 | -0.112 | 0.083 |
| | Periphery | FA | -2.725 | 0.007 | -0.228 | 0.083 |
| | Average | MD | 1.595 | 0.111 | 0.143 | 0.089 |
| | Core | MD | 2.450 | 0.015 | 0.220 | 0.089 |
| | Periphery | MD | 1.720 | 0.086 | 0.154 | 0.089 |
| | Average | RD | 1.977 | 0.049 | 0.177 | 0.089 |
| | Core | RD | 1.450 | 0.148 | 0.130 | 0.089 |
| | Periphery | RD | 2.377 | 0.018 | 0.213 | 0.089 |
| | Average | AD | -0.058 | 0.954 | -0.005 | 0.089 |
| | Core | AD | 1.236 | 0.217 | 0.111 | 0.089 |
| | Periphery | AD | -0.067 | 0.947 | -0.006 | 0.089 |
| PLIC | Average | FA | -0.824 | 0.411 | -0.069 | 0.083 |
| | Core | FA | 0.106 | 0.916 | 0.009 | 0.083 |
| | Periphery | FA | -1.418 | 0.157 | -0.118 | 0.083 |
| | Average | MD | -1.367 | 0.172 | -0.123 | 0.089 |
| | Core | MD | -1.898 | 0.058 | -0.170 | 0.089 |
| | Periphery | MD | -1.112 | 0.267 | -0.100 | 0.089 |
| | Average | RD | -0.196 | 0.845 | -0.018 | 0.089 |
| | Core | RD | -1.189 | 0.235 | -0.107 | 0.089 |
| | Periphery | RD | 0.223 | 0.824 | 0.020 | 0.089 |
| | Average | AD | -1.448 | 0.148 | -0.130 | 0.089 |
| | Core | AD | -0.883 | 0.378 | -0.079 | 0.089 |
| | Periphery | AD | -1.415 | 0.158 | -0.127 | 0.089 |
| PTR | Average | FA | -1.663 | 0.097 | -0.139 | 0.083 |
| | Core | FA | -0.743 | 0.458 | -0.062 | 0.083 |
| | Periphery | FA | -2.243 | 0.025 | -0.187 | 0.083 |
| | Average | MD | 1.484 | 0.138 | 0.133 | 0.089 |
| | Core | MD | 1.783 | 0.075 | 0.160 | 0.089 |
| | Periphery | MD | 1.608 | 0.108 | 0.144 | 0.089 |
| | Average | RD | 1.847 | 0.065 | 0.166 | 0.089 |
| | Core | RD | 1.127 | 0.260 | 0.101 | 0.089 |
| | Periphery | RD | 2.220 | 0.027 | 0.199 | 0.089 |
| | Average | AD | 0.039 | 0.969 | 0.004 | 0.089 |
| | Core | AD | 0.904 | 0.366 | 0.081 | 0.089 |
| | Periphery | AD | 0.030 | 0.976 | 0.003 | 0.089 |

| | | | | | | |
|---|---|---|---|---|---|---|
| RLIC | Average | FA | -1.576 | 0.115 | -0.132 | 0.083 |
| | Core | FA | -0.653 | 0.514 | -0.055 | 0.083 |
| | Periphery | FA | -2.199 | 0.028 | -0.184 | 0.083 |
| | Average | MD | -0.244 | 0.807 | -0.022 | 0.089 |
| | Core | MD | -0.614 | 0.540 | -0.055 | 0.089 |
| | Periphery | MD | 0.002 | 0.998 | 2.17e-04 | 0.089 |
| | Average | RD | 1.048 | 0.295 | 0.094 | 0.089 |
| | Core | RD | -0.103 | 0.918 | -0.009 | 0.089 |
| | Periphery | RD | 1.489 | 0.137 | 0.134 | 0.089 |
| | Average | AD | -1.652 | 0.099 | -0.148 | 0.089 |
| | Core | AD | -1.029 | 0.304 | -0.092 | 0.089 |
| | Periphery | AD | -1.593 | 0.112 | -0.143 | 0.089 |
| SCC | Average | FA | -0.862 | 0.389 | -0.072 | 0.083 |
| | Core | FA | 0.285 | 0.776 | 0.024 | 0.083 |
| | Periphery | FA | -1.513 | 0.131 | -0.126 | 0.083 |
| | Average | MD | -0.476 | 0.634 | -0.043 | 0.089 |
| | Core | MD | -0.745 | 0.457 | -0.067 | 0.089 |
| | Periphery | MD | -0.245 | 0.807 | -0.022 | 0.089 |
| | Average | RD | 0.388 | 0.699 | 0.035 | 0.089 |
| | Core | RD | -0.692 | 0.489 | -0.062 | 0.089 |
| | Periphery | RD | 0.853 | 0.394 | 0.076 | 0.089 |
| | Average | AD | -1.011 | 0.312 | -0.091 | 0.089 |
| | Core | AD | -0.257 | 0.797 | -0.023 | 0.089 |
| | Periphery | AD | -0.979 | 0.328 | -0.088 | 0.089 |
| SCR | Average | FA | 0.130 | 0.896 | 0.011 | 0.083 |
| | Core | FA | 1.604 | 0.109 | 0.134 | 0.083 |
| | Periphery | FA | -0.642 | 0.521 | -0.054 | 0.083 |
| | Average | MD | -0.317 | 0.751 | -0.028 | 0.089 |
| | Core | MD | -0.625 | 0.533 | -0.056 | 0.089 |
| | Periphery | MD | -0.049 | 0.961 | -0.004 | 0.089 |
| | Average | RD | 0.301 | 0.764 | 0.027 | 0.089 |
| | Core | RD | -1.026 | 0.305 | -0.092 | 0.089 |
| | Periphery | RD | 0.818 | 0.414 | 0.073 | 0.089 |
| | Average | AD | -0.726 | 0.468 | -0.065 | 0.089 |
| | Core | AD | 0.314 | 0.753 | 0.028 | 0.089 |
| | Periphery | AD | -0.708 | 0.479 | -0.063 | 0.089 |
| SFO | Average | FA | -2.330 | 0.020 | -0.195 | 0.083 |
| | Core | FA | -1.719 | 0.086 | -0.144 | 0.083 |
| | Periphery | FA | -2.825 | 0.005 | -0.236 | 0.083 |
| | Average | MD | -0.440 | 0.660 | -0.039 | 0.089 |
| | Core | MD | -0.628 | 0.530 | -0.056 | 0.089 |
| | Periphery | MD | -0.252 | 0.801 | -0.023 | 0.089 |
| | Average | RD | 0.959 | 0.338 | 0.086 | 0.089 |

| | | | | | | |
|---|---|---|---|---|---|---|
| | Core | RD | 0.180 | 0.857 | 0.016 | 0.089 |
| | Periphery | RD | 1.334 | 0.183 | 0.120 | 0.089 |
| | Average | AD | -1.602 | 0.110 | -0.144 | 0.089 |
| | Core | AD | -1.099 | 0.272 | -0.099 | 0.089 |
| | Periphery | AD | -1.581 | 0.114 | -0.142 | 0.089 |
| SLF | Average | FA | -2.781 | 0.006 | -0.232 | 0.083 |
| | Core | FA | -2.165 | 0.031 | -0.181 | 0.083 |
| | Periphery | FA | -3.360 | **0.001** | -0.281 | 0.083 |
| | Average | MD | 1.235 | 0.217 | 0.111 | 0.089 |
| | Core | MD | 2.146 | 0.032 | 0.192 | 0.089 |
| | Periphery | MD | 1.381 | 0.168 | 0.124 | 0.089 |
| | Average | RD | 2.514 | 0.012 | 0.226 | 0.089 |
| | Core | RD | 2.345 | 0.019 | 0.210 | 0.089 |
| | Periphery | RD | 2.880 | 0.004 | 0.258 | 0.089 |
| | Average | AD | -1.174 | 0.241 | -0.105 | 0.089 |
| | Core | AD | -0.157 | 0.876 | -0.014 | 0.089 |
| | Periphery | AD | -1.133 | 0.258 | -0.102 | 0.089 |
| SS | Average | FA | 0.245 | 0.807 | 0.020 | 0.083 |
| | Core | FA | 1.291 | 0.197 | 0.108 | 0.083 |
| | Periphery | FA | -0.495 | 0.621 | -0.041 | 0.083 |
| | Average | MD | 1.209 | 0.227 | 0.108 | 0.089 |
| | Core | MD | 1.782 | 0.075 | 0.160 | 0.089 |
| | Periphery | MD | 1.366 | 0.173 | 0.122 | 0.089 |
| | Average | RD | 1.352 | 0.177 | 0.121 | 0.089 |
| | Core | RD | 0.446 | 0.656 | 0.040 | 0.089 |
| | Periphery | RD | 1.785 | 0.075 | 0.160 | 0.089 |
| | Average | AD | 0.241 | 0.810 | 0.022 | 0.089 |
| | Core | AD | 1.558 | 0.120 | 0.140 | 0.089 |
| | Periphery | AD | 0.216 | 0.829 | 0.019 | 0.089 |
| UNC | Average | FA | -1.204 | 0.229 | -0.101 | 0.083 |
| | Core | FA | -0.533 | 0.594 | -0.045 | 0.083 |
| | Periphery | FA | -1.630 | 0.104 | -0.136 | 0.083 |
| | Average | MD | 2.688 | 0.007 | 0.241 | 0.089 |
| | Core | MD | 3.056 | **0.002** | 0.274 | 0.089 |
| | Periphery | MD | 2.745 | 0.006 | 0.246 | 0.089 |
| | Average | RD | 2.228 | 0.026 | 0.200 | 0.089 |
| | Core | RD | 1.806 | 0.072 | 0.162 | 0.089 |
| | Periphery | RD | 2.511 | 0.012 | 0.225 | 0.089 |
| | Average | AD | 2.056 | 0.040 | 0.185 | 0.089 |
| | Core | AD | 2.606 | 0.009 | 0.234 | 0.089 |
| | Periphery | AD | 1.987 | 0.047 | 0.178 | 0.089 |

Significant results are highlighted in bold ($p \leq 0.002$). Abbreviation: FA = fractional anisotropy, MD = mean diffusivity, RD = radial diffusivity, AD = axial diffusivity, S.E. = standard error. Abbreviations for tracts, see Table 1 in main text.

**Table S8| Multiple linear regression output for case-control differences in lateralized regional diffusion measures.**

| Tract | Side | metric | t-value | p-value | Cohen's *d* | S.E. |
|---|---|---|---|---|---|---|
| ACR | L | FA | -1.551 | 0.122 | -0.129 | 0.083 |
| | R | FA | -1.976 | 0.049 | -0.165 | 0.083 |
| ALIC | L | FA | -2.900 | 0.004 | -0.242 | 0.083 |
| | R | FA | -2.644 | 0.008 | -0.221 | 0.083 |
| CGC | L | FA | -2.658 | 0.008 | -0.222 | 0.083 |
| | R | FA | -1.970 | 0.049 | -0.165 | 0.083 |
| CGH | L | FA | -0.290 | 0.772 | -0.024 | 0.083 |
| | R | FA | -1.338 | 0.181 | -0.112 | 0.083 |
| CR | L | FA | -2.338 | 0.020 | -0.195 | 0.083 |
| | R | FA | -2.667 | 0.008 | -0.223 | 0.083 |
| CST | L | FA | -0.307 | 0.759 | -0.026 | 0.083 |
| | R | FA | -0.742 | 0.459 | -0.062 | 0.083 |
| EC | L | FA | -2.313 | 0.021 | -0.193 | 0.083 |
| | R | FA | -1.926 | 0.055 | -0.161 | 0.083 |
| FXST | L | FA | -2.862 | 0.004 | -0.239 | 0.083 |
| | R | FA | -2.588 | 0.010 | -0.216 | 0.083 |
| IC | L | FA | -3.873 | **1.197e-04** | -0.323 | 0.083 |
| | R | FA | -2.868 | 0.004 | -0.239 | 0.083 |
| IFO | L | FA | -1.110 | 0.267 | -0.093 | 0.083 |
| | R | FA | -1.324 | 0.186 | -0.111 | 0.083 |
| PCR | L | FA | -3.497 | **0.001** | -0.292 | 0.083 |
| | R | FA | -3.775 | **1.768e-04** | -0.315 | 0.083 |
| PLIC | L | FA | -2.564 | 0.011 | -0.214 | 0.083 |
| | R | FA | -1.927 | 0.055 | -0.161 | 0.083 |
| PTR | L | FA | -3.868 | **1.221e-04** | -0.323 | 0.083 |
| | R | FA | -1.979 | 0.048 | -0.165 | 0.083 |
| RLIC | L | FA | -3.780 | **1.733e-04** | -0.316 | 0.083 |
| | R | FA | -2.315 | 0.021 | -0.193 | 0.083 |
| SCR | L | FA | -1.709 | 0.088 | -0.143 | 0.083 |
| | R | FA | -1.565 | 0.118 | -0.131 | 0.083 |
| SFO | L | FA | -3.130 | **0.002** | -0.261 | 0.083 |
| | R | FA | -3.657 | **2.782e-04** | -0.305 | 0.083 |
| SLF | L | FA | -4.039 | **6.083e-05** | -0.337 | 0.083 |
| | R | FA | -4.266 | **2.324e-05** | -0.356 | 0.083 |
| SS | L | FA | -2.471 | 0.014 | -0.206 | 0.083 |
| | R | FA | -0.802 | 0.423 | -0.067 | 0.083 |
| UNC | L | FA | -1.028 | 0.304 | -0.086 | 0.083 |
| | R | FA | -2.797 | 0.005 | -0.234 | 0.083 |
| ACR | L | MD | 0.399 | 0.690 | 0.036 | 0.089 |
| | R | MD | 0.696 | 0.487 | 0.062 | 0.089 |
| ALIC | L | MD | -0.013 | 0.989 | -0.001 | 0.089 |
| | R | MD | 0.233 | 0.816 | 0.021 | 0.089 |

| | | | | | | |
|---|---|---|---|---|---|---|
| CGC | L | MD | 0.712 | 0.477 | 0.064 | 0.089 |
| | R | MD | 0.581 | 0.561 | 0.052 | 0.089 |
| CGH | L | MD | -0.772 | 0.441 | -0.069 | 0.089 |
| | R | MD | -0.294 | 0.769 | -0.026 | 0.089 |
| CR | L | MD | 1.164 | 0.245 | 0.104 | 0.089 |
| | R | MD | 0.938 | 0.349 | 0.084 | 0.089 |
| CST | L | MD | 0.098 | 0.922 | 0.009 | 0.089 |
| | R | MD | 0.454 | 0.650 | 0.041 | 0.089 |
| EC | L | MD | 1.307 | 0.192 | 0.117 | 0.089 |
| | R | MD | 0.851 | 0.395 | 0.076 | 0.089 |
| FXST | L | MD | -0.528 | 0.598 | -0.047 | 0.089 |
| | R | MD | 1.126 | 0.261 | 0.101 | 0.089 |
| IC | L | MD | -0.113 | 0.910 | -0.010 | 0.089 |
| | R | MD | 0.253 | 0.800 | 0.023 | 0.089 |
| IFO | L | MD | 1.414 | 0.158 | 0.127 | 0.089 |
| | R | MD | 0.919 | 0.359 | 0.082 | 0.089 |
| PCR | L | MD | 2.712 | 0.007 | 0.243 | 0.089 |
| | R | MD | 1.898 | 0.058 | 0.170 | 0.089 |
| PLIC | L | MD | -0.375 | 0.708 | -0.034 | 0.089 |
| | R | MD | -0.391 | 0.696 | -0.035 | 0.089 |
| PTR | L | MD | 2.445 | 0.015 | 0.219 | 0.089 |
| | R | MD | 1.629 | 0.104 | 0.146 | 0.089 |
| RLIC | L | MD | 0.106 | 0.915 | 0.010 | 0.089 |
| | R | MD | 1.019 | 0.308 | 0.091 | 0.089 |
| SCR | L | MD | 1.003 | 0.316 | 0.090 | 0.089 |
| | R | MD | 0.491 | 0.624 | 0.044 | 0.089 |
| SFO | L | MD | 0.147 | 0.883 | 0.013 | 0.089 |
| | R | MD | 0.331 | 0.741 | 0.030 | 0.089 |
| SLF | L | MD | 2.199 | 0.028 | 0.197 | 0.089 |
| | R | MD | 1.719 | 0.086 | 0.154 | 0.089 |
| SS | L | MD | 2.160 | 0.031 | 0.194 | 0.089 |
| | R | MD | 1.578 | 0.115 | 0.141 | 0.089 |
| UNC | L | MD | 3.226 | **0.001** | 0.289 | 0.089 |
| | R | MD | 2.445 | 0.015 | 0.219 | 0.089 |
| ACR | L | RD | 1.358 | 0.175 | 0.122 | 0.089 |
| | R | RD | 1.451 | 0.147 | 0.130 | 0.089 |
| ALIC | L | RD | 1.961 | 0.050 | 0.176 | 0.089 |
| | R | RD | 1.370 | 0.171 | 0.123 | 0.089 |
| CGC | L | RD | 3.245 | **0.001** | 0.291 | 0.089 |
| | R | RD | 2.511 | 0.012 | 0.225 | 0.089 |
| CGH | L | RD | -0.357 | 0.721 | -0.032 | 0.089 |
| | R | RD | 0.749 | 0.454 | 0.067 | 0.089 |
| CR | L | RD | 2.400 | 0.017 | 0.215 | 0.089 |
| | R | RD | 2.047 | 0.041 | 0.184 | 0.089 |
| CST | L | RD | 0.844 | 0.399 | 0.076 | 0.089 |
| | R | RD | 1.372 | 0.171 | 0.123 | 0.089 |

| | | | | | | |
|---|---|---|---|---|---|---|
| EC | L | RD | 2.296 | 0.022 | 0.206 | 0.089 |
| | R | RD | 1.483 | 0.139 | 0.133 | 0.089 |
| FXST | L | RD | 1.724 | 0.085 | 0.155 | 0.089 |
| | R | RD | 2.222 | 0.027 | 0.199 | 0.089 |
| IC | L | RD | 2.363 | 0.019 | 0.212 | 0.089 |
| | R | RD | 1.652 | 0.099 | 0.148 | 0.089 |
| IFO | L | RD | 1.734 | 0.084 | 0.155 | 0.089 |
| | R | RD | 1.340 | 0.181 | 0.120 | 0.089 |
| PCR | L | RD | 3.677 | **2.611e-04** | 0.330 | 0.089 |
| | R | RD | 3.038 | 0.003 | 0.272 | 0.089 |
| PLIC | L | RD | 1.403 | 0.161 | 0.126 | 0.089 |
| | R | RD | 0.883 | 0.378 | 0.079 | 0.089 |
| PTR | L | RD | 3.646 | **2.938e-04** | 0.327 | 0.089 |
| | R | RD | 2.155 | 0.032 | 0.193 | 0.089 |
| RLIC | L | RD | 2.490 | 0.013 | 0.223 | 0.089 |
| | R | RD | 1.932 | 0.054 | 0.173 | 0.089 |
| SCR | L | RD | 2.258 | 0.024 | 0.202 | 0.089 |
| | R | RD | 1.584 | 0.114 | 0.142 | 0.089 |
| SFO | L | RD | 1.852 | 0.065 | 0.166 | 0.089 |
| | R | RD | 2.145 | 0.032 | 0.192 | 0.089 |
| SLF | L | RD | 3.695 | **2.441e-04** | 0.331 | 0.089 |
| | R | RD | 3.758 | **1.913e-04** | 0.337 | 0.089 |
| SS | L | RD | 3.054 | **0.002** | 0.274 | 0.089 |
| | R | RD | 2.150 | 0.032 | 0.193 | 0.089 |
| UNC | L | RD | 2.699 | 0.007 | 0.242 | 0.089 |
| | R | RD | 3.123 | **0.002** | 0.280 | 0.089 |
| ACR | L | AD | -0.465 | 0.642 | -0.042 | 0.089 |
| | R | AD | -0.504 | 0.615 | -0.045 | 0.089 |
| ALIC | L | AD | -1.483 | 0.139 | -0.133 | 0.089 |
| | R | AD | -0.533 | 0.594 | -0.048 | 0.089 |
| CGC | L | AD | -1.170 | 0.243 | -0.105 | 0.089 |
| | R | AD | -0.607 | 0.544 | -0.054 | 0.089 |
| CGH | L | AD | -0.614 | 0.539 | -0.055 | 0.089 |
| | R | AD | -0.164 | 0.870 | -0.015 | 0.089 |
| CR | L | AD | -0.434 | 0.664 | -0.039 | 0.089 |
| | R | AD | -0.480 | 0.631 | -0.043 | 0.089 |
| CST | L | AD | -0.601 | 0.548 | -0.054 | 0.089 |
| | R | AD | -0.386 | 0.700 | -0.035 | 0.089 |
| EC | L | AD | 0.436 | 0.663 | 0.039 | 0.089 |
| | R | AD | 0.171 | 0.865 | 0.015 | 0.089 |
| FXST | L | AD | -1.843 | 0.066 | -0.165 | 0.089 |
| | R | AD | -0.110 | 0.912 | -0.010 | 0.089 |
| IC | L | AD | -2.280 | 0.023 | -0.204 | 0.089 |
| | R | AD | -0.870 | 0.385 | -0.078 | 0.089 |
| IFO | L | AD | 0.127 | 0.899 | 0.011 | 0.089 |
| | R | AD | -0.239 | 0.811 | -0.021 | 0.089 |

| | | | | | | |
|---|---|---|---|---|---|---|
| PCR | L | AD | 0.463 | 0.644 | 0.042 | 0.089 |
| | R | AD | -0.131 | 0.896 | -0.012 | 0.089 |
| PLIC | L | AD | -1.447 | 0.148 | -0.130 | 0.089 |
| | R | AD | -0.943 | 0.346 | -0.085 | 0.089 |
| PTR | L | AD | 0.049 | 0.961 | 0.004 | 0.089 |
| | R | AD | 0.326 | 0.744 | 0.029 | 0.089 |
| RLIC | L | AD | -2.262 | 0.024 | -0.203 | 0.089 |
| | R | AD | -0.386 | 0.700 | -0.035 | 0.089 |
| SCR | L | AD | -0.537 | 0.591 | -0.048 | 0.089 |
| | R | AD | -0.363 | 0.716 | -0.033 | 0.089 |
| SFO | L | AD | -1.258 | 0.209 | -0.113 | 0.089 |
| | R | AD | -1.365 | 0.173 | -0.122 | 0.089 |
| SLF | L | AD | -0.512 | 0.609 | -0.046 | 0.089 |
| | R | AD | -1.051 | 0.294 | -0.094 | 0.089 |
| SS | L | AD | 0.226 | 0.821 | 0.020 | 0.089 |
| | R | AD | 0.550 | 0.583 | 0.049 | 0.089 |
| UNC | L | AD | 2.903 | 0.004 | 0.260 | 0.089 |
| | R | AD | 0.814 | 0.416 | 0.073 | 0.089 |

Significant results are highlighted in bold ($p < 0.002$). Abbreviation: L = left, R = right, FA = fractional anisotropy, MD = mean diffusivity, RD = radial diffusivity, AD = axial diffusivity, S.E. = standard error. Abbreviations for tracts, see Table 1 in main text.

**Table S9| Multiple linear regression output for case-control differences in bilateral regional diffusion measures, stratified by sex.**

| Tract | contrast | metric | t-value | p-value | Cohen's *d* | S.E. |
|---|---|---|---|---|---|---|
| ACR | Females | FA | -0.255 | 0.799 | -0.030 | 0.115 |
| | Males | FA | -2.619 | 0.009 | -0.320 | 0.120 |
| | Females | MD | -0.761 | 0.447 | -0.094 | 0.124 |
| | Males | MD | 1.676 | 0.095 | 0.220 | 0.129 |
| | Females | RD | -0.235 | 0.814 | -0.029 | 0.124 |
| | Males | RD | 2.499 | 0.013 | 0.329 | 0.129 |
| | Females | AD | -0.996 | 0.320 | -0.124 | 0.124 |
| | Males | AD | 0.281 | 0.779 | 0.037 | 0.128 |
| ALIC | Females | FA | -1.713 | 0.088 | -0.198 | 0.116 |
| | Males | FA | -2.535 | 0.012 | -0.310 | 0.120 |
| | Females | MD | -0.346 | 0.730 | -0.043 | 0.124 |
| | Males | MD | 0.427 | 0.670 | 0.056 | 0.128 |
| | Females | RD | 0.982 | 0.327 | 0.122 | 0.124 |
| | Males | RD | 1.527 | 0.128 | 0.201 | 0.129 |
| | Females | AD | -1.267 | 0.206 | -0.157 | 0.124 |
| | Males | AD | -0.265 | 0.791 | -0.035 | 0.128 |
| Average | Females | FA | -0.701 | 0.484 | -0.081 | 0.116 |
| | Males | FA | -4.577 | **7.096e-06** | -0.560 | 0.121 |
| | Females | MD | 0.368 | 0.713 | 0.046 | 0.124 |
| | Males | MD | 2.293 | 0.023 | 0.302 | 0.129 |
| | Females | RD | 0.955 | 0.341 | 0.118 | 0.124 |
| | Males | RD | 3.760 | **2.135e-04** | 0.494 | 0.130 |
| | Females | AD | 0.493 | 0.623 | 0.061 | 0.124 |
| | Males | AD | 0.099 | 0.921 | 0.013 | 0.128 |
| BCC | Females | FA | -0.812 | 0.418 | -0.094 | 0.116 |
| | Males | FA | -3.376 | **0.001** | -0.413 | 0.120 |
| | Females | MD | -0.324 | 0.746 | -0.040 | 0.124 |
| | Males | MD | 0.625 | 0.532 | 0.082 | 0.128 |
| | Females | RD | 0.214 | 0.831 | 0.027 | 0.124 |
| | Males | RD | 2.930 | 0.004 | 0.385 | 0.129 |
| | Females | AD | -0.135 | 0.893 | -0.017 | 0.124 |
| | Males | AD | -2.215 | 0.028 | -0.291 | 0.129 |
| CC | Females | FA | -0.841 | 0.401 | -0.097 | 0.116 |
| | Males | FA | -3.999 | **8.152e-05** | -0.489 | 0.121 |
| | Females | MD | -0.899 | 0.370 | -0.111 | 0.124 |
| | Males | MD | 1.116 | 0.265 | 0.147 | 0.128 |
| | Females | RD | 0.064 | 0.949 | 0.008 | 0.124 |
| | Males | RD | 3.604 | **3.809e-04** | 0.474 | 0.130 |
| | Females | AD | -0.846 | 0.399 | -0.105 | 0.124 |
| | Males | AD | -1.682 | 0.094 | -0.221 | 0.129 |
| CGC | Females | FA | -0.419 | 0.676 | -0.049 | 0.115 |
| | Males | FA | -3.264 | **0.001** | -0.399 | 0.120 |
| | Females | MD | -0.049 | 0.961 | -0.006 | 0.124 |

| | | | | | | |
|---|---|---|---|---|---|---|
| | Males | MD | 0.909 | 0.364 | 0.120 | 0.128 |
| | Females | RD | 0.792 | 0.429 | 0.098 | 0.124 |
| | Males | RD | 3.657 | **3.137e-04** | 0.481 | 0.130 |
| | Females | AD | 0.012 | 0.990 | 0.002 | 0.124 |
| | Males | AD | -1.405 | 0.161 | -0.185 | 0.129 |
| CGH | Females | FA | 0.582 | 0.561 | 0.067 | 0.116 |
| | Males | FA | -1.871 | 0.062 | -0.229 | 0.119 |
| | Females | MD | -1.510 | 0.132 | -0.187 | 0.124 |
| | Males | MD | 0.711 | 0.478 | 0.093 | 0.128 |
| | Females | RD | -1.037 | 0.301 | -0.129 | 0.124 |
| | Males | RD | 1.486 | 0.139 | 0.195 | 0.129 |
| | Females | AD | -0.647 | 0.518 | -0.080 | 0.124 |
| | Males | AD | -0.134 | 0.894 | -0.018 | 0.128 |
| CR | Females | FA | -0.768 | 0.443 | -0.089 | 0.116 |
| | Males | FA | -3.022 | 0.003 | -0.370 | 0.120 |
| | Females | MD | -0.631 | 0.528 | -0.078 | 0.124 |
| | Males | MD | 2.286 | 0.023 | 0.301 | 0.129 |
| | Females | RD | 0.188 | 0.851 | 0.023 | 0.124 |
| | Males | RD | 3.283 | **0.001** | 0.432 | 0.130 |
| | Females | AD | -1.285 | 0.200 | -0.159 | 0.124 |
| | Males | AD | 0.589 | 0.556 | 0.077 | 0.128 |
| CST | Females | FA | 0.391 | 0.696 | 0.045 | 0.115 |
| | Males | FA | -1.206 | 0.229 | -0.147 | 0.119 |
| | Females | MD | -0.762 | 0.446 | -0.095 | 0.124 |
| | Males | MD | 1.190 | 0.235 | 0.156 | 0.128 |
| | Females | RD | -0.200 | 0.842 | -0.025 | 0.124 |
| | Males | RD | 1.880 | 0.061 | 0.247 | 0.129 |
| | Females | AD | -1.183 | 0.238 | -0.147 | 0.124 |
| | Males | AD | 0.431 | 0.667 | 0.057 | 0.128 |
| EC | Females | FA | -0.100 | 0.921 | -0.012 | 0.115 |
| | Males | FA | -3.191 | **0.002** | -0.390 | 0.120 |
| | Females | MD | 0.272 | 0.786 | 0.034 | 0.124 |
| | Males | MD | 1.394 | 0.165 | 0.183 | 0.129 |
| | Females | RD | 0.374 | 0.709 | 0.046 | 0.124 |
| | Males | RD | 2.600 | 0.010 | 0.342 | 0.129 |
| | Females | AD | 0.589 | 0.557 | 0.073 | 0.124 |
| | Males | AD | -0.191 | 0.849 | -0.025 | 0.128 |
| FX | Females | FA | 0.247 | 0.805 | 0.029 | 0.115 |
| | Males | FA | -3.242 | **0.001** | -0.396 | 0.120 |
| | Females | MD | 2.449 | 0.015 | 0.304 | 0.124 |
| | Males | MD | 3.015 | 0.003 | 0.396 | 0.130 |
| | Females | RD | 2.290 | 0.023 | 0.284 | 0.124 |
| | Males | RD | 3.186 | **0.002** | 0.419 | 0.130 |
| | Females | AD | 2.636 | 0.009 | 0.327 | 0.124 |
| | Males | AD | 2.672 | 0.008 | 0.351 | 0.129 |
| FXST | Females | FA | -0.624 | 0.533 | -0.072 | 0.116 |

| | | | | | | |
|---|---|---|---|---|---|---|
| | Males | FA | -3.581 | **4.04e-04** | -0.438 | 0.121 |
| | Females | MD | -1.334 | 0.183 | -0.165 | 0.124 |
| | Males | MD | 1.614 | 0.108 | 0.212 | 0.129 |
| | Females | RD | -0.464 | 0.643 | -0.058 | 0.124 |
| | Males | RD | 3.519 | **0.001** | 0.463 | 0.130 |
| | Females | AD | -0.993 | 0.321 | -0.123 | 0.124 |
| | Males | AD | -0.551 | 0.582 | -0.072 | 0.128 |
| GCC | Females | FA | -1.190 | 0.235 | -0.138 | 0.116 |
| | Males | FA | -3.709 | **2.507e-04** | -0.454 | 0.121 |
| | Females | MD | -0.899 | 0.369 | -0.112 | 0.124 |
| | Males | MD | 0.975 | 0.331 | 0.128 | 0.128 |
| | Females | RD | 0.377 | 0.707 | 0.047 | 0.124 |
| | Males | RD | 3.033 | 0.003 | 0.399 | 0.130 |
| | Females | AD | -1.495 | 0.136 | -0.185 | 0.124 |
| | Males | AD | -0.956 | 0.340 | -0.126 | 0.128 |
| IC | Females | FA | -2.048 | 0.041 | -0.237 | 0.116 |
| | Males | FA | -2.912 | 0.004 | -0.356 | 0.120 |
| | Females | MD | -1.032 | 0.303 | -0.128 | 0.124 |
| | Males | MD | 1.195 | 0.233 | 0.157 | 0.129 |
| | Females | RD | 0.602 | 0.548 | 0.075 | 0.124 |
| | Males | RD | 2.441 | 0.015 | 0.321 | 0.129 |
| | Females | AD | -2.276 | 0.024 | -0.282 | 0.124 |
| | Males | AD | -0.135 | 0.893 | -0.018 | 0.128 |
| IFO | Females | FA | 0.078 | 0.938 | 0.009 | 0.115 |
| | Males | FA | -2.163 | 0.031 | -0.264 | 0.120 |
| | Females | MD | 0.378 | 0.706 | 0.047 | 0.124 |
| | Males | MD | 1.507 | 0.133 | 0.198 | 0.129 |
| | Females | RD | 0.332 | 0.740 | 0.041 | 0.124 |
| | Males | RD | 2.397 | 0.017 | 0.315 | 0.129 |
| | Females | AD | 0.286 | 0.775 | 0.035 | 0.124 |
| | Males | AD | -0.624 | 0.533 | -0.082 | 0.128 |
| PCR | Females | FA | -2.212 | 0.028 | -0.256 | 0.116 |
| | Males | FA | -3.213 | **0.001** | -0.393 | 0.120 |
| | Females | MD | 0.159 | 0.874 | 0.020 | 0.124 |
| | Males | MD | 3.368 | **0.001** | 0.443 | 0.130 |
| | Females | RD | 1.244 | 0.215 | 0.154 | 0.124 |
| | Males | RD | 3.758 | **2.149e-04** | 0.494 | 0.130 |
| | Females | AD | -1.157 | 0.249 | -0.143 | 0.124 |
| | Males | AD | 1.356 | 0.176 | 0.178 | 0.129 |
| PLIC | Females | FA | -1.569 | 0.118 | -0.182 | 0.116 |
| | Males | FA | -1.754 | 0.081 | -0.214 | 0.119 |
| | Females | MD | -1.392 | 0.165 | -0.173 | 0.124 |
| | Males | MD | 0.932 | 0.352 | 0.123 | 0.128 |
| | Females | RD | 0.274 | 0.784 | 0.034 | 0.124 |
| | Males | RD | 1.468 | 0.143 | 0.193 | 0.129 |
| | Females | AD | -2.169 | 0.031 | -0.269 | 0.124 |

| | | | | | | |
|---|---|---|---|---|---|---|
| | Males | AD | 0.207 | 0.836 | 0.027 | 0.128 |
| PTR | Females | FA | -1.777 | 0.077 | -0.206 | 0.116 |
| | Males | FA | -2.640 | 0.009 | -0.323 | 0.120 |
| | Females | MD | 0.152 | 0.879 | 0.019 | 0.124 |
| | Males | MD | 3.013 | 0.003 | 0.396 | 0.130 |
| | Females | RD | 1.121 | 0.263 | 0.139 | 0.124 |
| | Males | RD | 3.312 | **0.001** | 0.435 | 0.130 |
| | Females | AD | -0.975 | 0.330 | -0.121 | 0.124 |
| | Males | AD | 1.274 | 0.204 | 0.167 | 0.129 |
| RLIC | Females | FA | -1.639 | 0.102 | -0.190 | 0.116 |
| | Males | FA | -2.891 | 0.004 | -0.354 | 0.120 |
| | Females | MD | -0.818 | 0.414 | -0.102 | 0.124 |
| | Males | MD | 1.842 | 0.067 | 0.242 | 0.129 |
| | Females | RD | 0.267 | 0.790 | 0.033 | 0.124 |
| | Males | RD | 3.167 | **0.002** | 0.416 | 0.130 |
| | Females | AD | -1.567 | 0.118 | -0.194 | 0.124 |
| | Males | AD | -0.429 | 0.668 | -0.056 | 0.128 |
| SCC | Females | FA | -0.162 | 0.871 | -0.019 | 0.115 |
| | Males | FA | -3.222 | **0.001** | -0.394 | 0.120 |
| | Females | MD | -1.138 | 0.256 | -0.141 | 0.124 |
| | Males | MD | 1.865 | 0.063 | 0.245 | 0.129 |
| | Females | RD | -0.769 | 0.443 | -0.095 | 0.124 |
| | Males | RD | 3.558 | **4.506e-04** | 0.468 | 0.130 |
| | Females | AD | -0.480 | 0.632 | -0.060 | 0.124 |
| | Males | AD | -0.715 | 0.475 | -0.094 | 0.128 |
| SCR | Females | FA | -0.393 | 0.695 | -0.046 | 0.115 |
| | Males | FA | -2.088 | 0.038 | -0.255 | 0.120 |
| | Females | MD | -0.825 | 0.410 | -0.102 | 0.124 |
| | Males | MD | 1.982 | 0.049 | 0.261 | 0.129 |
| | Females | RD | 0.032 | 0.975 | 0.004 | 0.124 |
| | Males | RD | 2.892 | 0.004 | 0.380 | 0.129 |
| | Females | AD | -1.126 | 0.261 | -0.140 | 0.124 |
| | Males | AD | 0.381 | 0.703 | 0.050 | 0.128 |
| SFO | Females | FA | -1.719 | 0.087 | -0.199 | 0.116 |
| | Males | FA | -3.664 | **2.966e-04** | -0.448 | 0.121 |
| | Females | MD | -0.709 | 0.479 | -0.088 | 0.124 |
| | Males | MD | 1.129 | 0.260 | 0.148 | 0.128 |
| | Females | RD | 0.786 | 0.433 | 0.097 | 0.124 |
| | Males | RD | 2.419 | 0.016 | 0.318 | 0.129 |
| | Females | AD | -1.620 | 0.107 | -0.201 | 0.124 |
| | Males | AD | -0.421 | 0.674 | -0.055 | 0.128 |
| SLF | Females | FA | -1.818 | 0.070 | -0.211 | 0.116 |
| | Males | FA | -4.616 | **5.954e-06** | -0.565 | 0.121 |
| | Females | MD | 0.240 | 0.811 | 0.030 | 0.124 |
| | Males | MD | 2.892 | 0.004 | 0.380 | 0.129 |
| | Females | RD | 1.408 | 0.160 | 0.175 | 0.124 |

| | | | | | | |
|---|---|---|---|---|---|---|
| | Males | RD | 4.456 | **1.279e-05** | 0.586 | 0.131 |
| | Females | AD | -1.146 | 0.253 | -0.142 | 0.124 |
| | Males | AD | -0.095 | 0.924 | -0.013 | 0.128 |
| SS | Females | FA | -0.252 | 0.802 | -0.029 | 0.115 |
| | Males | FA | -2.245 | 0.026 | -0.274 | 0.120 |
| | Females | MD | 0.183 | 0.855 | 0.023 | 0.124 |
| | Males | MD | 2.732 | 0.007 | 0.359 | 0.129 |
| | Females | RD | 0.567 | 0.571 | 0.070 | 0.124 |
| | Males | RD | 3.560 | **4.469e-04** | 0.468 | 0.130 |
| | Females | AD | -0.038 | 0.970 | -0.005 | 0.124 |
| | Males | AD | 0.577 | 0.565 | 0.076 | 0.128 |
| UNC | Females | FA | -0.633 | 0.527 | -0.073 | 0.116 |
| | Males | FA | -2.516 | 0.012 | -0.308 | 0.120 |
| | Females | MD | 1.623 | 0.106 | 0.201 | 0.124 |
| | Males | MD | 3.073 | **0.002** | 0.404 | 0.130 |
| | Females | RD | 1.289 | 0.198 | 0.160 | 0.124 |
| | Males | RD | 3.457 | **0.001** | 0.455 | 0.130 |
| | Females | AD | 1.884 | 0.061 | 0.234 | 0.124 |
| | Males | AD | 1.058 | 0.291 | 0.139 | 0.128 |

Significant results are highlighted in bold ($p < 0.002$). Abbreviation: FA = fractional anisotropy, MD = mean diffusivity, RD = radial diffusivity, AD = axial diffusivity, S.E. = standard error. Abbreviations for tracts, see Table 1 in main text.

**Table S10| Multiple linear regression output for diagnostic subgroup differences relative to healthy controls in bilateral regional diffusion measures.**

| Tract | Group | metric | t-value | p-value | Cohen's *d* | S.E. |
|---|---|---|---|---|---|---|
| ACR | EOS | FA | -1.895 | 0.059 | -0.161 | 0.095 |
| ALIC | EOS | FA | -3.133 | **0.002** | -0.266 | 0.095 |
| AverageFA | EOS | FA | -3.926 | **9.677e-05** | -0.333 | 0.096 |
| BCC | EOS | FA | -3.334 | **0.001** | -0.283 | 0.095 |
| CC | EOS | FA | -4.225 | **2.776e-05** | -0.358 | 0.096 |
| CGC | EOS | FA | -2.654 | 0.008 | -0.225 | 0.095 |
| CGH | EOS | FA | -1.194 | 0.233 | -0.101 | 0.095 |
| CR | EOS | FA | -3.125 | **0.002** | -0.265 | 0.095 |
| CST | EOS | FA | -0.701 | 0.483 | -0.059 | 0.095 |
| EC | EOS | FA | -2.934 | 0.003 | -0.249 | 0.095 |
| FX | EOS | FA | -2.369 | 0.018 | -0.201 | 0.095 |
| FXST | EOS | FA | -3.293 | **0.001** | -0.279 | 0.095 |
| GCC | EOS | FA | -4.025 | **6.448e-05** | -0.341 | 0.096 |
| IC | EOS | FA | -4.020 | **6.604e-05** | -0.341 | 0.096 |
| IFO | EOS | FA | -1.491 | 0.136 | -0.126 | 0.095 |
| PCR | EOS | FA | -4.413 | **1.215e-05** | -0.374 | 0.096 |
| PLIC | EOS | FA | -2.801 | 0.005 | -0.238 | 0.095 |
| PTR | EOS | FA | -3.605 | **3.392e-04** | -0.306 | 0.096 |
| RLIC | EOS | FA | -3.766 | **1.828e-04** | -0.319 | 0.096 |
| SCC | EOS | FA | -3.589 | **3.6e-04** | -0.304 | 0.096 |
| SCR | EOS | FA | -2.579 | 0.010 | -0.219 | 0.095 |
| SFO | EOS | FA | -3.638 | **2.999e-04** | -0.309 | 0.096 |
| SLF | EOS | FA | -4.753 | **2.535e-06** | -0.403 | 0.096 |
| SS | EOS | FA | -2.115 | 0.035 | -0.179 | 0.095 |
| UNC | EOS | FA | -2.608 | 0.009 | -0.221 | 0.095 |
| ACR | EOS | MD | 0.777 | 0.437 | 0.072 | 0.105 |
| ALIC | EOS | MD | 0.710 | 0.478 | 0.066 | 0.105 |
| AverageMD | EOS | MD | 1.371 | 0.171 | 0.127 | 0.105 |
| BCC | EOS | MD | 0.124 | 0.902 | 0.011 | 0.105 |
| CC | EOS | MD | 0.410 | 0.682 | 0.038 | 0.105 |
| CGC | EOS | MD | 0.689 | 0.491 | 0.064 | 0.105 |
| CGH | EOS | MD | 0.143 | 0.886 | 0.013 | 0.105 |
| CR | EOS | MD | 1.351 | 0.177 | 0.125 | 0.105 |
| CST | EOS | MD | 0.710 | 0.478 | 0.066 | 0.105 |
| EC | EOS | MD | 1.860 | 0.064 | 0.172 | 0.105 |
| FX | EOS | MD | 4.152 | **3.88e-05** | 0.384 | 0.106 |
| FXST | EOS | MD | 0.490 | 0.624 | 0.045 | 0.105 |
| GCC | EOS | MD | 0.613 | 0.540 | 0.057 | 0.105 |
| IC | EOS | MD | 0.995 | 0.320 | 0.092 | 0.105 |
| IFO | EOS | MD | 1.366 | 0.173 | 0.126 | 0.105 |
| PCR | EOS | MD | 2.562 | 0.011 | 0.237 | 0.105 |
| PLIC | EOS | MD | 0.469 | 0.639 | 0.043 | 0.105 |
| PTR | EOS | MD | 2.613 | 0.009 | 0.242 | 0.105 |

| | | | | | | |
|---|---|---|---|---|---|---|
| RLIC | EOS | MD | 1.616 | 0.107 | 0.150 | 0.105 |
| SCC | EOS | MD | 1.016 | 0.310 | 0.094 | 0.105 |
| SCR | EOS | MD | 1.084 | 0.279 | 0.100 | 0.105 |
| SFO | EOS | MD | 1.084 | 0.279 | 0.100 | 0.105 |
| SLF | EOS | MD | 2.633 | 0.009 | 0.244 | 0.105 |
| SS | EOS | MD | 2.767 | 0.006 | 0.256 | 0.105 |
| UNC | EOS | MD | 2.641 | 0.009 | 0.244 | 0.105 |
| ACR | EOS | RD | 1.464 | 0.144 | 0.135 | 0.105 |
| ALIC | EOS | RD | 2.103 | 0.036 | 0.195 | 0.105 |
| AverageRD | EOS | RD | 2.596 | 0.010 | 0.240 | 0.105 |
| BCC | EOS | RD | 2.465 | 0.014 | 0.228 | 0.105 |
| CC | EOS | RD | 3.106 | **0.002** | 0.287 | 0.105 |
| CGC | EOS | RD | 3.195 | **0.001** | 0.296 | 0.105 |
| CGH | EOS | RD | 0.822 | 0.412 | 0.076 | 0.105 |
| CR | EOS | RD | 2.643 | 0.008 | 0.244 | 0.105 |
| CST | EOS | RD | 1.340 | 0.181 | 0.124 | 0.105 |
| EC | EOS | RD | 2.805 | 0.005 | 0.259 | 0.105 |
| FX | EOS | RD | 4.231 | **2.769e-05** | 0.391 | 0.106 |
| FXST | EOS | RD | 2.513 | 0.012 | 0.233 | 0.105 |
| GCC | EOS | RD | 2.841 | 0.005 | 0.263 | 0.105 |
| IC | EOS | RD | 2.749 | 0.006 | 0.254 | 0.105 |
| IFO | EOS | RD | 1.613 | 0.107 | 0.149 | 0.105 |
| PCR | EOS | RD | 3.908 | **1.061e-04** | 0.361 | 0.105 |
| PLIC | EOS | RD | 1.719 | 0.086 | 0.159 | 0.105 |
| PTR | EOS | RD | 3.489 | **0.001** | 0.323 | 0.105 |
| RLIC | EOS | RD | 3.296 | **0.001** | 0.305 | 0.105 |
| SCC | EOS | RD | 2.999 | 0.003 | 0.277 | 0.105 |
| SCR | EOS | RD | 2.602 | 0.010 | 0.241 | 0.105 |
| SFO | EOS | RD | 2.462 | 0.014 | 0.228 | 0.105 |
| SLF | EOS | RD | 4.398 | **1.337e-05** | 0.407 | 0.106 |
| SS | EOS | RD | 3.311 | **0.001** | 0.306 | 0.105 |
| UNC | EOS | RD | 2.990 | 0.003 | 0.277 | 0.105 |
| ACR | EOS | AD | -0.184 | 0.854 | -0.017 | 0.105 |
| ALIC | EOS | AD | -0.524 | 0.600 | -0.048 | 0.105 |
| AverageAD | EOS | AD | 0.035 | 0.972 | 0.003 | 0.105 |
| BCC | EOS | AD | -2.427 | 0.016 | -0.225 | 0.105 |
| CC | EOS | AD | -2.173 | 0.030 | -0.201 | 0.105 |
| CGC | EOS | AD | -1.069 | 0.285 | -0.099 | 0.105 |
| CGH | EOS | AD | 0.154 | 0.878 | 0.014 | 0.105 |
| CR | EOS | AD | -0.548 | 0.584 | -0.051 | 0.105 |
| CST | EOS | AD | 0.292 | 0.770 | 0.027 | 0.105 |
| EC | EOS | AD | 0.337 | 0.736 | 0.031 | 0.105 |
| FX | EOS | AD | 3.772 | **1.813e-04** | 0.349 | 0.105 |
| FXST | EOS | AD | -1.281 | 0.201 | -0.119 | 0.105 |
| GCC | EOS | AD | -1.550 | 0.122 | -0.143 | 0.105 |
| IC | EOS | AD | -0.985 | 0.325 | -0.091 | 0.105 |

| | | | | | | |
|---|---|---|---|---|---|---|
| IFO | EOS | AD | 0.221 | 0.825 | 0.020 | 0.105 |
| PCR | EOS | AD | -0.239 | 0.811 | -0.022 | 0.105 |
| PLIC | EOS | AD | -0.786 | 0.432 | -0.073 | 0.105 |
| PTR | EOS | AD | 0.371 | 0.711 | 0.034 | 0.105 |
| RLIC | EOS | AD | -0.891 | 0.373 | -0.082 | 0.105 |
| SCC | EOS | AD | -0.959 | 0.338 | -0.089 | 0.105 |
| SCR | EOS | AD | -0.875 | 0.382 | -0.081 | 0.105 |
| SFO | EOS | AD | -0.732 | 0.464 | -0.068 | 0.105 |
| SLF | EOS | AD | -0.459 | 0.646 | -0.042 | 0.105 |
| SS | EOS | AD | 1.040 | 0.299 | 0.096 | 0.105 |
| UNC | EOS | AD | 1.186 | 0.236 | 0.110 | 0.105 |
| ACR | AFP | FA | -0.921 | 0.357 | -0.087 | 0.106 |
| ALIC | AFP | FA | -1.328 | 0.185 | -0.125 | 0.106 |
| AverageFA | AFP | FA | -1.826 | 0.068 | -0.172 | 0.106 |
| BCC | AFP | FA | -1.418 | 0.157 | -0.134 | 0.106 |
| CC | AFP | FA | -1.055 | 0.292 | -0.100 | 0.106 |
| CGC | AFP | FA | -1.115 | 0.265 | -0.105 | 0.106 |
| CGH | AFP | FA | -0.394 | 0.693 | -0.037 | 0.106 |
| CR | AFP | FA | -0.940 | 0.348 | -0.089 | 0.106 |
| CST | AFP | FA | 0.401 | 0.688 | 0.038 | 0.106 |
| EC | AFP | FA | -0.201 | 0.841 | -0.019 | 0.106 |
| FX | AFP | FA | -0.903 | 0.367 | -0.085 | 0.106 |
| FXST | AFP | FA | -1.211 | 0.226 | -0.114 | 0.106 |
| GCC | AFP | FA | -1.250 | 0.212 | -0.118 | 0.106 |
| IC | AFP | FA | -1.249 | 0.212 | -0.118 | 0.106 |
| IFO | AFP | FA | -0.620 | 0.535 | -0.059 | 0.106 |
| PCR | AFP | FA | -1.716 | 0.087 | -0.162 | 0.106 |
| PLIC | AFP | FA | -0.362 | 0.718 | -0.034 | 0.106 |
| PTR | AFP | FA | -1.283 | 0.200 | -0.121 | 0.106 |
| RLIC | AFP | FA | -1.549 | 0.122 | -0.146 | 0.106 |
| SCC | AFP | FA | -0.012 | 0.990 | -0.001 | 0.106 |
| SCR | AFP | FA | -0.220 | 0.826 | -0.021 | 0.106 |
| SFO | AFP | FA | -2.195 | 0.029 | -0.207 | 0.106 |
| SLF | AFP | FA | -2.150 | 0.032 | -0.203 | 0.106 |
| SS | AFP | FA | -0.664 | 0.507 | -0.063 | 0.106 |
| UNC | AFP | FA | -1.184 | 0.237 | -0.112 | 0.106 |
| ACR | AFP | MD | 0.280 | 0.779 | 0.028 | 0.111 |
| ALIC | AFP | MD | -0.612 | 0.541 | -0.060 | 0.111 |
| AverageMD | AFP | MD | 2.437 | 0.015 | 0.240 | 0.112 |
| BCC | AFP | MD | 0.632 | 0.528 | 0.062 | 0.111 |
| CC | AFP | MD | -0.029 | 0.977 | -0.003 | 0.111 |
| CGC | AFP | MD | 0.167 | 0.868 | 0.016 | 0.111 |
| CGH | AFP | MD | -1.245 | 0.214 | -0.122 | 0.111 |
| CR | AFP | MD | 0.581 | 0.562 | 0.057 | 0.111 |
| CST | AFP | MD | -0.170 | 0.865 | -0.017 | 0.111 |

| | | | | | | |
|---|---|---|---|---|---|---|
| EC | AFP | MD | -0.077 | 0.938 | -0.008 | 0.111 |
| FX | AFP | MD | 2.232 | 0.026 | 0.220 | 0.111 |
| FXST | AFP | MD | -0.255 | 0.798 | -0.025 | 0.111 |
| GCC | AFP | MD | -0.573 | 0.567 | -0.056 | 0.111 |
| IC | AFP | MD | -0.859 | 0.391 | -0.085 | 0.111 |
| IFO | AFP | MD | 0.597 | 0.551 | 0.059 | 0.111 |
| PCR | AFP | MD | 1.463 | 0.144 | 0.144 | 0.111 |
| PLIC | AFP | MD | -1.308 | 0.192 | -0.129 | 0.111 |
| PTR | AFP | MD | 0.978 | 0.328 | 0.096 | 0.111 |
| RLIC | AFP | MD | -0.338 | 0.736 | -0.033 | 0.111 |
| SCC | AFP | MD | -0.617 | 0.538 | -0.061 | 0.111 |
| SCR | AFP | MD | 0.376 | 0.707 | 0.037 | 0.111 |
| SFO | AFP | MD | 0.016 | 0.987 | 0.002 | 0.111 |
| SLF | AFP | MD | 0.961 | 0.337 | 0.095 | 0.111 |
| SS | AFP | MD | 0.503 | 0.615 | 0.050 | 0.111 |
| UNC | AFP | MD | 2.840 | 0.005 | 0.279 | 0.112 |
| ACR | AFP | RD | 0.970 | 0.332 | 0.095 | 0.111 |
| ALIC | AFP | RD | 0.691 | 0.490 | 0.068 | 0.111 |
| AverageRD | AFP | RD | 3.321 | **0.001** | 0.327 | 0.112 |
| BCC | AFP | RD | 1.488 | 0.137 | 0.146 | 0.111 |
| CC | AFP | RD | 1.196 | 0.232 | 0.118 | 0.111 |
| CGC | AFP | RD | 1.686 | 0.093 | 0.166 | 0.111 |
| CGH | AFP | RD | -0.210 | 0.834 | -0.021 | 0.111 |
| CR | AFP | RD | 1.298 | 0.195 | 0.128 | 0.111 |
| CST | AFP | RD | 0.519 | 0.604 | 0.051 | 0.111 |
| EC | AFP | RD | 0.232 | 0.816 | 0.023 | 0.111 |
| FX | AFP | RD | 2.160 | 0.031 | 0.213 | 0.111 |
| FXST | AFP | RD | 0.802 | 0.423 | 0.079 | 0.111 |
| GCC | AFP | RD | 1.066 | 0.287 | 0.105 | 0.111 |
| IC | AFP | RD | 0.558 | 0.577 | 0.055 | 0.111 |
| IFO | AFP | RD | 1.046 | 0.296 | 0.103 | 0.111 |
| PCR | AFP | RD | 2.012 | 0.045 | 0.198 | 0.111 |
| PLIC | AFP | RD | -0.190 | 0.849 | -0.019 | 0.111 |
| PTR | AFP | RD | 1.532 | 0.126 | 0.151 | 0.111 |
| RLIC | AFP | RD | 0.866 | 0.387 | 0.085 | 0.111 |
| SCC | AFP | RD | -0.032 | 0.975 | -0.003 | 0.111 |
| SCR | AFP | RD | 0.954 | 0.341 | 0.094 | 0.111 |
| SFO | AFP | RD | 1.493 | 0.136 | 0.147 | 0.111 |
| SLF | AFP | RD | 2.166 | 0.031 | 0.213 | 0.111 |
| SS | AFP | RD | 1.272 | 0.204 | 0.125 | 0.111 |
| UNC | AFP | RD | 2.677 | 0.008 | 0.263 | 0.112 |
| ACR | AFP | AD | -0.489 | 0.625 | -0.048 | 0.111 |
| ALIC | AFP | AD | -1.228 | 0.220 | -0.121 | 0.111 |
| AverageAD | AFP | AD | 1.287 | 0.199 | 0.127 | 0.111 |
| BCC | AFP | AD | -0.439 | 0.661 | -0.043 | 0.111 |
| CC | AFP | AD | -1.061 | 0.289 | -0.104 | 0.111 |

| | | | | | | |
|---|---|---|---|---|---|---|
| CGC | AFP | AD | -0.717 | 0.474 | -0.071 | 0.111 |
| CGH | AFP | AD | -1.504 | 0.133 | -0.148 | 0.111 |
| CR | AFP | AD | -0.230 | 0.818 | -0.023 | 0.111 |
| CST | AFP | AD | -0.990 | 0.323 | -0.097 | 0.111 |
| EC | AFP | AD | 0.413 | 0.680 | 0.041 | 0.111 |
| FX | AFP | AD | 2.481 | 0.013 | 0.244 | 0.112 |
| FXST | AFP | AD | -0.786 | 0.432 | -0.077 | 0.111 |
| GCC | AFP | AD | -1.459 | 0.145 | -0.144 | 0.111 |
| IC | AFP | AD | -1.691 | 0.091 | -0.166 | 0.111 |
| IFO | AFP | AD | -0.394 | 0.693 | -0.039 | 0.111 |
| PCR | AFP | AD | 0.218 | 0.828 | 0.021 | 0.111 |
| PLIC | AFP | AD | -1.166 | 0.244 | -0.115 | 0.111 |
| PTR | AFP | AD | 0.020 | 0.984 | 0.002 | 0.111 |
| RLIC | AFP | AD | -1.580 | 0.115 | -0.156 | 0.111 |
| SCC | AFP | AD | -0.950 | 0.343 | -0.094 | 0.111 |
| SCR | AFP | AD | -0.057 | 0.955 | -0.006 | 0.111 |
| SFO | AFP | AD | -1.272 | 0.204 | -0.125 | 0.111 |
| SLF | AFP | AD | -0.847 | 0.398 | -0.083 | 0.111 |
| SS | AFP | AD | -0.541 | 0.589 | -0.053 | 0.111 |
| UNC | AFP | AD | 2.192 | 0.029 | 0.216 | 0.111 |
| ACR | OTP | FA | -0.978 | 0.328 | -0.115 | 0.114 |
| ALIC | OTP | FA | -1.425 | 0.155 | -0.167 | 0.114 |
| AverageFA | OTP | FA | -1.056 | 0.291 | -0.124 | 0.114 |
| BCC | OTP | FA | -0.311 | 0.756 | -0.036 | 0.114 |
| CC | OTP | FA | -0.556 | 0.578 | -0.065 | 0.114 |
| CGC | OTP | FA | -0.929 | 0.353 | -0.109 | 0.114 |
| CGH | OTP | FA | 0.067 | 0.947 | 0.008 | 0.114 |
| CR | OTP | FA | -0.729 | 0.466 | -0.085 | 0.114 |
| CST | OTP | FA | -1.007 | 0.315 | -0.118 | 0.114 |
| EC | OTP | FA | -0.977 | 0.329 | -0.115 | 0.114 |
| FX | OTP | FA | -0.669 | 0.504 | -0.078 | 0.114 |
| FXST | OTP | FA | -0.870 | 0.385 | -0.102 | 0.114 |
| GCC | OTP | FA | -0.825 | 0.410 | -0.097 | 0.114 |
| IC | OTP | FA | -1.369 | 0.171 | -0.161 | 0.114 |
| IFO | OTP | FA | -0.723 | 0.470 | -0.085 | 0.114 |
| PCR | OTP | FA | -0.927 | 0.354 | -0.109 | 0.114 |
| PLIC | OTP | FA | -1.411 | 0.159 | -0.166 | 0.114 |
| PTR | OTP | FA | -0.905 | 0.366 | -0.106 | 0.114 |
| RLIC | OTP | FA | -0.455 | 0.649 | -0.053 | 0.114 |
| SCC | OTP | FA | -0.556 | 0.578 | -0.065 | 0.114 |
| SCR | OTP | FA | 0.000 | 1.000 | 0.000 | 0.114 |
| SFO | OTP | FA | -1.529 | 0.127 | -0.179 | 0.114 |
| SLF | OTP | FA | -1.600 | 0.110 | -0.188 | 0.114 |
| SS | OTP | FA | -0.383 | 0.702 | -0.045 | 0.114 |
| UNC | OTP | FA | -0.214 | 0.831 | -0.025 | 0.114 |
| ACR | OTP | MD | -0.273 | 0.785 | -0.033 | 0.121 |

| | | | | | | |
|---|---|---|---|---|---|---|
| ALIC | OTP | MD | -0.298 | 0.766 | -0.036 | 0.121 |
| AverageMD | OTP | MD | -0.278 | 0.781 | -0.033 | 0.121 |
| BCC | OTP | MD | -0.535 | 0.593 | -0.064 | 0.121 |
| CC | OTP | MD | -0.328 | 0.743 | -0.039 | 0.121 |
| CGC | OTP | MD | 0.464 | 0.643 | 0.056 | 0.121 |
| CGH | OTP | MD | -0.345 | 0.730 | -0.042 | 0.121 |
| CR | OTP | MD | 0.022 | 0.982 | 0.003 | 0.121 |
| CST | OTP | MD | -0.300 | 0.764 | -0.036 | 0.121 |
| EC | OTP | MD | 0.357 | 0.721 | 0.043 | 0.121 |
| FX | OTP | MD | 1.282 | 0.200 | 0.154 | 0.121 |
| FXST | OTP | MD | 0.247 | 0.805 | 0.030 | 0.121 |
| GCC | OTP | MD | -0.169 | 0.866 | -0.020 | 0.121 |
| IC | OTP | MD | -0.326 | 0.744 | -0.039 | 0.121 |
| IFO | OTP | MD | 0.670 | 0.503 | 0.080 | 0.121 |
| PCR | OTP | MD | 0.568 | 0.571 | 0.068 | 0.121 |
| PLIC | OTP | MD | -0.230 | 0.818 | -0.028 | 0.121 |
| PTR | OTP | MD | 0.556 | 0.578 | 0.067 | 0.121 |
| RLIC | OTP | MD | -0.301 | 0.763 | -0.036 | 0.121 |
| SCC | OTP | MD | 0.569 | 0.570 | 0.068 | 0.121 |
| SCR | OTP | MD | -0.036 | 0.971 | -0.004 | 0.121 |
| SFO | OTP | MD | -1.036 | 0.301 | -0.125 | 0.121 |
| SLF | OTP | MD | 0.376 | 0.707 | 0.045 | 0.121 |
| SS | OTP | MD | 0.529 | 0.597 | 0.064 | 0.121 |
| UNC | OTP | MD | 1.175 | 0.241 | 0.141 | 0.121 |
| ACR | OTP | RD | 0.339 | 0.735 | 0.041 | 0.121 |
| ALIC | OTP | RD | 0.662 | 0.509 | 0.080 | 0.121 |
| AverageRD | OTP | RD | 0.663 | 0.508 | 0.080 | 0.121 |
| BCC | OTP | RD | -0.072 | 0.942 | -0.009 | 0.121 |
| CC | OTP | RD | 0.284 | 0.776 | 0.034 | 0.121 |
| CGC | OTP | RD | 1.318 | 0.188 | 0.158 | 0.121 |
| CGH | OTP | RD | -0.334 | 0.738 | -0.040 | 0.121 |
| CR | OTP | RD | 0.516 | 0.606 | 0.062 | 0.121 |
| CST | OTP | RD | 0.320 | 0.749 | 0.038 | 0.121 |
| EC | OTP | RD | 0.996 | 0.320 | 0.120 | 0.121 |
| FX | OTP | RD | 1.307 | 0.192 | 0.157 | 0.121 |
| FXST | OTP | RD | 0.754 | 0.451 | 0.091 | 0.121 |
| GCC | OTP | RD | 0.555 | 0.579 | 0.067 | 0.121 |
| IC | OTP | RD | 0.762 | 0.447 | 0.092 | 0.121 |
| IFO | OTP | RD | 1.150 | 0.251 | 0.138 | 0.121 |
| PCR | OTP | RD | 0.765 | 0.445 | 0.092 | 0.121 |
| PLIC | OTP | RD | 0.905 | 0.366 | 0.109 | 0.121 |
| PTR | OTP | RD | 1.060 | 0.290 | 0.127 | 0.121 |
| RLIC | OTP | RD | 0.242 | 0.809 | 0.029 | 0.121 |
| SCC | OTP | RD | 0.779 | 0.436 | 0.094 | 0.121 |
| SCR | OTP | RD | 0.230 | 0.818 | 0.028 | 0.121 |

| | | | | | | |
|---|---|---|---|---|---|---|
| SFO | OTP | RD | 0.268 | 0.789 | 0.032 | 0.121 |
| SLF | OTP | RD | 1.260 | 0.208 | 0.151 | 0.121 |
| SS | OTP | RD | 0.940 | 0.348 | 0.113 | 0.121 |
| UNC | OTP | RD | 0.920 | 0.358 | 0.111 | 0.121 |
| ACR | OTP | AD | -0.663 | 0.508 | -0.080 | 0.121 |
| ALIC | OTP | AD | -0.766 | 0.444 | -0.092 | 0.121 |
| AverageAD | OTP | AD | -0.663 | 0.508 | -0.080 | 0.121 |
| BCC | OTP | AD | -0.252 | 0.801 | -0.030 | 0.121 |
| CC | OTP | AD | -0.103 | 0.918 | -0.012 | 0.121 |
| CGC | OTP | AD | -0.102 | 0.919 | -0.012 | 0.121 |
| CGH | OTP | AD | 0.209 | 0.834 | 0.025 | 0.121 |
| CR | OTP | AD | -0.207 | 0.836 | -0.025 | 0.121 |
| CST | OTP | AD | -1.015 | 0.311 | -0.122 | 0.121 |
| EC | OTP | AD | -0.346 | 0.729 | -0.042 | 0.121 |
| FX | OTP | AD | 1.223 | 0.222 | 0.147 | 0.121 |
| FXST | OTP | AD | 0.129 | 0.898 | 0.015 | 0.121 |
| GCC | OTP | AD | -0.432 | 0.666 | -0.052 | 0.121 |
| IC | OTP | AD | -0.971 | 0.332 | -0.117 | 0.121 |
| IFO | OTP | AD | -0.647 | 0.518 | -0.078 | 0.121 |
| PCR | OTP | AD | 0.492 | 0.623 | 0.059 | 0.121 |
| PLIC | OTP | AD | -0.914 | 0.361 | -0.110 | 0.121 |
| PTR | OTP | AD | -0.211 | 0.833 | -0.025 | 0.121 |
| RLIC | OTP | AD | -0.448 | 0.654 | -0.054 | 0.121 |
| SCC | OTP | AD | 0.548 | 0.584 | 0.066 | 0.121 |
| SCR | OTP | AD | 0.106 | 0.915 | 0.013 | 0.121 |
| SFO | OTP | AD | -1.367 | 0.172 | -0.164 | 0.121 |
| SLF | OTP | AD | -0.676 | 0.499 | -0.081 | 0.121 |
| SS | OTP | AD | 0.090 | 0.929 | 0.011 | 0.121 |
| UNC | OTP | AD | 1.207 | 0.228 | 0.145 | 0.121 |

Significant results are highlighted in bold ($p < 0.002$). Abbreviation: FA = fractional anisotropy, MD = mean diffusivity, RD = radial diffusivity, AD = axial diffusivity, S.E. = standard error, EOS = early-onset schizophrenia, AFP = affective psychosis, OTP = other psychosis. Abbreviations for tracts, see Table 1 in main text.

**Table S11| Multiple linear regression output for sex-by-diagnostic group interactions in bilateral regional diffusion measures.**

| Tract | metric | t-value | p-value | Cohen's *d* | S.E. |
|---|---|---|---|---|---|
| ACR | FA | 1.605 | 0.109 | 0.134 | 0.083 |
| | MD | -1.704 | 0.089 | -0.153 | 0.089 |
| | RD | -1.878 | 0.061 | -0.169 | 0.089 |
| | AD | -0.888 | 0.375 | -0.080 | 0.089 |
| ALIC | FA | 0.644 | 0.520 | 0.054 | 0.083 |
| | MD | -0.548 | 0.584 | -0.049 | 0.089 |
| | RD | -0.438 | 0.662 | -0.039 | 0.089 |
| | AD | -0.688 | 0.492 | -0.062 | 0.089 |
| Average | FA | 2.792 | 0.005 | 0.233 | 0.083 |
| | MD | -1.458 | 0.145 | -0.131 | 0.089 |
| | RD | -2.126 | 0.034 | -0.191 | 0.089 |
| | AD | 0.262 | 0.793 | 0.024 | 0.089 |
| BCC | FA | 1.781 | 0.076 | 0.149 | 0.083 |
| | MD | -0.684 | 0.494 | -0.061 | 0.089 |
| | RD | -1.953 | 0.051 | -0.175 | 0.089 |
| | AD | 1.638 | 0.102 | 0.147 | 0.089 |
| CC | FA | 2.266 | 0.024 | 0.189 | 0.083 |
| | MD | -1.433 | 0.153 | -0.129 | 0.089 |
| | RD | -2.549 | 0.011 | -0.229 | 0.089 |
| | AD | 0.781 | 0.435 | 0.070 | 0.089 |
| CGC | FA | 1.952 | 0.051 | 0.163 | 0.083 |
| | MD | -0.731 | 0.465 | -0.066 | 0.089 |
| | RD | -2.119 | 0.035 | -0.190 | 0.089 |
| | AD | 1.079 | 0.281 | 0.097 | 0.089 |
| CGH | FA | 1.791 | 0.074 | 0.150 | 0.083 |
| | MD | -1.549 | 0.122 | -0.139 | 0.089 |
| | RD | -1.798 | 0.073 | -0.161 | 0.089 |
| | AD | -0.328 | 0.743 | -0.029 | 0.089 |
| CR | FA | 1.636 | 0.102 | 0.137 | 0.083 |
| | MD | -2.082 | 0.038 | -0.187 | 0.089 |
| | RD | -2.198 | 0.028 | -0.197 | 0.089 |
| | AD | -1.297 | 0.195 | -0.116 | 0.089 |
| CST | FA | 1.150 | 0.251 | 0.096 | 0.083 |
| | MD | -1.386 | 0.166 | -0.124 | 0.089 |
| | RD | -1.498 | 0.135 | -0.134 | 0.089 |
| | AD | -1.135 | 0.257 | -0.102 | 0.089 |
| EC | FA | 2.225 | 0.026 | 0.186 | 0.083 |
| | MD | -0.849 | 0.396 | -0.076 | 0.089 |
| | RD | -1.645 | 0.101 | -0.148 | 0.089 |
| | AD | 0.538 | 0.591 | 0.048 | 0.089 |
| FX | FA | 2.575 | 0.010 | 0.215 | 0.083 |
| | MD | -0.795 | 0.427 | -0.071 | 0.089 |
| | RD | -1.072 | 0.284 | -0.096 | 0.089 |

| | | | | | |
|---|---|---|---|---|---|
| | AD | -0.339 | 0.735 | -0.030 | 0.089 |
| FXST | FA | 2.130 | 0.034 | 0.178 | 0.083 |
| | MD | -2.105 | 0.036 | -0.189 | 0.089 |
| | RD | -2.904 | 0.004 | -0.261 | 0.089 |
| | AD | -0.224 | 0.823 | -0.020 | 0.089 |
| GCC | FA | 1.757 | 0.079 | 0.147 | 0.083 |
| | MD | -1.329 | 0.184 | -0.119 | 0.089 |
| | RD | -1.943 | 0.053 | -0.174 | 0.089 |
| | AD | -0.256 | 0.798 | -0.023 | 0.089 |
| IC | FA | 0.755 | 0.450 | 0.063 | 0.083 |
| | MD | -1.581 | 0.115 | -0.142 | 0.089 |
| | RD | -1.362 | 0.174 | -0.122 | 0.089 |
| | AD | -1.371 | 0.171 | -0.123 | 0.089 |
| IFO | FA | 1.622 | 0.105 | 0.136 | 0.083 |
| | MD | -0.804 | 0.422 | -0.072 | 0.089 |
| | RD | -1.391 | 0.165 | -0.125 | 0.089 |
| | AD | 0.663 | 0.508 | 0.059 | 0.089 |
| PCR | FA | 0.879 | 0.380 | 0.073 | 0.083 |
| | MD | -2.292 | 0.022 | -0.206 | 0.089 |
| | RD | -1.880 | 0.061 | -0.169 | 0.089 |
| | AD | -1.784 | 0.075 | -0.160 | 0.089 |
| PLIC | FA | 0.202 | 0.840 | 0.017 | 0.083 |
| | MD | -1.636 | 0.102 | -0.147 | 0.089 |
| | RD | -0.865 | 0.388 | -0.078 | 0.089 |
| | AD | -1.518 | 0.130 | -0.136 | 0.089 |
| PTR | FA | 0.774 | 0.439 | 0.065 | 0.083 |
| | MD | -2.089 | 0.037 | -0.187 | 0.089 |
| | RD | -1.664 | 0.097 | -0.149 | 0.089 |
| | AD | -1.597 | 0.111 | -0.143 | 0.089 |
| RLIC | FA | 1.106 | 0.269 | 0.092 | 0.083 |
| | MD | -1.921 | 0.055 | -0.172 | 0.089 |
| | RD | -2.197 | 0.028 | -0.197 | 0.089 |
| | AD | -0.702 | 0.483 | -0.063 | 0.089 |
| SCC | FA | 2.379 | 0.018 | 0.199 | 0.083 |
| | MD | -2.148 | 0.032 | -0.193 | 0.089 |
| | RD | -3.202 | **0.001** | -0.287 | 0.089 |
| | AD | 0.225 | 0.822 | 0.020 | 0.089 |
| SCR | FA | 1.291 | 0.197 | 0.108 | 0.083 |
| | MD | -2.034 | 0.042 | -0.183 | 0.089 |
| | RD | -2.141 | 0.033 | -0.192 | 0.089 |
| | AD | -1.027 | 0.305 | -0.092 | 0.089 |
| SFO | FA | 1.338 | 0.182 | 0.112 | 0.083 |
| | MD | -1.330 | 0.184 | -0.119 | 0.089 |
| | RD | -1.231 | 0.219 | -0.111 | 0.089 |
| | AD | -0.751 | 0.453 | -0.067 | 0.089 |
| SLF | FA | 1.960 | 0.050 | 0.164 | 0.083 |

| | | | | | |
|---|---|---|---|---|---|
| | MD | -1.898 | 0.058 | -0.170 | 0.089 |
| | RD | -2.114 | 0.035 | -0.190 | 0.089 |
| | AD | -0.682 | 0.496 | -0.061 | 0.089 |
| SS | FA | 1.493 | 0.136 | 0.125 | 0.083 |
| | MD | -1.891 | 0.059 | -0.170 | 0.089 |
| | RD | -2.184 | 0.029 | -0.196 | 0.089 |
| | AD | -0.463 | 0.643 | -0.042 | 0.089 |
| UNC | FA | 1.496 | 0.135 | 0.125 | 0.083 |
| | MD | -0.988 | 0.323 | -0.089 | 0.089 |
| | RD | -1.620 | 0.106 | -0.145 | 0.089 |
| | AD | 0.482 | 0.630 | 0.043 | 0.089 |

Significant results are highlighted in bold ($p < 0.002$). Abbreviation: FA = fractional anisotropy, MD = mean diffusivity, RD = radial diffusivity, AD = axial diffusivity, S.E. = standard error. Abbreviations for tracts, see Table 1 in main text.

**Table S12| Multiple linear regression output for age-by-diagnostic group interactions in bilateral regional diffusion measures.**

| Tract | metric | t-value | p-value | Cohen's *d* | S.E. |
|---|---|---|---|---|---|
| ACR | FA | 1.293 | 0.197 | 0.108 | 0.083 |
| | MD | -0.327 | 0.744 | -0.029 | 0.089 |
| | RD | -0.841 | 0.401 | -0.075 | 0.089 |
| | AD | -0.888 | 0.375 | -0.080 | 0.089 |
| ALIC | FA | 0.144 | 0.886 | 0.012 | 0.083 |
| | MD | 0.641 | 0.522 | 0.058 | 0.089 |
| | RD | -0.176 | 0.860 | -0.016 | 0.089 |
| | AD | -0.688 | 0.492 | -0.062 | 0.089 |
| Average | FA | 0.981 | 0.327 | 0.082 | 0.083 |
| | MD | 0.028 | 0.978 | 0.002 | 0.089 |
| | RD | -0.226 | 0.821 | -0.020 | 0.089 |
| | AD | 0.262 | 0.793 | 0.024 | 0.089 |
| BCC | FA | -0.573 | 0.567 | -0.048 | 0.083 |
| | MD | 0.936 | 0.350 | 0.084 | 0.089 |
| | RD | 0.637 | 0.524 | 0.057 | 0.089 |
| | AD | 1.638 | 0.102 | 0.147 | 0.089 |
| CC | FA | 0.149 | 0.881 | 0.012 | 0.083 |
| | MD | 0.643 | 0.520 | 0.058 | 0.089 |
| | RD | 0.123 | 0.902 | 0.011 | 0.089 |
| | AD | 0.781 | 0.435 | 0.070 | 0.089 |
| CGC | FA | -0.298 | 0.766 | -0.025 | 0.083 |
| | MD | -0.185 | 0.853 | -0.017 | 0.089 |
| | RD | 0.418 | 0.676 | 0.038 | 0.089 |
| | AD | 1.079 | 0.281 | 0.097 | 0.089 |
| CGH | FA | 0.207 | 0.836 | 0.017 | 0.083 |
| | MD | 0.815 | 0.416 | 0.073 | 0.089 |
| | RD | 0.262 | 0.794 | 0.023 | 0.089 |
| | AD | -0.328 | 0.743 | -0.029 | 0.089 |
| CR | FA | 0.776 | 0.438 | 0.065 | 0.083 |
| | MD | -0.226 | 0.821 | -0.020 | 0.089 |
| | RD | -0.698 | 0.486 | -0.063 | 0.089 |
| | AD | -1.297 | 0.195 | -0.116 | 0.089 |
| CST | FA | 0.131 | 0.896 | 0.011 | 0.083 |
| | MD | 1.570 | 0.117 | 0.141 | 0.089 |
| | RD | 1.164 | 0.245 | 0.104 | 0.089 |
| | AD | -1.135 | 0.257 | -0.102 | 0.089 |
| EC | FA | -0.096 | 0.923 | -0.008 | 0.083 |
| | MD | 0.405 | 0.686 | 0.036 | 0.089 |
| | RD | 0.145 | 0.884 | 0.013 | 0.089 |
| | AD | 0.538 | 0.591 | 0.048 | 0.089 |
| FX | FA | 0.388 | 0.698 | 0.032 | 0.083 |
| | MD | 1.701 | 0.090 | 0.153 | 0.089 |
| | RD | 1.730 | 0.084 | 0.155 | 0.089 |

| | | | | | |
|---|---|---|---|---|---|
| | AD | -0.339 | 0.735 | -0.030 | 0.089 |
| FXST | FA | -1.252 | 0.211 | -0.105 | 0.083 |
| | MD | 0.561 | 0.575 | 0.050 | 0.089 |
| | RD | 0.697 | 0.486 | 0.063 | 0.089 |
| | AD | -0.224 | 0.823 | -0.020 | 0.089 |
| GCC | FA | 0.839 | 0.402 | 0.070 | 0.083 |
| | MD | 0.373 | 0.709 | 0.034 | 0.089 |
| | RD | -0.526 | 0.599 | -0.047 | 0.089 |
| | AD | -0.256 | 0.798 | -0.023 | 0.089 |
| IC | FA | 0.853 | 0.394 | 0.071 | 0.083 |
| | MD | 0.083 | 0.934 | 0.007 | 0.089 |
| | RD | -0.837 | 0.403 | -0.075 | 0.089 |
| | AD | -1.371 | 0.171 | -0.123 | 0.089 |
| IFO | FA | 0.939 | 0.348 | 0.079 | 0.083 |
| | MD | 0.331 | 0.741 | 0.030 | 0.089 |
| | RD | -0.550 | 0.582 | -0.049 | 0.089 |
| | AD | 0.663 | 0.508 | 0.059 | 0.089 |
| PCR | FA | 0.356 | 0.722 | 0.030 | 0.083 |
| | MD | -0.401 | 0.688 | -0.036 | 0.089 |
| | RD | -0.795 | 0.427 | -0.071 | 0.089 |
| | AD | -1.784 | 0.075 | -0.160 | 0.089 |
| PLIC | FA | 1.664 | 0.097 | 0.139 | 0.083 |
| | MD | -0.359 | 0.720 | -0.032 | 0.089 |
| | RD | -1.482 | 0.139 | -0.133 | 0.089 |
| | AD | -1.518 | 0.130 | -0.136 | 0.089 |
| PTR | FA | 0.128 | 0.899 | 0.011 | 0.083 |
| | MD | -1.115 | 0.265 | -0.100 | 0.089 |
| | RD | -0.801 | 0.423 | -0.072 | 0.089 |
| | AD | -1.597 | 0.111 | -0.143 | 0.089 |
| RLIC | FA | 0.177 | 0.860 | 0.015 | 0.083 |
| | MD | 0.063 | 0.950 | 0.006 | 0.089 |
| | RD | -0.268 | 0.789 | -0.024 | 0.089 |
| | AD | -0.702 | 0.483 | -0.063 | 0.089 |
| SCC | FA | 0.689 | 0.491 | 0.058 | 0.083 |
| | MD | 0.367 | 0.714 | 0.033 | 0.089 |
| | RD | -0.304 | 0.761 | -0.027 | 0.089 |
| | AD | 0.225 | 0.822 | 0.020 | 0.089 |
| SCR | FA | -0.114 | 0.909 | -0.010 | 0.083 |
| | MD | 0.156 | 0.876 | 0.014 | 0.089 |
| | RD | -0.056 | 0.955 | -0.005 | 0.089 |
| | AD | -1.027 | 0.305 | -0.092 | 0.089 |
| SFO | FA | -0.133 | 0.894 | -0.011 | 0.083 |
| | MD | 0.134 | 0.893 | 0.012 | 0.089 |
| | RD | 0.067 | 0.946 | 0.006 | 0.089 |
| | AD | -0.751 | 0.453 | -0.067 | 0.089 |
| SLF | FA | 1.081 | 0.280 | 0.090 | 0.083 |

| | | | | | |
|---|---|---|---|---|---|
| | MD | -0.816 | 0.415 | -0.073 | 0.089 |
| | RD | -1.177 | 0.240 | -0.106 | 0.089 |
| | AD | -0.682 | 0.496 | -0.061 | 0.089 |
| SS | FA | -0.106 | 0.915 | -0.009 | 0.083 |
| | MD | -0.037 | 0.970 | -0.003 | 0.089 |
| | RD | -0.220 | 0.826 | -0.020 | 0.089 |
| | AD | -0.463 | 0.643 | -0.042 | 0.089 |
| UNC | FA | -0.344 | 0.731 | -0.029 | 0.083 |
| | MD | 0.208 | 0.835 | 0.019 | 0.089 |
| | RD | 0.048 | 0.962 | 0.004 | 0.089 |
| | AD | 0.482 | 0.630 | 0.043 | 0.089 |

Abbreviation: FA = fractional anisotropy, MD = mean diffusivity, RD = radial diffusivity, AD = axial diffusivity, S.E. = standard error. Abbreviations for tracts, see Table 1 in main text.

**Table S13| Multiple linear regression output for association between medication use and bilateral regional diffusion measures, in patients with early-onset psychosis.**

| | | CPZ | | AP | | Lithium | | AD | | AE | |
|---|---|---|---|---|---|---|---|---|---|---|---|
| Tract | metric | t-value | p-value | t-value | p-value | t-value | p-value | t-value | p-value | t-value | p-value |
| AverageFA | FA | 1.108 | 0.269 | -0.190 | 0.850 | -0.325 | 0.746 | -0.400 | 0.690 | 0.474 | 0.636 |
| ACR | FA | 0.576 | 0.565 | -0.004 | 0.997 | 0.964 | 0.336 | 0.359 | 0.720 | -0.204 | 0.839 |
| ALIC | FA | 1.592 | 0.113 | 0.521 | 0.603 | 1.293 | 0.197 | -0.384 | 0.701 | 0.229 | 0.819 |
| BCC | FA | -0.523 | 0.602 | 0.701 | 0.484 | 0.843 | 0.400 | 0.040 | 0.968 | -0.168 | 0.867 |
| CC | FA | -0.793 | 0.429 | 0.417 | 0.677 | 1.255 | 0.211 | 0.295 | 0.768 | -0.920 | 0.359 |
| CGC | FA | 0.889 | 0.375 | 0.602 | 0.547 | 1.210 | 0.227 | 1.125 | 0.262 | -0.540 | 0.590 |
| CGH | FA | 1.088 | 0.278 | -0.383 | 0.702 | 0.905 | 0.366 | 0.773 | 0.440 | -0.611 | 0.542 |
| CR | FA | 0.626 | 0.532 | -0.478 | 0.633 | 0.923 | 0.357 | 0.372 | 0.710 | 0.739 | 0.461 |
| CST | FA | 0.071 | 0.944 | -0.878 | 0.381 | 0.143 | 0.886 | 0.838 | 0.403 | -0.956 | 0.340 |
| EC | FA | 0.678 | 0.499 | -0.202 | 0.840 | 0.278 | 0.781 | 1.013 | 0.312 | -0.938 | 0.349 |
| FX | FA | 0.323 | 0.747 | 0.211 | 0.833 | 1.379 | 0.169 | -0.073 | 0.942 | -0.518 | 0.605 |
| FXST | FA | 1.157 | 0.249 | -0.779 | 0.437 | -0.244 | 0.807 | 0.604 | 0.547 | 0.906 | 0.366 |
| GCC | FA | -0.500 | 0.618 | -0.144 | 0.886 | 0.385 | 0.700 | 0.828 | 0.409 | -0.651 | 0.516 |
| IC | FA | 0.883 | 0.378 | -0.210 | 0.834 | 1.937 | 0.054 | 0.180 | 0.857 | 0.423 | 0.673 |
| IFO | FA | 0.007 | 0.994 | 0.828 | 0.408 | 0.054 | 0.957 | 1.587 | 0.114 | -0.568 | 0.570 |
| PCR | FA | -0.059 | 0.953 | -0.449 | 0.654 | 1.700 | 0.090 | -0.468 | 0.640 | 0.715 | 0.475 |
| PLIC | FA | -0.199 | 0.842 | -0.592 | 0.554 | 1.714 | 0.088 | 0.780 | 0.436 | 1.147 | 0.252 |
| PTR | FA | 0.946 | 0.345 | -1.693 | 0.092 | 0.324 | 0.746 | -0.118 | 0.906 | 1.457 | 0.146 |
| RLIC | FA | 0.838 | 0.403 | -0.387 | 0.699 | 1.594 | 0.112 | 0.166 | 0.868 | -0.225 | 0.822 |
| SCC | FA | -1.202 | 0.231 | 0.263 | 0.793 | 1.819 | 0.070 | 0.156 | 0.877 | -1.997 | 0.047 |
| SCR | FA | 0.924 | 0.356 | -0.976 | 0.330 | -0.042 | 0.967 | 0.892 | 0.373 | 1.814 | 0.071 |
| SFO | FA | 0.940 | 0.348 | 1.881 | 0.061 | 0.334 | 0.739 | 0.444 | 0.657 | 2.744 | 0.006 |
| SLF | FA | 0.410 | 0.682 | -0.289 | 0.773 | 0.825 | 0.410 | 0.980 | 0.328 | 0.027 | 0.978 |
| SS | FA | 0.857 | 0.392 | -1.352 | 0.177 | -0.121 | 0.904 | -0.040 | 0.968 | -0.152 | 0.879 |
| UNC | FA | 1.003 | 0.317 | 0.310 | 0.757 | 0.050 | 0.960 | 0.119 | 0.906 | 0.482 | 0.630 |
| AverageMD | MD | -0.071 | 0.944 | 1.931 | 0.055 | 0.231 | 0.817 | 1.396 | 0.164 | 0.408 | 0.684 |
| ACR | MD | -0.537 | 0.592 | 0.897 | 0.370 | 0.464 | 0.643 | -0.272 | 0.786 | 0.452 | 0.651 |
| ALIC | MD | -1.247 | 0.214 | 0.367 | 0.714 | -0.091 | 0.928 | -1.255 | 0.211 | -0.579 | 0.563 |
| BCC | MD | 0.136 | 0.892 | 0.269 | 0.788 | 0.181 | 0.856 | -0.637 | 0.525 | -0.386 | 0.700 |

| | | | | | | | | | | | |
|---|---|---|---|---|---|---|---|---|---|---|---|
| CC | MD | 0.142 | 0.887 | 0.225 | 0.822 | -0.025 | 0.980 | -0.885 | 0.377 | 0.029 | 0.977 |
| CGC | MD | 0.441 | 0.660 | 1.164 | 0.246 | -0.475 | 0.635 | -1.206 | 0.229 | -1.319 | 0.189 |
| CGH | MD | -0.811 | 0.418 | -0.679 | 0.498 | -1.036 | 0.301 | -1.131 | 0.259 | -0.551 | 0.582 |
| CR | MD | -0.536 | 0.593 | 1.264 | 0.208 | -0.086 | 0.932 | -0.547 | 0.585 | -0.212 | 0.832 |
| CST | MD | -0.651 | 0.516 | 1.209 | 0.228 | -0.052 | 0.959 | -0.917 | 0.360 | 0.514 | 0.608 |
| EC | MD | -1.083 | 0.280 | 0.492 | 0.623 | -0.273 | 0.785 | -0.989 | 0.324 | 0.828 | 0.409 |
| FX | MD | 0.528 | 0.598 | 0.512 | 0.609 | -1.495 | 0.136 | 0.078 | 0.938 | 1.362 | 0.175 |
| FXST | MD | -0.740 | 0.460 | 0.361 | 0.719 | -1.137 | 0.257 | 1.071 | 0.285 | -0.168 | 0.866 |
| GCC | MD | 0.033 | 0.974 | 0.070 | 0.944 | 0.836 | 0.404 | -0.744 | 0.458 | 0.867 | 0.387 |
| IC | MD | -0.975 | 0.331 | 0.783 | 0.434 | -1.061 | 0.290 | -0.946 | 0.345 | -0.518 | 0.605 |
| IFO | MD | -0.989 | 0.324 | 0.196 | 0.844 | -0.180 | 0.857 | -1.729 | 0.085 | -0.072 | 0.943 |
| PCR | MD | -0.437 | 0.662 | 0.924 | 0.357 | -0.575 | 0.566 | 0.007 | 0.995 | -1.433 | 0.153 |
| PLIC | MD | -0.826 | 0.410 | 1.488 | 0.138 | -1.453 | 0.148 | -1.286 | 0.200 | -0.220 | 0.826 |
| PTR | MD | -0.811 | 0.418 | 1.416 | 0.158 | 0.255 | 0.799 | 0.107 | 0.915 | -1.786 | 0.076 |
| RLIC | MD | -0.370 | 0.712 | 0.279 | 0.781 | -1.171 | 0.243 | -0.033 | 0.974 | -0.410 | 0.682 |
| SCC | MD | 0.239 | 0.811 | 0.210 | 0.834 | -1.129 | 0.260 | -1.074 | 0.284 | 0.035 | 0.972 |
| SCR | MD | -0.394 | 0.694 | 1.715 | 0.088 | -0.500 | 0.617 | -1.071 | 0.285 | -0.264 | 0.792 |
| SFO | MD | -0.093 | 0.926 | 1.391 | 0.165 | 0.834 | 0.405 | -0.999 | 0.319 | -1.654 | 0.099 |
| SLF | MD | -0.123 | 0.902 | 0.945 | 0.345 | -0.892 | 0.373 | -0.980 | 0.328 | -0.774 | 0.440 |
| SS | MD | -0.965 | 0.336 | 1.349 | 0.178 | -0.739 | 0.460 | -0.851 | 0.396 | -0.426 | 0.670 |
| UNC | MD | 0.297 | 0.767 | 0.087 | 0.931 | -0.269 | 0.788 | 0.014 | 0.989 | -1.128 | 0.261 |
| AverageRD | RD | -0.144 | 0.886 | 1.671 | 0.096 | 0.457 | 0.648 | 1.526 | 0.128 | 0.201 | 0.841 |
| ACR | RD | -0.618 | 0.537 | 1.079 | 0.282 | 0.153 | 0.879 | -0.419 | 0.676 | 0.473 | 0.636 |
| ALIC | RD | -1.393 | 0.165 | -0.187 | 0.852 | -0.775 | 0.439 | -0.349 | 0.728 | -0.398 | 0.691 |
| BCC | RD | 0.519 | 0.605 | -0.379 | 0.705 | -0.238 | 0.812 | -0.150 | 0.881 | -0.018 | 0.985 |
| CC | RD | 0.644 | 0.521 | -0.333 | 0.740 | -0.551 | 0.582 | -0.449 | 0.654 | 0.309 | 0.758 |
| CGC | RD | -0.761 | 0.447 | 0.335 | 0.738 | -1.212 | 0.227 | -1.423 | 0.156 | -0.393 | 0.694 |
| CGH | RD | -0.673 | 0.501 | -0.087 | 0.931 | -1.070 | 0.286 | -0.822 | 0.412 | -0.569 | 0.570 |
| CR | RD | -0.758 | 0.450 | 1.397 | 0.164 | -0.364 | 0.717 | -0.609 | 0.543 | -0.608 | 0.544 |
| CST | RD | -0.441 | 0.660 | 1.466 | 0.144 | 0.005 | 0.996 | -0.892 | 0.373 | 1.010 | 0.313 |
| EC | RD | -0.976 | 0.331 | 0.775 | 0.439 | -0.367 | 0.714 | -1.160 | 0.247 | 1.153 | 0.250 |
| FX | RD | 0.521 | 0.603 | 0.625 | 0.533 | -1.581 | 0.115 | 0.153 | 0.879 | 1.400 | 0.163 |
| FXST | RD | -0.726 | 0.469 | 0.574 | 0.567 | -0.889 | 0.375 | 0.184 | 0.854 | -0.610 | 0.543 |
| GCC | RD | 0.128 | 0.898 | 0.016 | 0.987 | 0.258 | 0.797 | -0.644 | 0.521 | 0.067 | 0.947 |

| | | | | | | | | | | | |
|---|---|---|---|---|---|---|---|---|---|---|---|
| IC | RD | -0.958 | 0.339 | 0.687 | 0.493 | -1.514 | 0.131 | -0.524 | 0.601 | -0.742 | 0.459 |
| IFO | RD | -0.963 | 0.337 | -0.194 | 0.846 | 0.195 | 0.845 | -1.789 | 0.075 | 0.229 | 0.819 |
| PCR | RD | -0.431 | 0.667 | 1.026 | 0.306 | -1.098 | 0.274 | 0.273 | 0.785 | -1.625 | 0.106 |
| PLIC | RD | -0.174 | 0.862 | 1.145 | 0.253 | -1.552 | 0.122 | -0.916 | 0.361 | -1.221 | 0.223 |
| PTR | RD | -0.763 | 0.446 | 1.582 | 0.115 | 0.173 | 0.863 | 0.011 | 0.991 | -1.828 | 0.069 |
| RLIC | RD | -0.880 | 0.380 | 0.645 | 0.519 | -1.499 | 0.135 | -0.165 | 0.869 | -0.357 | 0.721 |
| SCC | RD | 0.986 | 0.326 | -0.278 | 0.782 | -1.574 | 0.117 | -0.600 | 0.549 | 1.038 | 0.300 |
| SCR | RD | -0.868 | 0.387 | 1.744 | 0.082 | -0.305 | 0.761 | -1.284 | 0.200 | -1.326 | 0.186 |
| SFO | RD | -1.058 | 0.291 | -0.404 | 0.687 | 0.245 | 0.807 | -0.948 | 0.344 | -2.420 | 0.016 |
| SLF | RD | -0.557 | 0.578 | 0.896 | 0.371 | -0.863 | 0.389 | -1.159 | 0.248 | -0.964 | 0.336 |
| SS | RD | -0.967 | 0.335 | 1.835 | 0.068 | -0.044 | 0.965 | -0.355 | 0.723 | -0.585 | 0.559 |
| UNC | RD | -0.476 | 0.635 | -0.069 | 0.945 | 0.028 | 0.978 | 0.215 | 0.830 | -0.783 | 0.434 |
| AverageAD | AD | 0.297 | 0.767 | 1.600 | 0.111 | -0.361 | 0.719 | 0.786 | 0.432 | 0.663 | 0.508 |
| ACR | AD | 0.132 | 0.895 | 0.600 | 0.549 | 0.955 | 0.341 | -0.044 | 0.965 | 0.323 | 0.747 |
| ALIC | AD | -0.963 | 0.337 | 0.643 | 0.521 | 0.933 | 0.352 | -2.056 | 0.041 | -0.624 | 0.533 |
| BCC | AD | -0.057 | 0.955 | 0.972 | 0.332 | 0.979 | 0.329 | -1.348 | 0.179 | -0.216 | 0.829 |
| CC | AD | -0.299 | 0.765 | 0.648 | 0.518 | 0.771 | 0.441 | -1.178 | 0.240 | 0.046 | 0.963 |
| CGC | AD | 1.333 | 0.184 | 0.832 | 0.406 | 0.701 | 0.484 | -0.581 | 0.562 | -0.996 | 0.320 |
| CGH | AD | -0.225 | 0.822 | -0.833 | 0.406 | -0.312 | 0.755 | -1.285 | 0.200 | -0.166 | 0.868 |
| CR | AD | 0.203 | 0.840 | 0.558 | 0.578 | 0.472 | 0.637 | -0.300 | 0.765 | 0.590 | 0.556 |
| CST | AD | -0.771 | 0.441 | 0.496 | 0.620 | -0.006 | 0.996 | -1.167 | 0.244 | -0.293 | 0.770 |
| EC | AD | -0.927 | 0.355 | -0.070 | 0.944 | 0.257 | 0.798 | -0.339 | 0.735 | 0.253 | 0.800 |
| FX | AD | 0.514 | 0.608 | 0.525 | 0.600 | -1.121 | 0.263 | 0.079 | 0.937 | 1.253 | 0.211 |
| FXST | AD | -0.211 | 0.833 | -0.132 | 0.895 | -1.127 | 0.261 | 1.560 | 0.120 | 0.935 | 0.351 |
| GCC | AD | -0.132 | 0.895 | 0.067 | 0.946 | 1.296 | 0.196 | -0.582 | 0.561 | 1.356 | 0.176 |
| IC | AD | -0.612 | 0.541 | 0.431 | 0.667 | 0.065 | 0.948 | -1.355 | 0.177 | 0.183 | 0.855 |
| IFO | AD | -0.331 | 0.741 | 0.774 | 0.440 | -0.212 | 0.832 | -0.719 | 0.473 | -0.242 | 0.809 |
| PCR | AD | -0.329 | 0.742 | 0.137 | 0.891 | 0.660 | 0.510 | -0.470 | 0.639 | -0.449 | 0.654 |
| PLIC | AD | -1.314 | 0.191 | 1.098 | 0.273 | -0.245 | 0.807 | -1.267 | 0.206 | 0.963 | 0.337 |
| PTR | AD | -0.326 | 0.745 | 0.283 | 0.777 | 0.322 | 0.748 | 0.314 | 0.754 | -0.578 | 0.564 |
| RLIC | AD | 0.892 | 0.374 | -0.542 | 0.589 | -0.490 | 0.625 | 0.037 | 0.971 | 0.121 | 0.904 |
| SCC | AD | -0.600 | 0.549 | 0.400 | 0.689 | -0.292 | 0.771 | -1.021 | 0.308 | -0.656 | 0.512 |
| SCR | AD | 0.531 | 0.596 | 0.566 | 0.572 | -0.474 | 0.636 | -0.473 | 0.637 | 1.297 | 0.196 |
| SFO | AD | 0.526 | 0.600 | 2.258 | 0.025 | 1.148 | 0.252 | -0.636 | 0.526 | 0.028 | 0.978 |

| | | | | | | | | | | | |
|---|---|---|---|---|---|---|---|---|---|---|---|
| SLF | AD | 0.750 | 0.454 | 0.431 | 0.667 | -0.544 | 0.587 | -0.608 | 0.544 | 0.103 | 0.918 |
| SS | AD | -0.560 | 0.576 | -0.391 | 0.696 | -1.131 | 0.259 | -1.393 | 0.165 | 0.163 | 0.871 |
| UNC | AD | 1.134 | 0.258 | 0.286 | 0.775 | -0.222 | 0.825 | -0.383 | 0.702 | -1.239 | 0.217 |

Abbreviation: FA = fractional anisotropy, MD = mean diffusivity, RD = radial diffusivity, AD = axial diffusivity, CPZ = chlorpromazine equivalent, AP = antipsychotics, AD = antidepressants, AE = antiepileptics. Abbreviations for tracts, see Table 1 in main text.

**Table S14| Multiple linear regression output for association between clinical measures and bilateral regional diffusion measures, in patients with early-onset psychosis.**

| | | AOO | | DOI | | PANSS, negative | | PANSS, positive | |
|---|---|---|---|---|---|---|---|---|---|
| **Tract** | **metric** | **t-value** | **p-value** | **t-value** | **p-value** | **t-value** | **p-value** | **t-value** | **p-value** |
| AverageFA | FA | 1.532 | 0.127 | 1.096 | 0.274 | 1.224 | 0.222 | -0.440 | 0.660 |
| ACR | FA | 2.160 | 0.032 | -0.166 | 0.868 | 0.111 | 0.912 | 0.901 | 0.368 |
| ALIC | FA | 2.761 | 0.006 | 0.658 | 0.511 | 0.527 | 0.599 | 0.826 | 0.409 |
| BCC | FA | 1.058 | 0.291 | -0.241 | 0.810 | 1.458 | 0.146 | 0.054 | 0.957 |
| CC | FA | 1.887 | 0.060 | -1.047 | 0.296 | 1.372 | 0.171 | 0.188 | 0.851 |
| CGC | FA | 2.484 | 0.014 | 0.516 | 0.606 | 1.517 | 0.131 | -0.196 | 0.845 |
| CGH | FA | 1.582 | 0.115 | 0.323 | 0.747 | 0.646 | 0.519 | -1.201 | 0.231 |
| CR | FA | 1.897 | 0.059 | -0.181 | 0.857 | -0.014 | 0.989 | 0.027 | 0.979 |
| CST | FA | 1.901 | 0.058 | -0.043 | 0.966 | 0.727 | 0.468 | 0.096 | 0.924 |
| EC | FA | 1.800 | 0.073 | 0.446 | 0.656 | 2.314 | 0.022 | 1.646 | 0.101 |
| FX | FA | 0.547 | 0.585 | 1.325 | 0.186 | -0.957 | 0.339 | 0.065 | 0.949 |
| FXST | FA | 1.544 | 0.124 | 0.369 | 0.712 | 1.431 | 0.154 | -0.933 | 0.352 |
| GCC | FA | 1.751 | 0.081 | -1.073 | 0.284 | 0.793 | 0.428 | 0.353 | 0.725 |
| IC | FA | 2.245 | 0.026 | 0.529 | 0.597 | 0.934 | 0.351 | 0.052 | 0.959 |
| IFO | FA | 0.650 | 0.516 | 1.035 | 0.301 | 0.869 | 0.386 | 0.617 | 0.538 |
| PCR | FA | 1.582 | 0.115 | -0.654 | 0.514 | -1.281 | 0.201 | -0.583 | 0.561 |
| PLIC | FA | 1.052 | 0.294 | 0.832 | 0.406 | 0.785 | 0.433 | 0.029 | 0.977 |
| PTR | FA | 1.567 | 0.118 | -0.793 | 0.429 | 1.143 | 0.254 | -1.585 | 0.114 |
| RLIC | FA | 1.604 | 0.110 | 0.062 | 0.950 | 1.006 | 0.316 | -0.675 | 0.501 |
| SCC | FA | 2.433 | 0.016 | -1.602 | 0.110 | 0.893 | 0.372 | 0.343 | 0.732 |
| SCR | FA | 0.788 | 0.431 | 0.377 | 0.706 | 0.684 | 0.495 | -0.785 | 0.433 |
| SFO | FA | 1.092 | 0.276 | 1.325 | 0.186 | 1.581 | 0.115 | 0.018 | 0.986 |
| SLF | FA | 2.778 | 0.006 | -0.370 | 0.712 | 1.321 | 0.188 | -0.166 | 0.868 |
| SS | FA | 2.057 | 0.041 | -0.239 | 0.811 | 2.228 | 0.027 | -0.623 | 0.534 |
| UNC | FA | 1.495 | 0.136 | -0.218 | 0.827 | -0.025 | 0.980 | -0.517 | 0.606 |
| AverageMD | MD | 1.515 | 0.131 | -2.961 | 0.003 | 0.196 | 0.845 | 0.487 | 0.627 |
| ACR | MD | 0.185 | 0.853 | -2.275 | 0.024 | -0.041 | 0.968 | 1.780 | 0.077 |
| ALIC | MD | 1.026 | 0.306 | -3.127 | **0.002** | 0.235 | 0.815 | 1.860 | 0.064 |
| BCC | MD | 1.305 | 0.193 | -2.323 | 0.021 | -1.594 | 0.113 | -0.477 | 0.634 |
| CC | MD | 1.197 | 0.232 | -2.628 | 0.009 | -1.077 | 0.283 | 0.165 | 0.869 |
| CGC | MD | 0.024 | 0.981 | -2.074 | 0.039 | -0.516 | 0.607 | 1.133 | 0.259 |
| CGH | MD | -0.374 | 0.709 | -0.427 | 0.669 | -0.185 | 0.853 | 1.164 | 0.246 |
| CR | MD | 0.250 | 0.803 | -2.537 | 0.012 | -0.040 | 0.968 | 2.165 | 0.032 |
| CST | MD | -0.668 | 0.505 | 0.119 | 0.905 | 0.389 | 0.698 | 0.497 | 0.620 |
| EC | MD | 0.633 | 0.527 | -2.341 | 0.020 | -0.360 | 0.719 | 0.814 | 0.417 |
| FX | MD | 1.355 | 0.177 | -0.790 | 0.431 | 0.720 | 0.472 | 0.101 | 0.919 |
| FXST | MD | -0.144 | 0.885 | -1.335 | 0.183 | -0.185 | 0.853 | 0.051 | 0.959 |
| GCC | MD | 0.963 | 0.336 | -1.942 | 0.053 | -0.583 | 0.560 | 1.339 | 0.182 |
| IC | MD | 0.797 | 0.426 | -3.116 | **0.002** | -0.060 | 0.952 | 1.790 | 0.075 |

| | | | | | | | | | |
|---|---|---|---|---|---|---|---|---|---|
| IFO | MD | -0.583 | 0.561 | -0.668 | 0.505 | -0.131 | 0.896 | 1.854 | 0.065 |
| PCR | MD | -0.168 | 0.867 | -2.015 | 0.045 | 0.876 | 0.382 | 1.983 | 0.049 |
| PLIC | MD | 0.777 | 0.438 | -2.911 | 0.004 | -0.248 | 0.805 | 2.013 | 0.046 |
| PTR | MD | -1.819 | 0.070 | -1.001 | 0.318 | 0.707 | 0.480 | 1.595 | 0.112 |
| RLIC | MD | 0.369 | 0.713 | -2.428 | 0.016 | 0.013 | 0.989 | 0.849 | 0.397 |
| SCC | MD | 0.482 | 0.630 | -2.021 | 0.044 | -0.115 | 0.909 | 0.114 | 0.910 |
| SCR | MD | 0.564 | 0.573 | -2.656 | 0.008 | -0.705 | 0.482 | 2.296 | 0.023 |
| SFO | MD | 0.458 | 0.647 | -2.440 | 0.015 | -0.075 | 0.940 | 1.589 | 0.114 |
| SLF | MD | -0.288 | 0.774 | -2.323 | 0.021 | -0.223 | 0.824 | 1.950 | 0.053 |
| SS | MD | -0.920 | 0.359 | -1.207 | 0.229 | 0.057 | 0.955 | 1.115 | 0.266 |
| UNC | MD | 0.215 | 0.830 | -1.376 | 0.170 | -0.103 | 0.918 | 0.720 | 0.472 |
| AverageRD | RD | 1.330 | 0.185 | -3.144 | **0.002** | 0.107 | 0.915 | 0.202 | 0.840 |
| ACR | RD | -0.485 | 0.628 | -1.912 | 0.057 | -0.254 | 0.800 | 0.630 | 0.530 |
| ALIC | RD | -1.330 | 0.185 | -2.136 | 0.034 | -0.319 | 0.750 | 0.406 | 0.685 |
| BCC | RD | 0.037 | 0.970 | -0.823 | 0.411 | -1.707 | 0.089 | -0.863 | 0.389 |
| CC | RD | -0.338 | 0.736 | -0.738 | 0.461 | -1.318 | 0.189 | -0.655 | 0.513 |
| CGC | RD | -1.566 | 0.119 | -1.115 | 0.266 | -1.302 | 0.194 | 0.484 | 0.629 |
| CGH | RD | -0.713 | 0.477 | -0.843 | 0.400 | -0.043 | 0.966 | 1.066 | 0.288 |
| CR | RD | -0.410 | 0.682 | -1.964 | 0.051 | 0.055 | 0.956 | 1.384 | 0.168 |
| CST | RD | -1.314 | 0.190 | -0.091 | 0.928 | 0.115 | 0.909 | 0.145 | 0.885 |
| EC | RD | -0.275 | 0.783 | -2.065 | 0.040 | -1.290 | 0.198 | -0.521 | 0.603 |
| FX | RD | 1.139 | 0.256 | -0.689 | 0.491 | 0.864 | 0.389 | -0.050 | 0.960 |
| FXST | RD | -0.904 | 0.367 | -1.353 | 0.177 | -0.735 | 0.463 | 0.201 | 0.841 |
| GCC | RD | -0.101 | 0.919 | -0.849 | 0.397 | -0.773 | 0.441 | 0.094 | 0.925 |
| IC | RD | -0.785 | 0.433 | -2.342 | 0.020 | -0.500 | 0.618 | 0.617 | 0.538 |
| IFO | RD | -0.178 | 0.859 | -1.593 | 0.112 | -0.990 | 0.323 | 0.497 | 0.619 |
| PCR | RD | -0.863 | 0.389 | -0.936 | 0.350 | 1.389 | 0.166 | 1.520 | 0.130 |
| PLIC | RD | -0.229 | 0.819 | -2.113 | 0.036 | -0.614 | 0.540 | 0.402 | 0.688 |
| PTR | RD | -1.380 | 0.169 | -0.561 | 0.575 | -0.170 | 0.865 | 1.225 | 0.222 |
| RLIC | RD | -0.318 | 0.751 | -1.952 | 0.052 | -0.333 | 0.739 | 0.764 | 0.446 |
| SCC | RD | -1.140 | 0.255 | -0.129 | 0.897 | -0.470 | 0.639 | -0.657 | 0.512 |
| SCR | RD | 0.270 | 0.787 | -2.148 | 0.033 | -0.590 | 0.556 | 2.024 | 0.044 |
| SFO | RD | -0.421 | 0.674 | -2.073 | 0.039 | -1.302 | 0.195 | 0.556 | 0.579 |
| SLF | RD | -1.328 | 0.185 | -1.646 | 0.101 | -0.617 | 0.538 | 1.323 | 0.187 |
| SS | RD | -1.140 | 0.255 | -1.393 | 0.165 | -0.901 | 0.369 | 0.917 | 0.360 |
| UNC | RD | -0.248 | 0.805 | -0.913 | 0.362 | 0.037 | 0.971 | 0.668 | 0.505 |
| AverageAD | AD | 1.160 | 0.247 | -1.364 | 0.174 | 0.504 | 0.615 | 0.677 | 0.499 |
| ACR | AD | 1.135 | 0.258 | -1.779 | 0.076 | -0.047 | 0.962 | 2.815 | 0.005 |
| ALIC | AD | 3.095 | **0.002** | -1.850 | 0.066 | 0.296 | 0.767 | 2.429 | 0.016 |
| BCC | AD | 2.660 | 0.008 | -3.491 | **0.001** | -0.726 | 0.468 | 0.320 | 0.749 |
| CC | AD | 2.622 | 0.009 | -3.383 | **0.001** | -0.427 | 0.670 | 1.056 | 0.292 |
| CGC | AD | 1.530 | 0.127 | -0.951 | 0.342 | 0.489 | 0.625 | 1.139 | 0.256 |
| CGH | AD | -0.004 | 0.997 | 0.909 | 0.364 | -0.621 | 0.535 | 0.621 | 0.535 |

| | | | | | | | | | |
|---|---|---|---|---|---|---|---|---|---|
| CR | AD | 1.043 | 0.298 | -2.143 | 0.033 | -0.535 | 0.593 | 2.574 | 0.011 |
| CST | AD | 0.005 | 0.996 | 0.465 | 0.643 | 0.712 | 0.477 | 0.867 | 0.387 |
| EC | AD | 1.459 | 0.146 | -1.160 | 0.247 | 0.881 | 0.379 | 2.534 | 0.012 |
| FX | AD | 1.639 | 0.102 | -0.921 | 0.358 | 0.529 | 0.597 | 0.397 | 0.692 |
| FXST | AD | 0.437 | 0.663 | -0.106 | 0.916 | 0.671 | 0.503 | -0.713 | 0.477 |
| GCC | AD | 1.882 | 0.061 | -2.058 | 0.041 | -0.221 | 0.825 | 1.849 | 0.066 |
| IC | AD | 2.114 | 0.035 | -1.975 | 0.049 | 0.119 | 0.906 | 2.434 | 0.016 |
| IFO | AD | -0.528 | 0.598 | 0.752 | 0.453 | 0.709 | 0.479 | 2.177 | 0.031 |
| PCR | AD | 0.970 | 0.333 | -2.175 | 0.031 | -0.547 | 0.585 | 1.736 | 0.084 |
| PLIC | AD | 1.104 | 0.271 | -1.425 | 0.155 | -0.097 | 0.922 | 2.650 | 0.009 |
| PTR | AD | -1.405 | 0.161 | -1.014 | 0.311 | 1.588 | 0.114 | 1.027 | 0.306 |
| RLIC | AD | 0.981 | 0.328 | -1.504 | 0.134 | 0.187 | 0.852 | 0.622 | 0.535 |
| SCC | AD | 1.795 | 0.074 | -2.570 | 0.011 | 0.185 | 0.853 | 0.723 | 0.470 |
| SCR | AD | 0.509 | 0.611 | -1.705 | 0.089 | -0.855 | 0.394 | 1.825 | 0.069 |
| SFO | AD | 1.019 | 0.309 | -1.039 | 0.300 | 1.015 | 0.311 | 1.514 | 0.132 |
| SLF | AD | 1.365 | 0.173 | -2.094 | 0.037 | 0.179 | 0.858 | 2.189 | 0.030 |
| SS | AD | -0.157 | 0.875 | -0.168 | 0.867 | 1.064 | 0.288 | 0.905 | 0.366 |
| UNC | AD | 0.753 | 0.452 | -1.302 | 0.194 | -0.323 | 0.747 | 0.585 | 0.559 |

Significant results are highlighted in bold ($p < 0.002$). Abbreviation: FA = fractional anisotropy, MD = mean diffusivity, RD = radial diffusivity, AD = axial diffusivity, AOO = age of onset, DOI = duration of illness, PANSS = positive and negative syndrome scale. Abbreviations for tracts see Table 1 in main text.

**Table S15| Meta-analytic results for fractional anisotropy differences between adolescents with early-onset psychosis and healthy controls.**

| Tract | Cohen's *d* | S.E. | lCI | uCI | z-value | p-value | $I^2$ | $H^2$ | $Tau^2$ |
|---|---|---|---|---|---|---|---|---|---|
| ACR | -0.12 | 0.14 | -0.39 | 0.14 | -0.91 | 0.365 | 54.2 | 2.18 | 0.08 |
| ALIC | -0.18 | 0.11 | -0.4 | 0.05 | -1.55 | 0.121 | 34.99 | 1.54 | 0.04 |
| AverageFA | -0.19 | 0.14 | -0.46 | 0.08 | -1.4 | 0.162 | 53.92 | 2.17 | 0.08 |
| BCC | -0.11 | 0.1 | -0.31 | 0.08 | -1.14 | 0.253 | 16.37 | 1.2 | 0.01 |
| CC | -0.16 | 0.14 | -0.43 | 0.11 | -1.14 | 0.256 | 55.22 | 2.23 | 0.08 |
| CGC | -0.11 | 0.14 | -0.38 | 0.16 | -0.81 | 0.418 | 54.97 | 2.22 | 0.08 |
| CGH | -0.05 | 0.14 | -0.32 | 0.22 | -0.36 | 0.720 | 56.25 | 2.29 | 0.08 |
| CR | -0.16 | 0.14 | -0.42 | 0.11 | -1.16 | 0.247 | 54.28 | 2.19 | 0.08 |
| CST | -0.04 | 0.09 | -0.22 | 0.13 | -0.45 | 0.651 | 0 | 1 | 0 |
| EC | -0.07 | 0.14 | -0.34 | 0.21 | -0.48 | 0.634 | 56.13 | 2.28 | 0.08 |
| FX | -0.16 | 0.11 | -0.37 | 0.06 | -1.44 | 0.149 | 27.6 | 1.38 | 0.03 |
| FXST | -0.2 | 0.12 | -0.43 | 0.03 | -1.7 | 0.090 | 38.9 | 1.64 | 0.04 |
| GCC | -0.18 | 0.14 | -0.47 | 0.1 | -1.28 | 0.202 | 59.39 | 2.46 | 0.1 |
| IC | -0.21 | 0.11 | -0.43 | 0 | -1.95 | 0.051 | 29.63 | 1.42 | 0.03 |
| IFO | -0.14 | 0.1 | -0.33 | 0.05 | -1.4 | 0.163 | 13.35 | 1.15 | 0.01 |
| PCR | -0.25 | 0.11 | -0.48 | -0.03 | -2.2 | 0.028 | 35.58 | 1.55 | 0.04 |
| PLIC | -0.15 | 0.09 | -0.33 | 0.04 | -1.55 | 0.122 | 8.09 | 1.09 | 0.01 |
| PTR | -0.17 | 0.12 | -0.41 | 0.07 | -1.36 | 0.174 | 45.4 | 1.83 | 0.05 |
| RLIC | -0.2 | 0.1 | -0.39 | -0.01 | -2.05 | 0.040 | 13.77 | 1.16 | 0.01 |
| SCC | -0.11 | 0.14 | -0.38 | 0.17 | -0.76 | 0.445 | 56.63 | 2.31 | 0.09 |
| SCR | -0.08 | 0.12 | -0.32 | 0.16 | -0.67 | 0.502 | 44.17 | 1.79 | 0.05 |
| SFO | -0.26 | 0.09 | -0.43 | -0.08 | -2.86 | 0.004 | 0 | 1 | 0 |
| SLF | -0.35 | 0.09 | -0.53 | -0.18 | -3.93 | **8.41e-05** | 0 | 1 | 0 |
| SS | -0.04 | 0.11 | -0.25 | 0.18 | -0.35 | 0.726 | 30.19 | 1.43 | 0.03 |
| UNC | -0.15 | 0.1 | -0.34 | 0.04 | -1.5 | 0.133 | 14.03 | 1.16 | 0.01 |

Significant results are highlighted in bold ($p < 0.002$). Abbreviations: S.E. = standard error, lCI = lower confidence interval, uCI = upper confidence interval. Abbreviations for tracts, see Table 1 in main text.

**Table S16| Direct comparison of meta- and mega-analytically derived effect sizes for case-control FA differences between early-onset psychosis (EOP) and adult schizophrenia (SCZ)**

| | | Cohen's *d* | | S.E. | | | | | |
|---|---|---|---|---|---|---|---|---|---|
| **Tract** | **Contrast** | **EOP** | **SCZ** | **EOP** | **SCZ** | **Diff** | **S.E.$_{Diff}$** | **Z$_{Diff}$** | **p-value** |
| ACR | MetavsMeta | -0.12 | -0.40 | 0.14 | 0.05 | -0.28 | 0.43 | -0.65 | 0.52 |
| | MetavsMega | -0.16 | -0.40 | 0.08 | 0.05 | 0.24 | 0.36 | 0.68 | 0.50 |
| ALIC | MetavsMeta | -0.18 | -0.37 | 0.11 | 0.05 | -0.19 | 0.39 | -0.48 | 0.63 |
| | MetavsMega | -0.25 | -0.37 | 0.08 | 0.05 | 0.12 | 0.36 | 0.34 | 0.74 |
| AverageFA | MetavsMeta | -0.19 | -0.42 | 0.14 | 0.04 | -0.23 | 0.43 | -0.54 | 0.59 |
| | MetavsMega | -0.30 | -0.42 | 0.08 | 0.04 | 0.12 | 0.35 | 0.34 | 0.74 |
| BCC | MetavsMeta | -0.11 | -0.39 | 0.10 | 0.05 | -0.28 | 0.38 | -0.74 | 0.46 |
| | MetavsMega | -0.24 | -0.39 | 0.08 | 0.05 | 0.15 | 0.36 | 0.43 | 0.67 |
| CC | MetavsMeta | -0.16 | -0.40 | 0.14 | 0.05 | -0.24 | 0.43 | -0.56 | 0.58 |
| | MetavsMega | -0.28 | -0.40 | 0.08 | 0.05 | 0.12 | 0.36 | 0.34 | 0.73 |
| CGC | MetavsMeta | -0.11 | -0.27 | 0.14 | 0.05 | -0.16 | 0.43 | -0.37 | 0.71 |
| | MetavsMega | -0.20 | -0.27 | 0.08 | 0.05 | 0.07 | 0.36 | 0.18 | 0.85 |
| CGH | MetavsMeta | -0.05 | -0.11 | 0.14 | 0.04 | -0.06 | 0.43 | -0.14 | 0.89 |
| | MetavsMega | -0.08 | -0.11 | 0.08 | 0.04 | 0.03 | 0.35 | 0.09 | 0.93 |
| CR | MetavsMeta | -0.16 | -0.33 | 0.14 | 0.04 | -0.17 | 0.42 | -0.40 | 0.69 |
| | MetavsMega | -0.22 | -0.33 | 0.08 | 0.04 | 0.11 | 0.35 | 0.32 | 0.75 |
| CST | MetavsMeta | -0.04 | -0.04 | 0.09 | 0.04 | 0.00 | 0.36 | 0.00 | 1.00 |
| | MetavsMega | -0.05 | -0.04 | 0.08 | 0.04 | -0.01 | 0.35 | -0.02 | 0.98 |
| EC | MetavsMeta | -0.07 | -0.21 | 0.14 | 0.04 | -0.14 | 0.42 | -0.33 | 0.74 |
| | MetavsMega | -0.19 | -0.21 | 0.08 | 0.04 | 0.02 | 0.35 | 0.06 | 0.95 |
| FX | MetavsMeta | -0.16 | -0.31 | 0.11 | 0.05 | -0.15 | 0.39 | -0.38 | 0.70 |
| | MetavsMega | -0.18 | -0.31 | 0.08 | 0.05 | 0.13 | 0.36 | 0.38 | 0.71 |
| FXST | MetavsMeta | -0.20 | -0.32 | 0.12 | 0.04 | -0.12 | 0.40 | -0.30 | 0.77 |
| | MetavsMega | -0.24 | -0.32 | 0.08 | 0.04 | 0.08 | 0.35 | 0.22 | 0.82 |
| GCC | MetavsMeta | -0.18 | -0.37 | 0.14 | 0.04 | -0.19 | 0.43 | -0.45 | 0.66 |
| | MetavsMega | -0.28 | -0.37 | 0.08 | 0.04 | 0.09 | 0.35 | 0.25 | 0.80 |
| IC | MetavsMeta | -0.21 | -0.18 | 0.11 | 0.04 | 0.03 | 0.39 | 0.08 | 0.94 |
| | MetavsMega | -0.29 | -0.18 | 0.08 | 0.04 | -0.11 | 0.35 | -0.32 | 0.75 |
| IFO | MetavsMeta | -0.14 | -0.11 | 0.10 | 0.04 | 0.03 | 0.37 | 0.08 | 0.94 |
| | MetavsMega | -0.12 | -0.11 | 0.08 | 0.04 | -0.01 | 0.35 | -0.03 | 0.98 |
| PCR | MetavsMeta | -0.25 | -0.25 | 0.11 | 0.04 | 0.00 | 0.38 | 0.00 | 1.00 |
| | MetavsMega | -0.32 | -0.25 | 0.08 | 0.04 | -0.07 | 0.35 | -0.20 | 0.84 |
| PLIC | MetavsMeta | -0.15 | 0.04 | 0.09 | 0.05 | 0.19 | 0.37 | 0.52 | 0.61 |
| | MetavsMega | -0.20 | 0.04 | 0.08 | 0.05 | -0.24 | 0.36 | -0.66 | 0.51 |
| PTR | MetavsMeta | -0.17 | -0.31 | 0.12 | 0.04 | -0.14 | 0.39 | -0.36 | 0.72 |
| | MetavsMega | -0.26 | -0.31 | 0.08 | 0.04 | 0.05 | 0.34 | 0.14 | 0.89 |
| RLIC | MetavsMeta | -0.20 | -0.13 | 0.10 | 0.04 | 0.07 | 0.38 | 0.19 | 0.85 |
| | MetavsMega | -0.27 | -0.13 | 0.08 | 0.04 | -0.14 | 0.35 | -0.39 | 0.69 |

| | | | | | | | | | |
|---|---|---|---|---|---|---|---|---|---|
| SCC | MetavsMeta | -0.11 | -0.22 | 0.14 | 0.05 | -0.11 | 0.43 | -0.25 | 0.80 |
| | MetavsMega | -0.21 | -0.22 | 0.08 | 0.05 | 0.01 | 0.36 | 0.04 | 0.97 |
| SCR | MetavsMeta | -0.08 | -0.15 | 0.12 | 0.03 | -0.07 | 0.39 | -0.18 | 0.86 |
| | MetavsMega | -0.15 | -0.15 | 0.08 | 0.03 | 0.00 | 0.34 | 0.01 | 0.99 |
| SFO | MetavsMeta | -0.26 | -0.29 | 0.09 | 0.05 | -0.03 | 0.38 | -0.08 | 0.94 |
| | MetavsMega | -0.31 | -0.29 | 0.08 | 0.05 | -0.02 | 0.37 | -0.05 | 0.96 |
| SLF | MetavsMeta | -0.35 | -0.22 | 0.09 | 0.04 | 0.13 | 0.36 | 0.36 | 0.72 |
| | MetavsMega | -0.37 | -0.22 | 0.08 | 0.04 | -0.15 | 0.35 | -0.43 | 0.67 |
| SS | MetavsMeta | -0.04 | -0.30 | 0.11 | 0.04 | -0.26 | 0.39 | -0.67 | 0.50 |
| | MetavsMega | -0.15 | -0.30 | 0.08 | 0.04 | 0.15 | 0.35 | 0.44 | 0.66 |
| UNC | MetavsMeta | -0.15 | -0.16 | 0.10 | 0.03 | -0.01 | 0.36 | -0.03 | 0.98 |
| | MetavsMega | -0.19 | -0.16 | 0.08 | 0.03 | -0.03 | 0.34 | -0.08 | 0.93 |

Abbreviations: S.E. = standard error. Abbreviations for tracts see Table 1 in main text.
